# Protective effectiveness of prior SARS-CoV-2 infection and hybrid immunity against Omicron infection and severe disease: a systematic review and meta-regression

**DOI:** 10.1101/2022.10.02.22280610

**Authors:** Niklas Bobrovitz, Harriet Ware, Xiaomeng Ma, Zihan Li, Reza Hosseini, Christian Cao, Anabel Selemon, Mairead Whelan, Zahra Premji, Hanane Issa, Brianna Cheng, Laith J. Abu Raddad, David Buckeridge, Maria Van Kerkhove, Vanessa Piechotta, Melissa Higdon, Annelies Wilder-Smith, Isabel Bergeri, Daniel Feikin, Rahul K. Arora, Minal Patel, Lorenzo Subissi

**Affiliations:** Temerty Faculty of Medicine, University of Toronto, Toronto, Canada; Department of Critical Care Medicine, University of Calgary, Canada; Centre for Health Informatics, Cumming School of Medicine, University of Calgary, Canada; Institute of Health Policy Management and Evaluation, University of Toronto, Toronto, Canada; Department of Bioengineering, University of California, Berkeley, United States of America; School of Population and Public Health, University of British Columbia, Vancouver, Canada; Libraries, University of Victoria, Victoria, Canada; Institute of Health Informatics, University College London, United Kingdom; Infectious Disease Epidemiology Group, Weill Cornell Medicine–Qatar, Cornell University, Doha, Qatar; Department of Epidemiology and Biostatistics, School of Population and Global Health, McGill University, Montreal, Canada; Health Emergencies Programme, World Health Organization, Geneva, Switzerland; Department of Infectious Disease Epidemiology, Robert Koch Institute, Berlin, Germany; International Vaccine Access Center, Department of International Health, John Hopkins Bloomberg School of Public Health, Baltimore, United States of America; Department of Immunizations, Vaccines and Biologicals, World Health Organization, Geneva, Switzerland; Institute of Biomedical Engineering, University of Oxford, United Kingdom

## Abstract

**Background:** We aimed to systematically review the magnitude and duration of the protective effectiveness of prior infection (PE) and hybrid immunity (HE) against Omicron infection and severe disease.

**Methods:** We searched pre-print and peer-reviewed electronic databases for controlled studies from January 1, 2020, to June 1, 2022. Risk of bias (RoB) was assessed using the Risk of Bias In Non-Randomized Studies of Interventions (ROBINS-I)-Tool. We used random-effects meta-regression to estimate the magnitude of protection at 1-month intervals and the average change in protection since the last vaccine dose or infection from 3 months to 6 or 12 months. We compared our estimates of PE and HE to previously published estimates of the magnitude and durability of vaccine effectiveness (VE) against Omicron.

**Findings:** Eleven studies of prior infection and 15 studies of hybrid immunity were included. For prior infection, there were 97 estimates (27 at moderate RoB and 70 at serious RoB), with the longest follow up at 15 months. PE against hospitalization or severe disease was 82·5% [71·8-89·7%] at 3 months, and 74·6% [63·1-83·5%] at 12 months. PE against reinfection was 65·2% [52·9-75·9%] at 3 months, and 24·7% [16·4-35·5%] at 12 months. For HE, there were 153 estimates (78 at moderate RoB and 75 at serious RoB), with the longest follow up at 11 months for primary series vaccination and 4 months for first booster vaccination. Against hospitalization or severe disease, HE involving either primary series vaccination or first booster vaccination was consistently >95% for the available follow up. Against reinfection, HE involving primary series vaccination was 69·0% [58·9-77·5%] at 3 months after the most recent infection or vaccination, and 41·8% [31·5-52·8%] at 12 months, while HE involving first booster vaccination was 68·6% [58·8-76·9%] at 3 months, and 46·5% [36·0-57·3%] at 6 months. Against hospitalization or severe disease at 6 months, hybrid immunity with first booster vaccination (effectiveness 95·3% [81·9-98·9%]) or with primary series alone (96·5% [90·2-98·8%]) provided significantly greater protection than prior infection alone (80·1% [70·3-87·2%]), first booster vaccination alone (76·7% [72·5-80·4%]), or primary series alone (64·6% [54·5-73·6%]). Results for protection against reinfection were similar.

**Interpretation:** Prior infection and hybrid immunity both provided greater and more sustained protection against Omicron than vaccination alone. All protection estimates waned quickly against infection but remained high for hospitalisation or severe disease. Individuals with hybrid immunity had the highest magnitude and durability of protection against all outcomes, reinforcing the global imperative for vaccination.

**Funding:** WHO COVID-19 Solidarity Response Fund and the Coalition for Epidemic Preparedness Innovations.

**Research in context:** *Evidence before this study:* The global emergence and rapid spread of Omicron (B.1.1.529) variant of concern, characterized by their ability to escape immunity, has required scientists and policymakers to reassess the population protection against Omicron infection and severe disease. So far, few systematic reviews have incorporated data on Omicron, and none have examined the protection against Omicron conferred by hybrid immunity (i.e. the immunity gained from the combination of vaccination and prior infection) which is now widespread globally. While one preprint has recently reported protection from prior infection over time, no systematic review has systematically compared the magnitude and duration of vaccination, prior infection, and hybrid immunity. A large single-country study has reported that protection from either infection or hybrid immunity against Omicron infection wanes to low levels at 15 months, but is relatively stable against severe disease.

*Added value of this study:* Prior infection and hybrid immunity both provided greater and more sustained protection against Omicron than vaccination alone. Individuals with hybrid immunity had the highest magnitude and durability of protection against all outcomes; protection against severe disease remained above 95% until the end of available follow-up at 11 months after hybrid immunity with primary series and 4 months after hybrid immunity with booster vaccination, and was sustained at these high levels of protection in projections to 12 months and 6 months, respectively.

*Implications of all the available evidence:* These results may serve to tailor guidance on the optimal number and timing of vaccinations. At the public health level, these findings can be combined with data on local infection prevalence, vaccination rates, and their timing. In settings with high seroprevalence, limited resources, and competing health priorities, it may be reasonable to focus on achieving high coverage rates with primary series among individuals who are at higher risk of poor outcome, as this will provide a high level of protection against severe disease for at least one year among those with prior infection. Furthermore, given the waning protection for both infection-and vaccine induced immunity against infection or reinfection, mass vaccination could be timed for roll-out prior to periods of expected increased incidence, such as the winter season. At the individual level, these results can be combined with knowledge of a person’s infection and vaccination history. A six-month delay in booster may be justified after the last infection or vaccination for individuals with a known prior infection and full primary series vaccination. Further follow-up of the protective effectiveness of hybrid immunity against hospitalization or severe disease for all vaccines is needed to clarify how much waning of protection might occur with longer duration since the last infection or vaccination. Producing estimates of protection for new variant-containing vaccines will be crucial for COVID-19 vaccine policy and decision-making bodies. Policy makers considering the use and timing of vaccinations should include the local extent of past infection, the protection conferred by prior infection or hybrid immunity, and the duration of this protection as key considerations to inform their decision-making.

## Introduction

Limiting the extent of Severe Acute Respiratory Coronavirus-2 (SARS-CoV-2) infection and preventing severe COVID-19 remains a priority at the global scale. Immunologic protection from SARS-CoV-2 can result from prior infection or vaccination.^1,2^ However, estimating the magnitude and durability of protection in the population has become a challenge due to the Omicron surge, which resulted in many individuals with a combination of infection and vaccination (“hybrid immunity”), varying rates and timing of past infection and vaccination, multiple types of vaccination and numbers of doses, and variants of concern (VOC) that can escape pre-existing immunity.^3,4^

Systematic reviews of SARS-CoV-2 vaccine effectiveness (VE) studies have provided clarity on the durability of protection for different VOCs.^5,6^ These studies have compared protection amongst vaccinated persons to unvaccinated persons and compared protection between different numbers of doses. However, there are gaps in the literature on protection conferred by prior infection, both among persons who have been infected but not vaccinated (PE - prior infection effectiveness) and protection among persons who have been infected and vaccinated (HE - hybrid immunity effectiveness). A recent preprint systematic review has estimated the durability of PE.^7^ However, no systematic review has estimated the durability of protection from HE, or compared the durability of the different types of protection (VE, PE, HE).

Here, we systematically reviewed the evidence for the magnitude and duration of PE and HE against multiple clinical outcomes of SARS-CoV-2 infection by the Omicron variant. We also sought to examine the protection conferred by different amounts of vaccination among individuals with prior infection (VE-infected) and the protection conferred by prior infection among individuals with prior vaccination (PE-vaccinated). For comprehensiveness, we compared our estimates of PE and HE to previously-published estimates of the magnitude and durability of VE against Omicron.

## Methods

### Methodological guidance and registration

This systematic review and meta-regression was registered with PROSPERO (CRD42022318605), conducted in alignment with the Cochrane Handbook for Systematic Reviews of Interventions^8^, and reported according to the PRISMA guidelines (Supplementary File S1).^9^

### Search strategy and selection criteria

We searched MEDLINE (Ovid), Embase (Ovid), Web of Science (Core Collection), ClinicalTrials.gov, Cochrane Central Register of Controlled Trials (Ovid), WHO COVID-19 database, and Europe PMC (limited to pre-prints) from 1 January 2020 to 1 June 2022 (Supplementary File S2). Pre-print articles that were identified during this period and subsequently updated or published as peer-review articles between 1 June 2022 and 15 July 2022 were also included. Additional articles were identified via recommendations from corresponding authors of previous reviews on vaccine effectiveness and investigators in the WHO Solidarity 2 network.

We included studies examining protection against Omicron reinfection, where the comparator group was people with prior infection with any SARS-CoV-2 variant or hybrid immunity and the control group was immune-naive, previously infected individuals, or previously vaccinated individuals (Supplementary File S3). Infection due to the Omicron variant was determined by genomic sequencing or inferred based on periods when the variants were predominant according to GISAID.^10^ Included study designs were test-negative case-control, traditional case-control, cross-sectional, cohort, non-randomized controlled trials, and randomized controlled trials. We included people of any age in any setting.

We excluded case reports, case series, incomplete randomized controlled trials, and review papers. We also excluded articles reporting identical information to previously included articles as duplicates, and excluded the pre-print version of articles subsequently published in peer-reviewed journals. Studies were excluded if they did not report evidence of previously confirmed SARS-CoV-2 cases or did not report the period of time between the index infection and reinfection.

### Outcome definition and comparison groups

SARS-CoV-2 reinfection was defined as a possible, probable, or confirmed reinfection case per adapted WHO definitions; infections with laboratory confirmation were considered confirmed cases.^11^ Partial primary series and full primary series vaccination were defined according to the original trials (Supplementary File S3). First booster vaccination included individuals ≥ 7 days from receipt of the first booster dose (Supplementary File S3).

Estimates of VE (i.e., vaccine vs. immune naive) against Omicron variant were obtained from a systematic review and meta-regression involving 15 studies (n=15 studies of primary series studies, n=12 first booster studies).^6^ The VE estimates were compared to PE and HE estimates generated in our analysis. Details of these comparisons are reported in Supplementary File S3.

### Data extraction

Data was extracted by one reviewer and verified by a second reviewer. We extracted data for each outcome (infection [asymptomatic/symptomatic disease], hospitalization, or severe disease [a combination of the WHO definitions of severe, critical, or fatal COVID-19^12^), stratifying by age, sex, vaccine type, variant of concern causing the index infection, and severity of the index infection. See Supplementary File S3 for detailed definitions of outcomes.

### Risk of bias assessment

Risk of bias was assessed using the Risk of Bias In Non-Randomized Studies of Interventions (ROBINS-I)-Tool.^13^ Assessments were completed for each outcome reported in each study. Two reviewers independently completed the assessments. Conflicts were resolved by discussion and consensus.

### Data analysis

Data was analyzed using R statistical software version 4.1.2.^14^ Four effect measures were calculated based on different comparisons of immunity status: protective effectiveness of prior infection (PE); protective effectiveness of hybrid immunity (HE); vaccine effectiveness (full series or booster) among individuals with prior infection (VE-infected); and protective effectiveness of prior infection among individuals with prior vaccination (PE-vaccinated). These immunity comparisons and effect measures are mapped in Figure 1 and listed in full with descriptions of their epidemiological significance in Supplementary File S4. These measures were calculated for the two outcomes of interest: infection (asymptomatic or symptomatic) and hospitalization or severe disease. The primary outcomes were the PE, HE, VE-infected, and PE-vaccinated against COVID-19 hospitalization or severe disease at discrete time points. The secondary outcomes were the PE, HE, VE-infected, and PE-vaccinated against Omicron infection at discrete time points. For prior infection, we used estimates starting from two months after the primary infection, reflecting the length required for a possible reinfection. For hybrid immunity, we used estimates starting from two months after the primary infection or ≥ 7 days after the most recent vaccination (cutoff threshold varied by vaccine according to the original trials [Supplementary File S3]).

**Figure 1.**
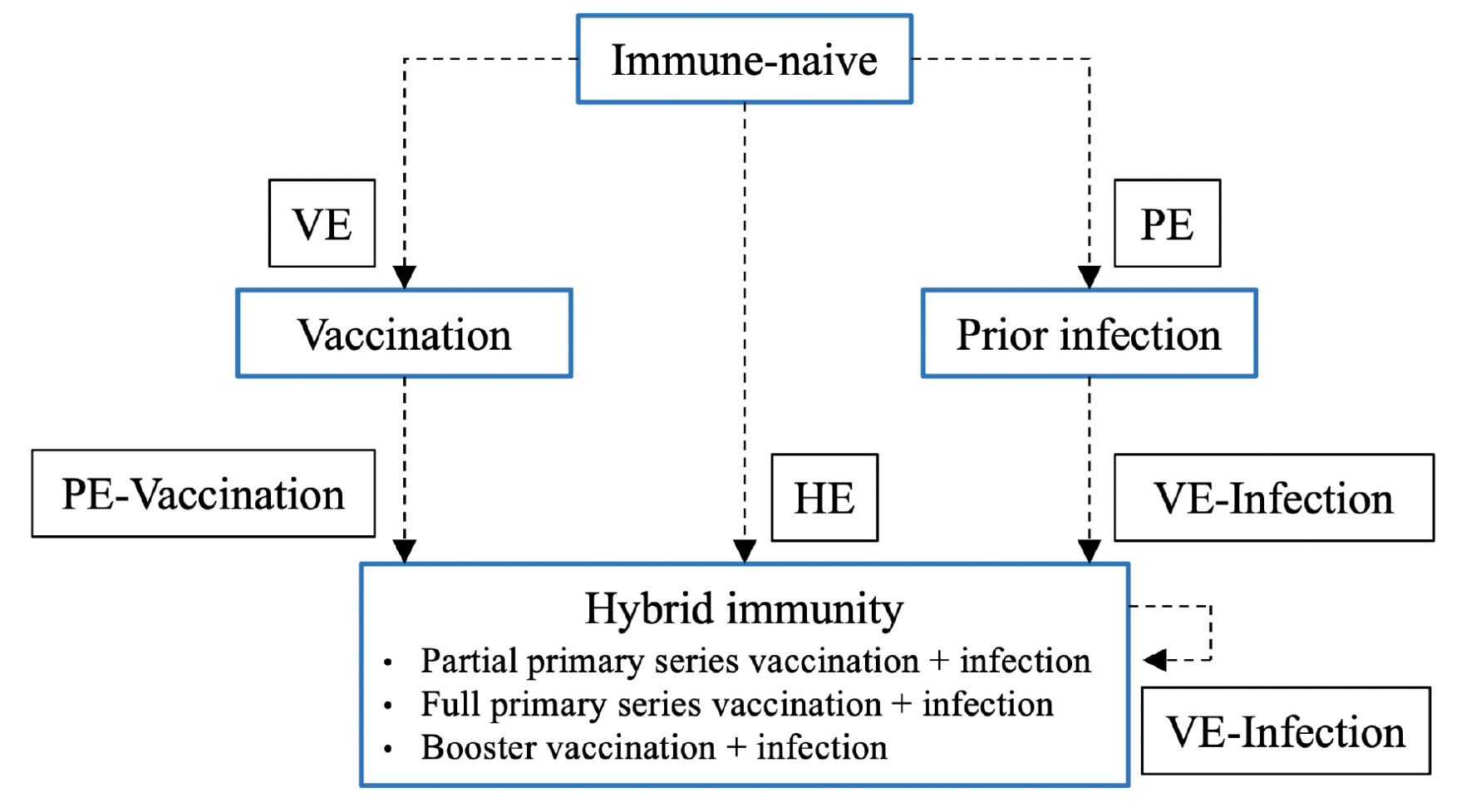
Conceptual map of effect measures derived from different immunity status comparisons. The arrow indicates the comparison informing the measure of effect, with protection in the group at the tip of the arrow compared to the group at the tail of the arrow. Measures of effect where exposure is compared to immune naive: VE - vaccine effectiveness (not assessed in this study); PE - protective effectiveness of prior infection; HE - protective effectiveness of hybrid immunity. Measures of effect where exposure is compared to other immunity states: VE-infected - vaccine effectiveness among individuals with prior infection, including comparisons of hybrid immunity with different levels of vaccinations (e.g., full primary series vaccination and infection vs. booster vaccination and infection); PE-vaccinated - protective effectiveness of prior infection among individuals with prior vaccination. For PE-vaccinated, when the exposure and comparison groups have an equivalent number of vaccinations the difference between groups is prior infection. It is also possible that there may be a non-equivalent number of vaccinations between groups. When the comparison group has a greater number of vaccinations than the hybrid immunity exposure group this measure helps to determine whether prior infection serves as a substitute for additional vaccination.

For prior infection and hybrid immunity, using the log-odds meta-regression model, we generated estimates of effectiveness in successive months as well as percentage point changes in the effectiveness from 3 to 6 months and 3 to 12 months, with 95% confidence intervals for the changes calculated by the bootstrap method. For hybrid immunity, we predicted estimates 1 month and 2 months beyond final follow-up times, yielding estimates of protection at 12 months for hybrid immunity with primary series and 6 months for hybrid immunity with booster vaccination, respectively. For a given outcome measure, if there was insufficient data for meta-regression over time then estimates were pooled via meta-analysis (same log-odds model with no time covariate).^15–17^ These estimates represented an average of the timepoints reported in different studies. More details on modeling technique can be found in Supplementary File S3.

Sensitivity analyses were conducted which focused only on severe disease and excluded all-cause hospitalization (Supplementary File S3). There were insufficient data to report results stratified by age, sex, severity of the index infection, and Omicron subvariant causing reinfection.

## Results

In total, 4,268 articles were screened and 895 underwent full-text review (Supplementary File S5). Sixteen unique articles reporting data for 26 studies were included for analysis.^18–33^ Seven articles reported data for multiple studies, defined by multiple sets of study methods and independent populations with unique estimates of protection. Eleven studies reported on prior infection and 15 studies reported on hybrid immunity, which included six reporting on both. A summary of study characteristics is reported in Table 1. Individual study characteristics are reported in Supplementary File S6 and S7. The most common reason for increased risk of bias was incomplete adjustment for confounding factors (e.g., health seeking behavior including testing frequency, comorbidity, calendar time) and not reporting an *a priori* protocol to enable ruling out reporting bias.

**Table 1.** Study characteristics.

|  | Prior infection studies | Hybrid immunity studies |  |  |  |
| --- | --- | --- | --- | --- | --- |
|  | PE<br>Prior infection vs.<br>immune-naïve<sup>a</sup> | HE<br>Hybrid immunity vs.<br>immune-naïve<sup>a</sup> | VE-Infected<br>Hybrid immunity vs.<br>prior infection or<br>hybrid immunity with<br>fewer vaccinations<sup>a</sup> | PE-Vaccinated<br>Hybrid immunity<br>vs. vaccination<sup>a</sup> | Summary of all<br>hybrid<br>immunity<br>studies |
| Characteristic | N = 11<br>n (%) | N = 9<br>n (%) | N = 7<br>n (%) | N = 1<br>n (%) | N = 15<br>n (%) |
| Study design |  |  |  |  |  |
| Cohort | 2 (18.2%) | 0 (0%) | 2 (29%) | 1 (100%) | 3 (20%) |
| Cross-sectional | 1 (9.1%) | 0 (0%) | 0 (0%) | 0 (0%) | 0 (0%) |
| Test-negative design case-control | 8 (73%) | 7 (78%) | 4 (57%) | 0 (40%) | 9 (60%) |
| Traditional case-control | 0 (0%) | 2 (22%) | 1 (14%) | 0 (0%) | 3 (20%) |
| Total sample size, Median (IQR) | 294,900<br>(83,251, 1,142,605) | 317,110<br>(50,576, 696,439) | 130,073<br>(14,625, 470,984) | 38 | 130,073<br>(14,625, 335,882) |
| Study population |  |  |  |  |  |
| General population | 11 (100%) | 9 (100%) | 6 (86%) | 1 (100%) | 14 (93%) |
| Healthcare workers | 0 (0%) | 0 (0%) | 1 (14%) | 0 (0%) | 1 (6.7%) |
| WHO region |  |  |  |  |  |
| AFR | 0 (0%) | 0 (0%) | 0 (0%) | 0 (0%) | 0 (0%) |
| AMR | 4 (36%) | 3 (33%) | 5 (71%) | 0 (0%) | 6 (40%) |
| EMR | 2 (18%) | 2 (22%) | 0 (0%) | 0 (0%) | 2 (13%) |
| EUR | 5 (45%) | 4 (44%) | 2 (29%) | 1 (100%) | 7 (47%) |
| SEAR | 0 (0%) | 0 (0%) | 0 (0%) | 0 (0%) | 0 (0%) |
| WPR | 0 (0%) | 0 (0%) | 0 (0%) | 0 (0%) | 0 (0%) |
| Reported the sequence of index infection and vaccination | - | 4 (44%) | 5 (71%) | 1 (100%) | 9 (60%) |
| Reported Omicron subvariants | 4 (36%) | 5 (56%) | 1 (14%) | 0 (0%) | 6 (40%) |
| Primary infection dominant variant |  |  |  |  |  |
| Alpha | 1 (9.1%) | 1 (11%) | 0 (0%) | 0 (0%) | 1 (6.7%) |
| Delta | 3 (27%) | 4 (44%) | 0 (0%) | 0 (0%) | 4 (27%) |
| Mixed variant | 9 (82%) | 7 (78%) | 7 (100%) | 1 (100%) | 13 (87%) |
| Primary series vaccination for hybrid immunity<sup>b</sup> | - | 9 | 5 | - | 12 |
| Inactivated: Sinovac-CoronaVac | - | 1 (11%) | 1 (14%) | - | 1 (6.7%) |
| mRNA: Pfizer/BioNTech-Comirnaty | - | 1 (11%) | 1 (14%) | - | 2 (13%) |
| mRNA: Moderna-mRNA-1273 | - | 1 (11%) | 0 (0%) | - | 1 (6.7%) |
| mRNA: Pfizer/BioNTech-Comirnaty + Moderna-mRNA-1273 | - | 2 (22%) | 4 (57%) | - | 5 (33%) |
| NRVV:AstraZeneca-Vaxzevria | - | 1 (11%) | 1 (14%) | - | 1 (6.7%) |
| NRVV: Janssen-Ad26.COV2.S | - | 1 (11%) | 1 (14%) | - | 1 (6.7%) |
| Mixed (Inactivated + NRVV + mRNA)<sup>d</sup> | - | 5 (56%) | 4 (57%) | - | 8 (53%) |
| First booster vaccination for hybrid immunity<sup>b</sup> |  | 7 | 6 | 1 | 12 |
| Inactivated: Sinovac-CoronaVac | - | 1 (11%) | 1 (14%) | 0 (0%) | 1 (6.7%) |
| mRNA: Pfizer/BioNTech-Comirnaty |  | 2 (22%) | 1 (14%) | 0 (0%) | 3 (20%) |
| mRNA: Pfizer/BioNTech-Comirnaty + Moderna-mRNA-1273 | - | 2 (22%) | 4 (57%) | 1 (100%) | 7 (470%) |
| NRVV:AstraZeneca-Vaxzevria | - | 1 (11%) | 1 (14%) | 0 (0%) | 1 (6.7%) |
| NRVV: Janssen-Ad26.COV2.S |  | 1 (11%) | 1 (14%) | 0 (0%) | 1 (6.7%) |
| Mixed (Inactivated + NRVV + mRNA)<sup>d</sup> | - | 2 (22%) | 1 (14%) | 0 (0%) | 3 (20%) |
| Type of reinfection |  |  |  |  |  |
| Confirmed | 3 (27%) | 3 (33%) | 0 (0%) | 0 (0%) | 3 (20%) |
| Probable | 6 (55%) | 6 (67%) | 7 (100%) | 0 (0%) | 11 (73%) |
| Possible | 2 (18%) | 0 (0%) | 0 (0%) | 1 (100%) | 1 (6.7%) |
| Reinfection severity<sup>c</sup> |  |  |  |  |  |
| Any reinfection | 10 (91%) | 8 (89%) | 6 (86%) | 1 (100%) | 13 (87%) |
| Severe disease (includes hospitalisation) | 7 (64%) | 6 (67%) | 3 (43%) | 0 (0%) | 7 (47%) |
| Severe disease (as per WHO definition) | 4 (36%) | 4 (44%) | 0 (0%) | 0 (0%) | 4 (27%) |

<sup>a</sup>Seven studies reported results on both the protection conferred by prior infection and hybrid immunity. Hybrid immunity studies may be included in more than one column (HE, VE-Infected, PE-Vaccinated) where they report multiple effect measures. <sup>b</sup>Percentages in the hybrid immunity studies columns may add up to more than 100% where studies report multiple vaccine types. <sup>c</sup>Percentages may add up to more than 100% where studies report multiple reinfection severity levels. <sup>d</sup>The mixed vaccination category included the following types of vaccines: inactivated (Sinopharm-BBIBP-CorV); non-replicating viral vector (NRVV) (AstraZeneca-Vaxzevria, Janssen-Ad26.COV2.S, Gamaleja-Sputnik-V); and mRNA (Pfizer/BioNTech-Comirnaty, Moderna-mRNA-1273).

### PE: protective effectiveness of prior infection

Eleven studies involving a median of 294,900 participants (IQR 83,251 - 1,142,605) reported the protection conferred by prior infection (Supplementary File S8). Of the 97 estimates reported, 27 (27·8%) were at moderate risk of bias and 70 (72·2%) were at serious risk of bias (Supplementary File S8).

Of the 11 studies that evaluated PE, six reported protection against hospitalization or severe disease and ten studies reported protection against reinfection over time, with the longest follow-up at 15 months (Table 2, Figure 2). PE against hospitalization or severe disease was 82·5% [71·8-89·7%] at 3 months; this was stable over time reaching 74·6% [63·1-83·5%] at 12 months (−7·8 % [-20·9 to +12·1]) and 71·6% [57·1%-82·6%] at 15 months. PE against reinfection was 65·2% [52·9-75·9%] at 3 months, dropping to 24·7% [16·4-35·5%] at 12 months (−40·5% [-33·9 to -51·9]) and 15·5% [9·9-23·6%] at 15 months.

**Table 2.**
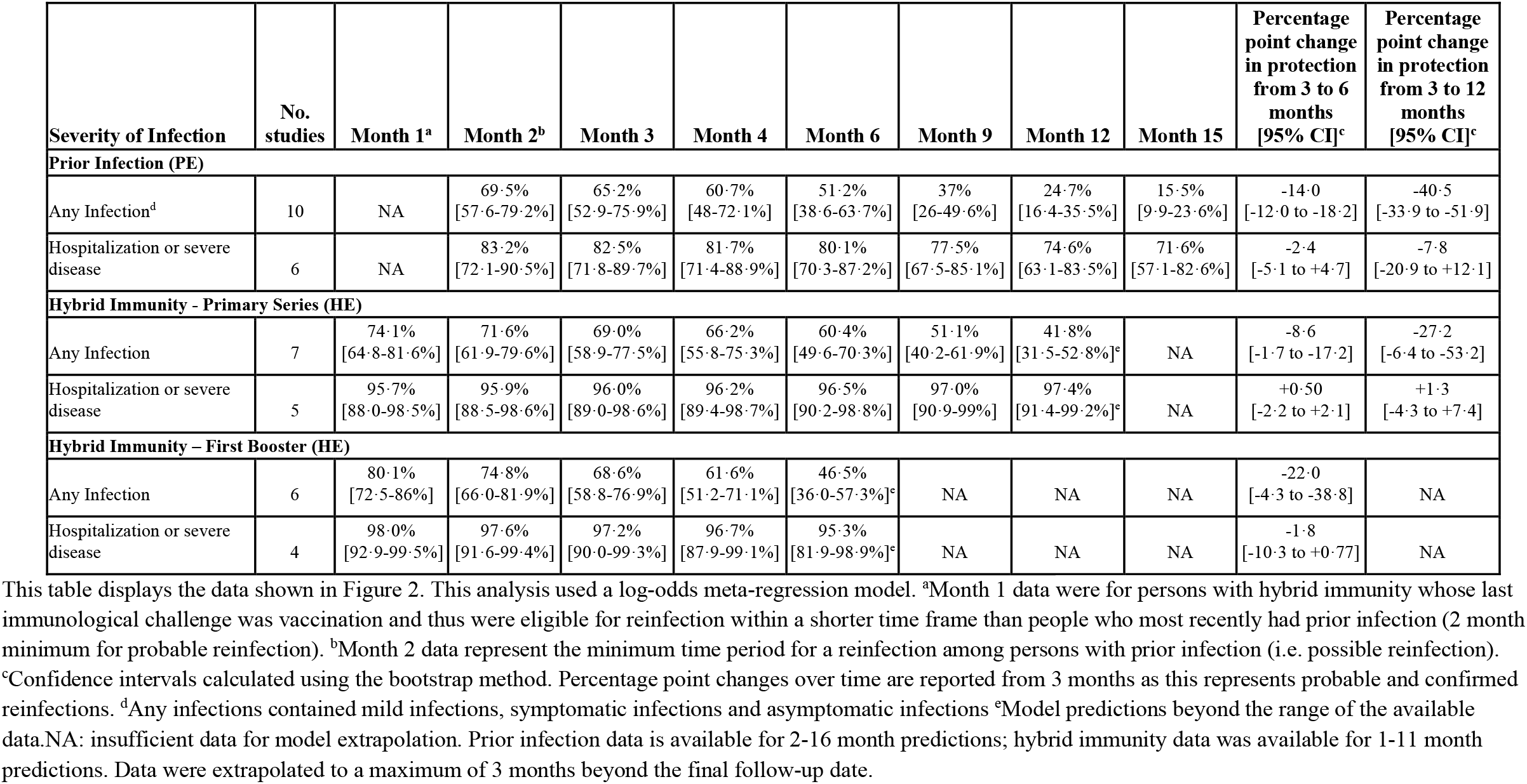
Protection conferred by prior infection and hybrid immunity compared to immune naive.

| Severity of Infection | No. studies | Month 1<sup>a</sup> | Month 2<sup>b</sup> | Month 3 | Month 4 | Month 6 | Month 9 | Month 12 | Month 15 | Percentage point change in protection from 3 to 6 months [95% CI]<sup>c</sup> | Percentage point change in protection from 3 to 12 months [95% CI]<sup>c</sup> |
| --- | --- | --- | --- | --- | --- | --- | --- | --- | --- | --- | --- |
| Prior Infection (PE) |  |  |  |  |  |  |  |  |  |  |  |
| Any Infection<sup>d</sup> | 10 | NA | 69·5%<br>[57·6-79·2%] | 65·2%<br>[52·9-75·9%] | 60·7%<br>[48-72·1%] | 51·2%<br>[38·6-63·7%] | 37%<br>[26-49·6%] | 24·7%<br>[16·4-35·5%] | 15·5%<br>[9·9-23·6%] | -14·0<br>[-12·0 to -18·2] | -40·5<br>[-33·9 to -51·9] |
| Hospitalization or severe disease | 6 | NA | 83·2%<br>[72·1-90·5%] | 82·5%<br>[71·8-89·7%] | 81·7%<br>[71·4-88·9%] | 80·1%<br>[70·3-87·2%] | 77·5%<br>[67·5-85·1%] | 74·6%<br>[63·1-83·5%] | 71·6%<br>[57·1-82·6%] | -2·4<br>[-5·1 to +4·7] | -7·8<br>[-20·9 to +12·1] |
| Hybrid Immunity - Primary Series (HE) |  |  |  |  |  |  |  |  |  |  |  |
| Any Infection | 7 | 74·1%<br>[64·8-81·6%] | 71·6%<br>[61·9-79·6%] | 69·0%<br>[58·9-77·5%] | 66·2%<br>[55·8-75·3%] | 60·4%<br>[49·6-70·3%] | 51·1%<br>[40·2-61·9%] | 41·8%<br>[31·5-52·8%]<sup>e</sup> | NA | -8·6<br>[-1·7 to -17·2] | -27·2<br>[-6·4 to -53·2] |
| Hospitalization or severe disease | 5 | 95·7%<br>[88·0-98·5%] | 95·9%<br>[88·5-98·6%] | 96·0%<br>[89·0-98·6%] | 96·2%<br>[89·4-98·7%] | 96·5%<br>[90·2-98·8%] | 97·0%<br>[90·9-99%] | 97·4%<br>[91·4-99·2%]<sup>e</sup> | NA | +0·50<br>[-2·2 to +2·1] | +1·3<br>[-4·3 to +7·4] |
| Hybrid Immunity – First Booster (HE) |  |  |  |  |  |  |  |  |  |  |  |
| Any Infection | 6 | 80·1%<br>[72·5-86%] | 74·8%<br>[66·0-81·9%] | 68·6%<br>[58·8-76·9%] | 61·6%<br>[51·2-71·1%] | 46·5%<br>[36·0-57·3%]<sup>e</sup> | NA | NA | NA | -22·0<br>[-4·3 to -38·8] | NA |
| Hospitalization or severe disease | 4 | 98·0%<br>[92·9-99·5%] | 97·6%<br>[91·6-99·4%] | 97·2%<br>[90·0-99·3%] | 96·7%<br>[87·9-99·1%] | 95·3%<br>[81·9-98·9%]<sup>e</sup> | NA | NA | NA | -1·8<br>[-10·3 to +0·77] | NA |

**Figure 2.**
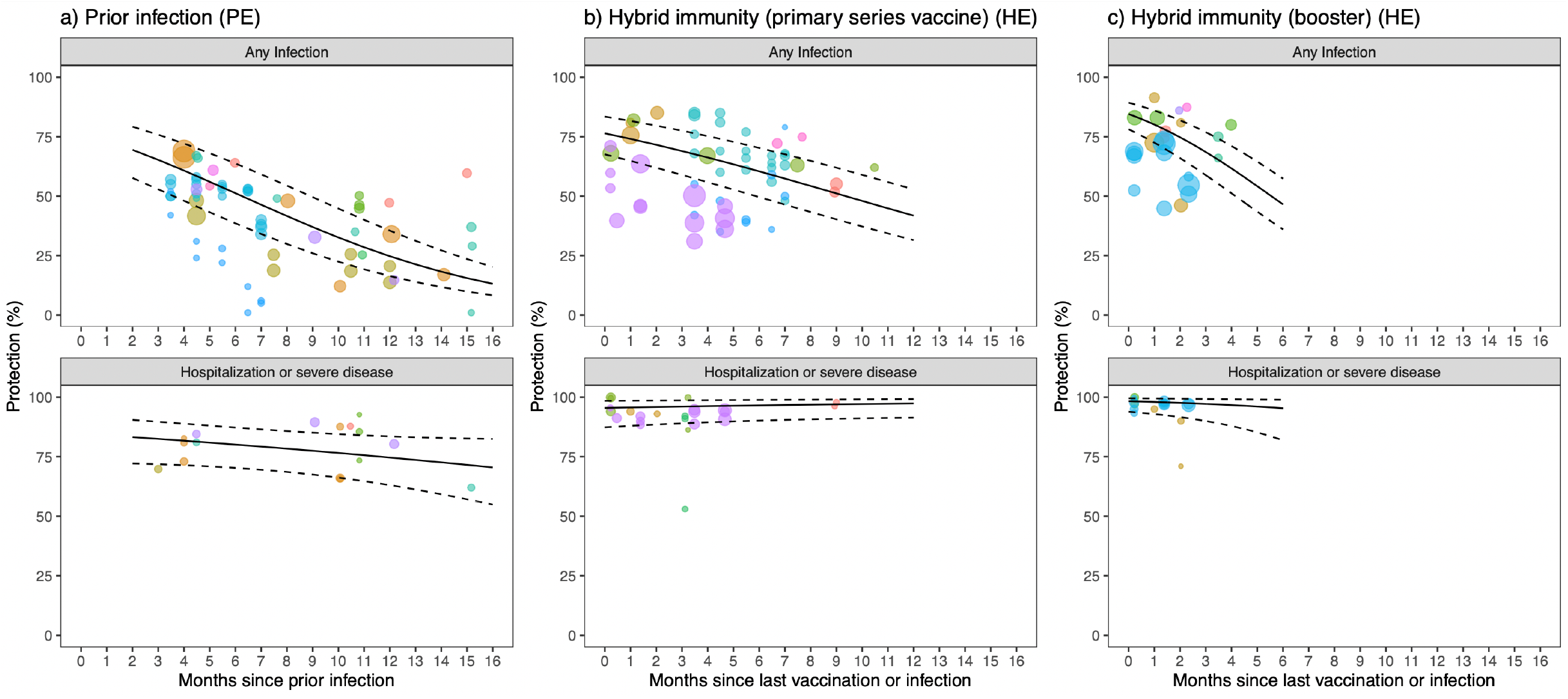
Protection against Omicron conferred by prior infection or hybrid immunity compared to immune naive over time. This analysis uses a log-odds meta-regression model. Points of the same color represent estimates from the same study. The diameter of points varies with the sample size of the study. Dotted lines represent 95% confidence intervals.

Sensitivity analysis that only included studies reporting severe disease, as defined in the methods, showed similar results to the main analysis (Supplementary File S9, S10). Definitions of severe disease used in each study are reported in Supplementary File S11.

Three PE studies reported reinfection results by age subgroups (Supplementary File S12). No differences by age were reported.

Subgroup meta-regression analysis by variant causing the index infection was limited to protection from Alpha, Delta, and mixed pre-Omicron variants, including Alpha and Delta. Data at 6 months showed no differences in the level of protection by the variant causing the index infection (Supplementary File S13).

### HE: protective effectiveness of hybrid immunity

Nine studies involving a median of 317,110 participants (IQR 50,576 - 696,439) reported the protection conferred by hybrid immunity compared to immune-naive individuals, all of which reported on infection in combination with primary series vaccination. Seven studies reported on infection in combination with first booster vaccination. Of the 153 estimates reported, 78 (51·0%) were at moderate risk of bias and 75 (49·0%) were at serious risk of bias (Supplementary File S8).

Of the nine studies that evaluated HE with primary series vaccination, five studies reported protection against hospitalization or severe disease and seven studies reported protection against reinfection over time, with the longest follow-up at 11 months (Table 2, Figure 2). HE (primary series) against hospitalization or severe disease was 96·0% [89·0-98·6%] at 3 months and remained stable at 97·4% [91·4-99·2%] in projections at 12 months (+1·3% [-4·3 to +7·4]). HE (primary series) against reinfection was 69·0% [58·9-77·5%] at 3 months, dropping to 41·8% [31·5-52·8%] at 12 months (−27·2% [-6·4 to -53·2]).

Of the seven studies that evaluated the HE with first booster vaccination, four studies reported HE against hospitalization or severe disease and six studies reported HE against reinfection over time, with longest follow-up of 4 months. HE against hospitalization or severe disease was 97·2% [90·0-99·3%] at 3 months, and remained stable at 95·3% [81·9-98·9%] in projections at 6 months (−1·8 percentage points [-10·3 to +0·77]). HE against reinfection was 68·6% [58·8-76·9%] at 3 months, dropping to 46·5% [36·0-57·3%] at 6 months (−22·0 percentage points [-4·3 to -38·8]).

Sensitivity analysis that only included studies reporting severe disease showed similar results to the main analysis (Supplementary File S9, S10).

Three HE studies reported reinfection results by age subgroups (Supplementary File S12). One study reported that children aged 12-17 and older adults aged 50-69 had more protection from HE with partial primary series and full primary series than adults aged 18-49. No other differences by age were reported.

Subgroup meta-regression analysis by variant causing the index infection was limited to protection from Alpha, Delta, and mixed pre-omicron variants, including Alpha and Delta. Data at 6 months showed no differences in the level of protection by the variant causing the index infection (Supplementary File S13).

Subgroup meta-regression analysis showed differences in the level of protection and patterns of waning protection over time by vaccine type (Supplementary File S14). At 3 months after last immunological challenge (i.e. vaccination or infection) HE against hospitalization or severe disease was initially high (>88%) across all vaccine types. At 6 months the available data showed a maintenance of protection for mRNA vaccines (98·1% [91·2-99·6%]) but a greater reduction in protection for a mixture of vaccines (75·5% [13·7-98·4%]). HE against reinfection was highest with a primary series of mRNA vaccines (60·9% [48·3-72·2%], n=5 studies), followed by mRNA + non-replicating viral vector (60·1% [43·0-75·1%], n=3 studies), non-replicating viral vectors vaccines (40·7% [39·6-41·7%], n=1 study) and inactivated vaccines (36·2% [34·9-37·4%], n=1 study). There were significant monthly reductions in reinfection from 3 to 6 months for mRNA vaccines but no change for mixtures of vaccines, with data being limited for other vaccine types.

### VE-infected: vaccine effectiveness among individuals with prior infection

Seven studies involving a median of 130,073 participants (IQR 14,625 -470,984) reported the VE-infected, of which five studies reported on infection in combination with primary series vaccination series, and six reported on infection in combination with first booster vaccination. Of the 90 estimates reported, 51 (56·7%) were at moderate risk of bias and 39 (43.3%) were at serious risk of bias (Supplementary File S8).

Since there was insufficient data for meta-regression over time, estimates of VE-infected were derived by conducting a meta-analysis. Longest follow-up was 11 months. Meta-analysis showed that hybrid immunity conferred a significant gain in protection compared to prior infection alone (i.e., protection relative to an active comparator), including with partial primary series (n=4 studies; 28·9% [14·4-49·6%] against hospitalization or severe disease, 59·0% [51·5-66·1%] against reinfection); full primary series (n=5 studies; 57·7% [28·6-82·2%] against hospitalization or severe disease, 46·1% [30·6-62·4%] against reinfection), and first booster vaccination (n=5 studies; 80·1% [48·6-94·5%] against hospitalization or severe disease, 46·5% [24·6-69·9%] against reinfection) (Table 3).

**Table 3.** Protection conferred by hybrid immunity compared to prior infection (VE-Infected), hybrid immunity (VE-Infected), or vaccination (PE-Vaccinated)

| Exposure | Comparator | Outcome<sup>a</sup> | No. estimates | Pooled relative protection [95% CI] |
| --- | --- | --- | --- | --- |
| **Hybrid immunity vs. Prior infection (VE-Infected)** |  |  |  |  |
| Prior infection + partial primary series vaccine | Prior infection | Any Infection | 2 | 59·0% [51·5-66·1%] |
| Prior infection + full primary series | Prior infection | Any Infection | 5 | 46·1% [30·6-62·4%] |
| Prior infection + first booster dose | Prior infection | Any Infection | 5 | 46·5% [24·6-69·9%] |
| Prior infection + partial primary series vaccine | Prior infection | Hospitalization or severe disease | 3 | 28·9% [14·4-49·6%] |
| Prior infection + full primary series | Prior infection | Hospitalization or severe disease | 4 | 57·7% [28·6-82·2%] |
| Prior infection + first booster dose | Prior infection | Hospitalization or severe disease | 3 | 80·1% [48·6-94·5%] |
| **Hybrid immunity vs. Hybrid immunity (VE-Infected)** |  |  |  |  |
| Prior infection + full primary series | Prior infection + partial primary series vaccine | Any Infection | 1 | 16·3% [11·1-21·2%] |
| Prior infection + first booster dose | Prior infection + partial primary series vaccine | Any Infection | 2 | 5·8% [0·2-65·2%] |
| Prior infection + first booster dose | Prior infection + primary series | Any Infection | 3 | 40·8% [22·0-62·7%] |
| Prior infection + full primary series | Prior infection + partial primary series vaccine | Hospitalization or severe disease | 2 | 49·6% [19·9-79·7%] |
| Prior infection + first booster dose | Prior infection + partial primary series vaccine | Hospitalization or severe disease | 1 | 67·1% [21·1-86·3%] |
| Prior infection + first booster dose | Prior infection + primary series | Hospitalization or severe disease | 2 | 37·0% [6·2-83·9%] |
| **Hybrid immunity vs. Vaccination (PE-Vaccinated)** |  |  |  |  |
| Prior infection + first booster dose | First booster dose | Any Infection | 1 | 88·9% [29·5-98·2%] |

<sup>a</sup>There were insufficient data for sensitivity analysis.

Five studies compared hybrid immunity at different levels of vaccination (Table 3). In general, results showed that hybrid immunity with a greater number of vaccinations conferred significant gains in protection against both hospitalization or severe disease and reinfection; however, data were limited for some comparisons. Sensitivity analysis restricting to severe disease showed similar results (results not presented).

### PE-vaccinated: protective effectiveness of prior infection among individuals with prior vaccination

One cohort study of 38 older adults (71–101 years of age) conducted in France compared prior infection in combination with first booster vaccination series against first booster vaccination only using BioNTech/Pfizer-BNT162b2 or Moderna-mRNA-1273. After 90 days since the last immunological challenge the estimated PE-vaccinated against Omicron reinfection was 88·9% (29·5-98·2%).

### Comparison of PE and HE estimates to previously published VE estimates^6^

Meta-regression showed that against hospitalization or severe disease at 6 months, HE with first booster vaccination (effectiveness 95·3% [81·9-98·9%]) or HE with primary series (96·5% [90·2-98·8%]) provided significantly greater protection than prior infection alone (80·1% [70·3-87·2%]), first booster vaccination alone (76·7% [72·5-80·4%]), or primary series alone (64·6% [54·5-73·6%]) (Figure 3, Supplementary File S15).

**Figure 3.**
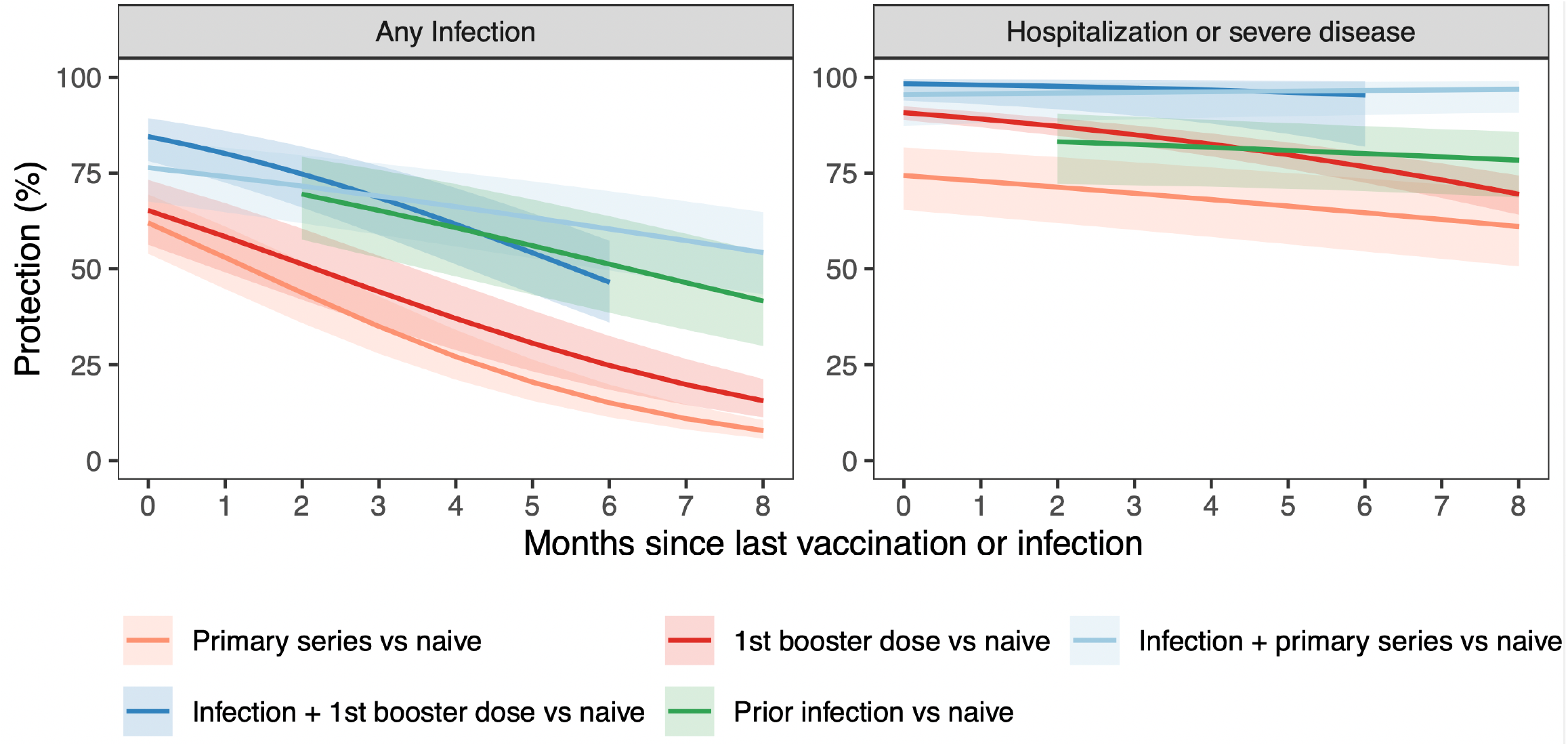
The protection against reinfection and hospitalization or severe disease conferred by the primary-series vaccine, first booster vaccine, prior infection, and hybrid immunity compared to immune naive individuals. Colour bands are 95% confidence intervals. Vaccine effectiveness data (orange and red lines) are from a previously published systematic review.^6^

Against reinfection at 6 months, there was similar protection from HE with first booster vaccination (effectiveness 46·5% [36·0-57·3%]), HE with primary series (60·4% [49·6-70·3%]), and prior infection alone (51·2% [38·6-63·7%]), with all three types of immunity conferring significantly greater protection than primary series vaccination alone (15·1% [11·3-19·8%]) or first booster vaccination alone (24·8% [18·5-32·5%]) (Figure 3, Supplementary File S15).

## Discussion

This systematic review found that prior infection and hybrid immunity conferred rapidly-waning protection against Omicron infection but sustained and high protection against COVID-19 hospitalization or severe disease caused by Omicron. Prior infection was found to provide higher protection against reinfection and more sustained protection against hospitalization or severe disease than vaccination alone. However, individuals with hybrid immunity had the highest magnitude and durability of protection against all outcomes, emphasizing the importance of providing vaccination to previously infected individuals.

A recent systematic review similarly reported rapid waning of protection against Omicron infection, but was unable to infer protection against severe disease over time and did not examine hybrid immunity.^7^ Previous studies have similarly reported that prior infection confers more durable protection than vaccination.^34^ This pattern may be explained by natural infection invoking a more diverse immune response to multiple antigenic sites on the virus relative to the immunity developed through spike-based vaccination.^7^

Protection from prior infection should not detract from the need for vaccination. First, infection-induced protection against reinfection wanes rapidly, and vaccination increases durability. Second, there are serious risks associated with infection. These include risks for hospitalization, ICU admission and mechanical ventilation, and death; they also include the risk of developing Post-COVID-19 condition (PCC).^35^ In addition, those who survive severe COVID-19 have a higher risk of cardiovascular complications, dementia, diabetes, and chronic respiratory problems.^36^ Vaccination is a safe way to avert severe disease outcomes, and reassuringly, vaccination after natural infection is not thought to be associated with an increased risk of reactogenicity or other safety concerns.^37^

On a population level, the optimal number of vaccine doses and interdose interval may differ in settings with various degrees of vaccine-induced versus infection-induced immunity. However, basing national vaccination policies on infection-induced seroprevalence rates or individual pre-vaccination screening is not practical in most settings. Programmatic vaccine roll-out should be simple; modifying the number of vaccines and intervals by infection-induced seroprevalence rates - which can only be measured in countries where inactivated vaccines were not rolled out, require well-designed serosurveys, and require use of less accurate serological tests that target anti-nucleocapsid antibodies^38,39^-will only complicate vaccine programmes and thus hamper vaccine uptake.

On the individual level, our results show that the need for and optimal timing of the primary vaccination series and booster dose may be different in an individual who has had a prior SARS-CoV-2 infection or who has experienced a breakthrough infection after initiation of the primary series when compared to a previously uninfected individual. Our findings are in line with a recent study that reported the quality and magnitude of immune responses (antibodies and B cells) to be higher if the interval between infection and booster vaccination is longer (>180 days).^40^ It may therefore be reasonable for individuals with a prior infection and full primary series vaccination to delay subsequent doses of vaccination by six months, while still maintaining high levels of protection against infection and severe disease. That said, pre-vaccination screening for past infections is currently not recommended by WHO^41^, similar to other mass vaccination programmes (e.g. measles) where past infections are not a reason to exclude persons from vaccination or delaying vaccination. In addition, there still may be benefit in providing boosters prior to periods with expected increased incidence, such as the winter seasons, to individuals whose last immunological challenge is unknown. In fact, that is a common scenario as prior infection has been largely underestimated in most settings throughout the epidemic.^3^

Our systematic review had several potential limitations. First, the patterns of declining protection shown in this study may be explained by waning in immunity; however, these results may also be in part attributable to unmeasured biases. The observational studies we included assume that individuals in the comparator and control groups are at the same risk of exposure. However, differential depletion-of-susceptibles bias may occur, where, when the vaccine is effective, the people who are infected are more likely to be unvaccinated than vaccinated, reducing the proportion of susceptible individuals in the unvaccinated group and creating the appearance of waning.^42^ Exposure could also differ between groups, as in the case of individuals who are unvaccinated because they are severely immunocompromised, and thus also have a greater risk of infection.^43–46^ The likelihood of measuring an outcome may also differ between groups — e.g., individuals with no reported prior infection who may not have had access to testing. Some of these factors were adjusted for in analysis within the individual studies (e.g., calendar time, age, comorbidities, testing frequency) and these adjustments were considered in the risk of bias assessment; however, not all studies reported these adjustments. Second, the sequence and timing between vaccination and prior infection for hybrid immunity was not considered in our analysis. Nine studies reported information on the sequence of immunological challenges, however there was insufficient data for analysis as the data were split across different types of exposures (e.g., HE with different numbers of vaccine doses) and sequence permutations. Data from studies measuring neutralizing antibodies suggest that the sequence and timing of immunological challenges may interact with the level of protection conferred^40,47^, but further studies are needed to inform policy making. Third, there was limited data for estimates of sublineage-specific protection for the distinct Omicron subvariants, such as BA.1, BA.2 or BA.5. Stratifications by key characteristics including age, sex, and severity of primary infection were also often omitted by researchers, and should be consistently reported. Fourth, in the eleven studies reporting data arising from multiple vaccines, it was unclear whether this referred to a single individual receiving different vaccine products or different sets individuals who received different sets of vaccine. This complicated our ability to generate vaccine-brand specific estimates. Fifth, we were only able to examine protection conferred by pre-Omicron variants. Future evidence synthesis will be needed on the protection conferred by Omicron infection against reinfection.

Our findings make clear the potent durability of hybrid immunity, and can help inform the timing and prioritization of vaccination programs in populations with high rates of past infection. Further follow-up of the protective effectiveness of hybrid immunity against hospitalization or severe disease, the outcome that drives most COVID-19 policy decisions, is needed to clarify how much waning of protection might occur over a longer duration. The duration of this protection will help inform the necessity and timing of future booster vaccinations. Policymakers can use these findings to project population protection from local vaccination and seroprevalence rates, helping inform their use and timing of COVID-19 vaccination as an important public health tool.

## Data Availability

Data extracted from published articles and used in our analysis will be made available upon request after approval of a study proposal. Data provided to us directly by authors of the included studies will not be shared unless permission from the original study authors is provided. Obtaining these permissions will be the responsibility of the investigators making the request for data.

## Contributors

Conceptualisation: NB, LAR, DB, IB, RA, AWS, DF, MP, LS

Data curation: NB, ZL, ZP, HI, BC, MH, RA, DF, MP, LS

Formal analysis: NB, ZL, HW, XM, LAR, RA, DF, MP, LS

Funding acquisition: NB, RA, MVK, IB

Investigation: NB, ZL, HW, XM, RH, CC, AS, MW, HI, BC, VP, DF, MP, LS

Methodology: NB, ZL, HW, XM, ZP, LAR, VP, MH, RA, AWS, DF, MP, LS

Project administration: NB, MW, DB, RA, AWS, IB, DF, MP, LS

Resources: NB, RA, MVK, IB, LS

Supervision: NB, LAR, DB, IB, MH, MVK, RA, AWS, IB, DF, MP, LS

Validation: NB, ZL, XM, RH, CC, AS, VP

Visualisation: NB, HW, XM, LAR

Writing – original draft, and writing: NB, HW, ZP, XM, RA, LS

Review & editing: all authors

All authors debated, discussed, edited, and approved the final manuscript. All authors had full access to the full data in the study and accepted responsibility to submit for publication.

## Acknowledgements

We thank the authors of the included studies that responded to our data requests and provided aggregate data for use in analysis.

