## Supplementary File for "Protective effectiveness of prior SARS-CoV-2 infection and hybrid immunity against Omicron infection and severe disease: a systematic review and meta-regression"

#### Supplementary files

S1. PRISMA checklist 1

S2. Search strategy 4

S3. Additional methodological details 9

S4. Detailed list of comparisons with corresponding effect measures and epidemiological questions 12

S5. Flowchart of Study Inclusion 13

S6. Characteristics and results of individual prior infection studies included in meta-analysis 14

S7. Characteristics and results of individual hybrid immunity studies included in meta-analysis 21

S8. Risk of bias Assessment using the ROBINS-I tool for observational studies 40

S9. Sensitivity analysis of protection conferred by prior infection or hybrid immunity over time using the WHO definition of severe disease. 44

S10. Sensitivity analysis of the protection against reinfection and severe disease conferred by the primary-series vaccine, first booster vaccine, prior infection, and hybrid immunity compared to immune naive 45

S11. Severe disease definitions from included articles 46

S12. Summary of results for studies reporting sub-group data by age 48

S13. Protective effectiveness of hybrid immunity by prior infection variant 50

S14. Protective effectiveness of hybrid immunity by vaccine type 51

S15. Six-month protection against reinfection and severe disease conferred by the primary-series vaccine, first booster vaccine, prior infection, and hybrid immunity compared to immune naïve individuals 52

References for the supplement 53

#### S1. PRISMA checklist

| Section and Topic | Item # | Checklist item | Location where item is reported |
|---|---|---|---|
| TITLE | | | |
| Title | 1 | Identify the report as a systematic review. | Page 1 |
| ABSTRACT | | | |
| Abstract | 2 | See the PRISMA 2020 for Abstracts checklist. | Page 2 |
| INTRODUCTION | | | |
| Rationale | 3 | Describe the rationale for the review in the context of existing knowledge. | Page 4, 6 |
| Objectives | 4 | Provide an explicit statement of the objective(s) or question(s) the review addresses. | Page 1, 2, 6 |
| METHODS | | | |
| Eligibility criteria | 5 | Specify the inclusion and exclusion criteria for the review and how studies were grouped for the syntheses. | Page 6, 7, Supplementary File S3 |
| Information sources | 6 | Specify all databases, registers, websites, organisations, reference lists and other sources searched or consulted to identify studies. Specify the date when each source was last searched or consulted. | Page 6, 7 |
| Search strategy | 7 | Present the full search strategies for all databases, registers and websites, including any filters and limits used. | Page 6, 7, Supplementary file 2 |
| Selection process | 8 | Specify the methods used to decide whether a study met the inclusion criteria of the review, including how many reviewers screened each record and each report retrieved, whether they worked independently, and if applicable, details of automation tools used in the process. | Page 6, 7 |
| Data collection process | 9 | Specify the methods used to collect data from reports, including how many reviewers collected data from each report, whether they worked independently, any processes for obtaining or confirming data from study investigators, and if applicable, details of automation tools used in the process. | Page 6, 7 |
| Data items | 10a | List and define all outcomes for which data were sought. Specify whether all results that were compatible with each outcome domain in each study were sought (e.g. for all measures, time points, analyses), and if not, the methods used to decide which results to collect. | Page 7, 8, Supplementary Fil S3 |
| | 10b | List and define all other variables for which data were sought (e.g. participant and intervention characteristics, funding sources). Describe any assumptions made about any missing or unclear information. | |
| Study risk of bias assessment | 11 | Specify the methods used to assess risk of bias in the included studies, including details of the tool(s) used, how many reviewers assessed each study and whether they worked independently, and if applicable, details of automation tools used in the process. | Page 8 |

| | | | |
|---|---|---|---|
| Effect measures | 12 | Specify for each outcome the effect measure(s) (e.g. risk ratio, mean difference) used in the synthesis or presentation of results. | Page 7, 8, Figure 1, Supplementary File S3 |
| Synthesis methods | 13a | Describe the processes used to decide which studies were eligible for each synthesis (e.g. tabulating the study intervention characteristics and comparing against the planned groups for each synthesis (item #5)). | Page 7, 8, Supplementary File S3 |
| | 13b | Describe any methods required to prepare the data for presentation or synthesis, such as handling of missing summary statistics, or data conversions. | Page 7, 8, Supplementary File S3 |
| | 13c | Describe any methods used to tabulate or visually display results of individual studies and syntheses. | Page 7, 8, Supplementary File S3 |
| | 13d | Describe any methods used to synthesize results and provide a rationale for the choice(s). If meta-analysis was performed, describe the model(s), method(s) to identify the presence and extent of statistical heterogeneity, and software package(s) used. | Page 7, 8, Supplementary File S3 |
| | 13e | Describe any methods used to explore possible causes of heterogeneity among study results (e.g. subgroup analysis, meta-regression). | Page 7, 8, Supplementary File S3 |
| | 13f | Describe any sensitivity analyses conducted to assess robustness of the synthesized results. | Page 7, 8, Supplementary File S3 |
| Reporting bias assessment | 14 | Describe any methods used to assess risk of bias due to missing results in a synthesis (arising from reporting biases). | Page 8 |
| Certainty assessment | 15 | Describe any methods used to assess certainty (or confidence) in the body of evidence for an outcome. | N/A |
| RESULTS | | | |
| Study selection | 16a | Describe the results of the search and selection process, from the number of records identified in the search to the number of studies included in the review, ideally using a flow diagram. | Page 9, Supplementary File S5 |
| | 16b | Cite studies that might appear to meet the inclusion criteria, but which were excluded, and explain why they were excluded. | Supplementary File S5 |
| Study characteristics | 17 | Cite each included study and present its characteristics. | Page 9, Table 1, Supplementary File S6, S7 |
| Risk of bias in studies | 18 | Present assessments of risk of bias for each included study. | Supplementary File S8 |
| Results of individual studies | 19 | For all outcomes, present, for each study: (a) summary statistics for each group (where appropriate) and (b) an effect estimate and its precision (e.g. confidence/credible interval), ideally using structured tables or plots. | Supplementary File S6, S7, S12 |
| Results of syntheses | 20a | For each synthesis, briefly summarise the characteristics and risk of bias among contributing studies. | Page 9-12, Supplementary File S8 |

| | | | |
|---|---|---|---|
| | 20b | Present results of all statistical syntheses conducted. If meta-analysis was done, present for each the summary estimate and its precision (e.g. confidence/credible interval) and measures of statistical heterogeneity. If comparing groups, describe the direction of the effect. | Page 9-12, Table 2, Table 3, Figure 2, Figure 3, Supplementary File S13, S14, S15 |
| | 20c | Present results of all investigations of possible causes of heterogeneity among study results. | Page 9-12, |
| | 20d | Present results of all sensitivity analyses conducted to assess the robustness of the synthesized results. | Page 9-12, Supplementary File S9, S10 |
| Reporting biases | 21 | Present assessments of risk of bias due to missing results (arising from reporting biases) for each synthesis assessed. | Supplementary File S8 |
| Certainty of evidence | 22 | Present assessments of certainty (or confidence) in the body of evidence for each outcome assessed. | N/A |
| DISCUSSION | | | |
| Discussion | 23a | Provide a general interpretation of the results in the context of other evidence. | Page 12 |
| | 23b | Discuss any limitations of the evidence included in the review. | Page 13, 14 |
| | 23c | Discuss any limitations of the review processes used. | Page 13, 14 |
| | 23d | Discuss implications of the results for practice, policy, and future research. | Page 13, 14 |
| OTHER INFORMATION | | | |
| Registration and protocol | 24a | Provide registration information for the review, including register name and registration number, or state that the review was not registered. | Page 6 |
| | 24b | Indicate where the review protocol can be accessed, or state that a protocol was not prepared. | Page 6 |
| | 24c | Describe and explain any amendments to information provided at registration or in the protocol. | Supplementary File S3 |
| Support | 25 | Describe sources of financial or non-financial support for the review, and the role of the funders or sponsors in the review. | Page 9 |
| Competing interests | 26 | Declare any competing interests of review authors. | Page 15 |
| Availability of data, code and other materials | 27 | Report which of the following are publicly available and where they can be found: template data collection forms; data extracted from included studies; data used for all analyses; analytic code; any other materials used in the review. | Page 15 |

#### S2. Search strategy

**Additional details of search strategies**

The search strategy comprised three search concepts: SARS-CoV-2, reinfection/protective effectiveness, and previous infection/presence of antibodies/hybrid immunity.

Comprehensive search strategies were designed by a medical librarian, for each database, incorporating both subject headings and keywords based on the individual database's unique thesaurus and available operators. The SARS-COV-2 search concept was adapted from the CADTH Covid-19 search string.[1] The search strategy was validated against known included articles from previously published reviews.

Results from the searches were exported as RIS and imported into Covidence software (www.covidence.org) for deduplication. Articles reporting identical information to previously included articles were excluded as duplicates. This rule extended to pre-print articles that were subsequently published in peer-reviewed journals. In these cases, the peer-reviewed articles were considered the definitive version.

**Search strategies used for electronic databases**

Ovid MEDLINE(R) ALL <1946 to June 02, 2022>

| # | Query | Results 2 Jun 2022 |
|---|---|---|
| 1 | COVID-19/ or exp COVID-19 Testing/ or COVID-19 Vaccines/ or SARS-CoV-2/ | 166,472 |
| 2 | (coronavirus/ or betacoronavirus/ or coronavirus infections/) and (disease outbreaks/ or epidemics/ or pandemics/) | 40,140 |
| 3 | (nCoV* or 2019nCoV or 19nCoV or COVID19* or COVID or SARS-COV-2 or SARSCOV-2 or SARS-COV2 or SARSCOV2 or SARS coronavirus 2 or Severe Acute Respiratory Syndrome Coronavirus 2 or Severe Acute Respiratory Syndrome Corona Virus 2).ti,ab,kf,nm,ot,ox,rx,px. | 252,595 |
| 4 | ((new or novel or "19" or "2019" or Wuhan or Hubei or China or Chinese) adj3 (coronavirus* or corona virus* or betacoronavirus* or CoV or HCoV)).ti,ab,kf,ot. | 70,526 |
| 5 | (longCOVID* or postCOVID* or postcoronavirus* or postSARS*).ti,ab,kf,ot. | 40 |
| 6 | ((coronavirus* or corona virus* or betacoronavirus*) adj3 (pandemic* or epidemic* or outbreak* or crisis)).ti,ab,kf,ot. | 12,534 |
| 7 | ((Wuhan or Hubei) adj5 pneumonia).ti,ab,kf,ot. | 396 |
| 8 | ((Alpha or "B.1.1.7" or Beta or "B.1.351" or Delta or "B.1.617.2" or Omicron or "B.1.1.529" or gamma or lambda) adj3 variant*).tw,kf. | 9,600 |
| 9 | 1 or 2 or 3 or 4 or 5 or 6 or 7 or 8 | 270,102 |
| 10 | Reinfection/ or Recurrence/ | 195,522 |
| 11 | (reinfect* or re-infect*).tw,kf,ot. | 13,909 |
| 12 | ((repeat* or second* or reactivat* or recurrent*) adj2 infect*).tw,kf,ot. | 32,668 |
| 13 | ((subsequent* or future) adj infect*).tw,kf,ot. | 3,601 |
| 14 | (("repeat positive" or "re-positive" or "two positive" or "2 positive") adj5 (pcr or polymerase chain reaction)).tw,kf,ot. | 231 |
| 15 | (new infection* or new SARS-CoV-2 infection*).tw,kf,ot. | 4,404 |
| 16 | ((risk adj3 infect*) or (protect* adj3 (infect* or postinfect*))).tw,kf,ot. | 83,466 |
| 17 | 10 or 11 or 12 or 13 or 14 or 15 or 16 | 321,390 |
| 18 | 9 and 17 | 8,847 |
| 19 | ((natural* or primary) adj2 (infect* or immunit*)).tw,kf,ot. | 41,996 |
| 20 | (low vaccin* or unvaccin* or un-vaccin* or "not vaccin*").tw,kf,ot. | 11,555 |
| 21 | ("anti-SARS-CoV-2 IgG" or seropositiv* or "SARS-CoV-2 antigen positiv*" or "anti-nucleocapsid IgG antibod*" or "antibody positiv*").tw,kf,ot. | 52,616 |

| | | |
|---|---|---|
| 22 | (((prior or previous* or earlier or initial or past) adj2 (infect* or disease or "covid-19" or covid19 or "SARS-CoV-2" or coronavirus or "corona virus")) or ((first or history) adj3 infection*)).tw,kf,ot. | 61,975 |
| 23 | (("covid-19" or covid19) adj2 (recovered or recovery)).tw,kf,ot. | 1,998 |
| 24 | ((recovered or convalescen*) adj1 (patient* or population* or person or persons or cases or adults or covid* or sars* or coronavirus*)).tw,kf,ot. | 18,021 |
| 25 | ((hybrid adj3 immunity) or (immunity adj3 (infect* or postinfect*))).tw,kf,ot. | 9,108 |
| 26 | 19 or 20 or 21 or 22 or 23 or 24 or 25 | 188,381 |
| 27 | 18 and 26 | 1,403 |
| 28 | limit 27 to yr="2020 - 2022" | 1,398 |

Embase <1974 to 2022 June 01>

| # | Query | Results 2 Jun 2022 |
|---|---|---|
| 1 | sars-related coronavirus/ or coronavirus disease 2019/ or asymptomatic coronavirus disease 2019/ or long covid/ or exp Severe acute respiratory syndrome coronavirus 2/ or exp SARS coronavirus/ or exp "variant of concern"/ | 239,763 |
| 2 | (coronavirinae/ or betacoronavirus/ or coronavirus infection/) and (epidemic/ or pandemic/) | 10,874 |
| 3 | (nCoV* or 2019nCoV or 19nCoV or COVID19* or COVID or SARS-COV-2 or SARSCOV-2 or SARS-COV2 or SARSCOV2 or SARS coronavirus 2 or Severe Acute Respiratory Syndrome Coronavirus 2 or Severe Acute Respiratory Syndrome Corona Virus 2).ti,ab,kf,hw,ot. | 273,124 |
| 4 | ((new or novel or "19" or "2019" or Wuhan or Hubei or China or Chinese) adj3 (coronavirus* or corona virus* or betacoronavirus* or CoV or HCoV)).ti,ab,kf,hw,ot. | 235,343 |
| 5 | (longCOVID* or postCOVID* or postcoronavirus* or postSARS*).ti,ab,kf,hw,ot. | 102 |
| 6 | ((coronavirus* or corona virus* or betacoronavirus*) adj3 (pandemic* or epidemic* or outbreak* or crisis)).ti,ab,kf,ot. | 12,307 |
| 7 | ((Wuhan or Hubei) adj5 pneumonia).ti,ab,kf,ot. | 462 |
| 8 | ((Alpha or "B.1.1.7" or Beta or "B.1.351" or Delta or "B.1.617.2" or Omicron or "B.1.1.529" or gamma or lambda) adj3 variant*).tw,kf,ot. | 10,083 |
| 9 | or/1-8 | 303,594 |
| 10 | reinfection/ or recurrent infection/ | 31,916 |
| 11 | (reinfect* or re-infect*).tw,kf,ot. | 17,000 |
| 12 | ((repeat* or second* or reactivat* or recurrent*) adj2 infect*).tw,kf,ot. | 47,197 |
| 13 | ((subsequent* or future) adj infect*).tw,kf,ot. | 4,331 |
| 14 | (("repeat positive" or "re-positive" or "two positive" or "2 positive") adj5 (pcr or polymerase chain reaction)).tw,kf,ot. | 356 |
| 15 | (new infection* or new SARS-CoV-2 infection*).tw,kf,ot. | 5,757 |
| 16 | ((risk adj3 infect*) or (protect* adj3 (infect* or postinfect*))).tw,kf,ot. | 111,501 |
| 17 | or/10-16 | 193,620 |
| 18 | 9 and 17 | 10,163 |
| 19 | primary infection/ | 4,596 |
| 20 | ((natural* or primary) adj2 (infect* or immunit*)).tw,kf,ot. | 47,657 |
| 21 | (low vaccin* or unvaccin* or un-vaccin* or "not vaccin*").tw,kf,ot. | 13,590 |
| 22 | ("anti-SARS-CoV-2 IgG" or seropositiv* or "SARS-CoV-2 antigen positiv*" or "anti-nucleocapsid IgG antibod*" or "antibody positiv*").tw,kf,ot. | 68,366 |
| 23 | (((prior or previous* or earlier or initial or past) adj2 (infect* or disease or "covid-19" or covid19 or "SARS-CoV-2" or coronavirus or "corona virus")) or ((first or history) adj3 infection*)).tw,kf,ot. | 93,696 |
| 24 | (("covid-19" or covid19) adj2 (recovered or recovery)).tw,kf,ot. | 2,295 |
| 25 | ((recovered or convalescen*) adj1 (patient* or population* or person or persons or cases or adults or covid* or sars* or coronavirus*)).tw,kf,ot. | 24,455 |

| | | |
|---|---|---|
| 26 | ((hybrid adj3 immunity) or (immunity adj3 (infect* or postinfect*))).tw,kf,ot. | 10,849 |
| 27 | or/19-26 | 250,722 |
| 28 | 18 and 27 | 1,659 |
| 29 | limit 28 to yr="2020 - 2022" | 1,642 |

Cochrane Central Register of Controlled Trials <April 2022>

| # | **Query** | **Results 2 Jun 2022** |
|---|---|---|
| 1 | COVID-19/ or exp COVID-19 Testing/ or COVID-19 Vaccines/ or SARS-CoV-2/ | 1,693 |
| 2 | (coronavirus/ or betacoronavirus/ or coronavirus infections/) and (disease outbreaks/ or epidemics/ or pandemics/) | 132 |
| 3 | (nCoV* or 2019nCoV or 19nCoV or COVID19* or COVID or SARS-COV-2 or SARSCOV-2 or SARS-COV2 or SARSCOV2 or SARS coronavirus 2 or Severe Acute Respiratory Syndrome Coronavirus 2 or Severe Acute Respiratory Syndrome Corona Virus 2).tw,kw. | 10,613 |
| 4 | ((new or novel or "19" or "2019" or Wuhan or Hubei or China or Chinese) adj3 (coronavirus* or corona virus* or betacoronavirus* or CoV or HCoV)).tw,kw. | 4,898 |
| 5 | (longCOVID* or postCOVID* or postcoronavirus* or postSARS*).tw,kw. | 6 |
| 6 | ((coronavirus* or corona virus* or betacoronavirus*) adj3 (pandemic* or epidemic* or outbreak* or crisis)).tw,kw. | 245 |
| 7 | ((Wuhan or Hubei) adj5 pneumonia).tw,kw. | 23 |
| 8 | ((Alpha or "B.1.1.7" or Beta or "B.1.351" or Delta or "B.1.617.2" or Omicron or "B.1.1.529" or gamma or lambda) adj3 variant*).tw,kw. | 116 |
| 9 | 1 or 2 or 3 or 4 or 5 or 6 or 7 or 8 | 10,921 |
| 10 | Reinfection/ or Recurrence/ | 12,532 |
| 11 | (reinfect* or re-infect*).tw,kw. | 1,308 |
| 12 | ((repeat* or second* or reactivat* or recurrent*) adj2 infect*).tw,kw. | 2,580 |
| 13 | ((subsequent* or future) adj infect*).tw,kw. | 162 |
| 14 | (("repeat positive" or "re-positive" or "two positive" or "2 positive") adj5 (pcr or polymerase chain reaction)).tw,kw. | 39 |
| 15 | (new infection* or new SARS-CoV-2 infection*).tw,kw. | 438 |
| 16 | ((risk adj3 infect*) or (protect* adj3 (infect* or postinfect*))).tw,kw. | 7,338 |
| 17 | 10 or 11 or 12 or 13 or 14 or 15 or 16 | 23,434 |
| 18 | 9 and 17 | 420 |
| 19 | ((natural* or primary) adj2 (infect* or immunit*)).tw,kw. | 1,581 |
| 20 | (low vaccin* or unvaccin* or un-vaccin* or "not vaccin*").tw,kw. | 778 |
| 21 | ("anti-SARS-CoV-2 IgG" or seropositiv* or "SARS-CoV-2 antigen positiv*" or "anti-nucleocapsid IgG antibod*" or "antibody positiv*").tw,kw. | 3,502 |
| 22 | (((prior or previous* or earlier or initial or past) adj2 (infect* or disease or "covid-19" or covid19 or "SARS-CoV-2" or coronavirus or "corona virus")) or ((first or history) adj3 infection*)).tw,kw. | 5,291 |
| 23 | (("covid-19" or covid19) adj2 (recovered or recovery)).tw,kw. | 154 |
| 24 | ((recovered or convalescen*) adj1 (patient* or population* or person or persons or cases or adults or covid* or sars* or coronavirus*)).tw,kw. | 767 |
| 25 | ((hybrid adj3 immunity) or (immunity adj3 (infect* or postinfect*))).tw,kw. | 297 |
| 26 | 19 or 20 or 21 or 22 or 23 or 24 or 25 | 11,924 |
| 27 | 18 and 26 | 56 |

Web of Science- (A&HCI , BKCI-SSH , BKCI-S , CCR-EXPANDED , ESCI , IC , CPCI-SSH , CPCI-S , SCI-EXPANDED , SSCI)

| # | Search string | Results |
|---|---|---|
| 1 | (TS=((nCoV* or 2019nCoV or 19nCoV or COVID19* or COVID or SARS-COV-2 or SARSCOV-2 or SARS-COV2 or SARSCOV2 or SARS coronavirus 2 or Severe Acute Respiratory Syndrome Coronavirus 2 or Severe Acute Respiratory Syndrome Corona Virus 2))) OR TS=((((new or novel or "19" or "2019" or Wuhan or Hubei or China or Chinese) NEAR/3 (coronavirus* or "corona virus*" or betacoronavirus* or CoV or HCoV)) OR (longCOVID* or postCOVID* or postcoronavirus* or postSARS*) OR ((coronavirus* or "corona virus*" or betacoronavirus*) NEAR/3 (pandemic* or epidemic* or outbreak* or crisis)) OR ((Wuhan or Hubei) NEAR/5 pneumonia) OR ((Alpha or "B.1.1.7" or Beta or "B.1.351" or Delta or "B.1.617.2" or Omicron or "B.1.1.529" or gamma or lambda) NEAR/3 variant*)) )<br><br>[Exact search applied] | 318,009 |
| 2 | TS=(((reinfect* or re-infect*) OR ((repeat* or second* or reactivat* or recurrent*) NEAR/2 infect*) OR ("new infection*" or "new SARS-CoV-2 infection*") OR (("repeat positive" or "re-positive" or "two positive" or "2 positive") NEAR/5 (pcr or “polymerase chain reaction”)) OR ((subsequent* or future) NEAR/1 infect*) OR (risk NEAR/3 infect*) OR (protect* NEAR/3 (infect* or postinfect*))))<br><br>[Exact search applied] | 165,361 |
| 3 | TS=((((natural* or primary) NEAR/2 (infect* or immunit*)) OR ("low vaccin*" or unvaccin* or "un-vaccin*" or "not vaccin*") OR ((prior or previous* or earlier or initial or past) NEAR/2 (infect* or disease or "covid-19" or covid19 or "SARS-CoV-2" or coronavirus or "corona virus")) or ((first or history) NEAR/4 infection*) OR "anti-SARS-CoV-2 IgG" or seropositiv* or "SARS-CoV-2 antigen positiv*" or "anti-nucleocapsid IgG antibod*" or "antibody positiv*" or (("immunity") NEAR/5 (infection* or postinfect* or hybrid)) OR (("covid-19" or covid19) NEAR/2 (recovered or recovery)) OR ((recovered or convalescen*) NEAR/1 (patient* or population* or person or persons or cases or adults or covid* or sars* or coronavirus*))) )<br><br>[Exact search applied] | 226,129 |
| 4 | (#1 AND #2 AND #3) and 2022 or 2021 or 2020 (Publication Years) | 1729 |

WHO Covid-19 Database - June 1, 2022 (1252 results)

(ti:(("reinfection" OR "reinfections" OR "re-infection" OR "re-infections" OR "Repeat infections" OR "recurrent infections" OR "repeat positive"))) OR (tw:(("reinfection" OR "reinfections" OR "re-infection" OR "re-infections" OR "Repeat infections" OR "recurrent infections" OR "repeat positive" OR "new infection" OR "risk of infection" OR "new SARS-CoV-2 infection") AND ("natural infection" OR "primary infection" OR "natural immunity" OR unvaccin* OR "un-vaccinated" OR "not vaccinated" OR "Prior infection" OR "previous infection" OR "first infection" OR "past infection" OR seropositiv* OR "antigen positive" OR "antibody positive" OR "postinfection immunity" OR "infection acquired immunity" OR "naturally acquired immunity" OR "recovered patients" OR "convalescent patients" OR "hybrid immunity" OR "hybrid protection")))

EuropePMC – June 1, 2022 (1044 results)

(ABSTRACT:(COVID19* OR COVID OR “SARS-COV-2” OR “SARSCOV-2” OR SARSCOV2 OR “Corona Virus” OR coronavirus OR postcovid OR longcovid) OR TITLE:(COVID19* OR COVID OR “SARS-COV-2” OR “SARSCOV-2” OR SARSCOV2 OR “Corona Virus” OR coronavirus OR postcovid OR longcovid) OR KW:(COVID19* OR COVID OR “SARS-COV-2” OR “SARSCOV-2” OR SARSCOV2 OR “Corona Virus” OR coronavirus OR postcovid OR longcovid)) AND (ABSTRACT:(reinfect* OR "re-infect*" OR "second* infection*" OR "repeat* infection*" OR “reactivated infection*” OR ”recurrent infection*” OR "new infection*" OR "new

SARS-CoV-2 infection*" OR “subsequent infection*” OR “future infection*” OR “risk of infection*” OR “risk of Covid* infection”) OR TITLE:(reinfect* OR "re-infect*" OR "second* infection*" OR "repeat* infection*" OR “reactivated infection*” OR ”recurrent infection*” OR "new infection*" OR "new SARS-CoV-2 infection*" OR “subsequent infection*” OR “future infection*” OR “risk of infection*” OR “risk of Covid* infection”) OR KW:(reinfect* OR "re-infect*" OR "second* infection*" OR "repeat* infection*" OR “reactivated infection*” OR ”recurrent infection*” OR "new infection*" OR "new SARS-CoV-2 infection*" OR “subsequent infection*” OR “future infection*” OR “risk of infection*” OR “risk of Covid* infection”)) AND (ABSTRACT:(“natural infection*” OR “primary infection*” OR “natural immunity” OR “naturally acquired immunity” OR unvaccin* OR "un-vaccin*" OR "not vaccin*" OR “Prior infection*” OR “previous infection*” OR “previously infected” OR "first infection*" OR “past infection*” OR seropositiv* OR "antigen positiv*" or "anti-nucleocapsid IgG antibod*" or "antibody positiv*" OR "postinfection immunity” OR “infection acquired immunity” OR “recovered patient*" OR “convalescen* patient*” OR “hybrid immunity” OR “hybrid protection”) OR TITLE:(“natural infection*” OR “primary infection*” OR “natural immunity” OR “naturally acquired immunity” OR unvaccin* OR "un-vaccin*" OR "not vaccin*" OR “Prior infection*” OR “previous infection*” OR “previously infected” OR "first infection*" OR “past infection*” OR seropositiv* OR "antigen positiv*" or "anti-nucleocapsid IgG antibod*" or "antibody positiv*" OR "postinfection immunity” OR “infection acquired immunity” OR “recovered patient*" OR “convalescen* patient*” OR “hybrid immunity” OR “hybrid protection”) OR KW:(“natural infection*” OR “primary infection*” OR “natural immunity” OR “naturally acquired immunity” OR unvaccin* OR "un-vaccin*" OR "not vaccin*" OR “Prior infection*” OR “previous infection*” OR “previously infected” OR "first infection*" OR “past infection*” OR seropositiv* OR "antigen positiv*" or "anti-nucleocapsid IgG antibod*" or "antibody positiv*" OR "postinfection immunity” OR “infection acquired immunity” OR “recovered patient*" OR “convalescen* patient*” OR “hybrid immunity” OR “hybrid protection”)) AND (SRC:"PPR")

ClinicalTrials.Gov - June 1, 2022 (12 studies)

reinfection OR reinfections OR "re-infection" OR "re-infections" OR "repeat infection" OR "second infection" OR "future infection" OR "subsequent infection" OR "recurrent infection" OR "prior infection" OR "previous infection" | Completed, Unknown status Studies | COVID-19 OR Coronavirus

#### S3. Additional methodological details

##### Detailed inclusion criteria

| | |
|---|---|
| **Population** | Humans of any age, in any geographical setting. |
| **Exposure group** | Confirmed case of SARS-CoV-2 infection with or without COVID-19 vaccination.<br><br>SARS-CoV-2 infection will be defined as a **confirmed case** according to the following criteria, adapted from WHO case definitions[2]: positive nucleic acid amplification test (NAAT) according to laboratory records or self report, positive SARS-CoV-2 antigen rapid diagnostic test (AgRDT) according to laboratory records or self report, or a positive serology test from a lab-based assay (i.e. CLIA/ELISA) or an antibody-detecting rapid diagnostic test (Ab-RDT).<br><br>Studies will be included if they report on individuals with previously confirmed infection that have documented vaccination (partial primary series, full primary series, or boosted), as defined in the randomized controlled trials for each vaccine.<br><br>**Partial vaccination** will be defined as ≥14 days after a single dose of Pfizer/BioNTech-Comirnaty (BNT162b2), AstraZeneca-Vaxzevria, Moderna-mRNA-1273, or Sinovac-CoronaVac, ≥21 days after a single dose of Sinopharm-BBIBP-CorV or Gamaleja-Sputnik-V, <7 days from the second dose for Pfizer/BioNTech-Comirnaty (BNT162b2), <14 days from the second dose for AstraZeneca-Vaxzevria, Moderna-mRNA-1273, or Sinovac-CoronaVac, <21 days from the second dose of Sinopharm-BBIBP-CorV or Gamaleja-Sputnik-V, and <14 days from the first dose of Janssen-Ad26.COV2.S.<br><br>**Primary series vaccination** will be defined as ≥7 days from the second dose for Pfizer/BioNTech-Comirnaty, ≥14 days from the first dose of Janssen-Ad26.COV2.S, ≥14 days from the second dose for AstraZeneca-Vaxzevria, Moderna-mRNA-1273, Sinovac-CoronaVac, or BBIBP-CorV Sinopharm, and ≥21 days from the second dose of Gamaleja-Sputnik-V.<br><br>**Booster vaccination one** will be defined as ≥7 days from an additional dose after primary series vaccination. |
| **Comparison group** | Five comparison groups will be eligible:<br>(1) no previous vaccinations and no previously confirmed SARS-CoV-2 infection defined using WHO criteria;<br>(2) previously confirmed SARS-CoV-2 infection defined using WHO criteria;<br>(3) partial primary series vaccination (defined above);<br>(4) full primary series vaccination (defined above);<br>(5) booster vaccination (defined above). |
| **Outcome** | SARS-CoV-2 Omicron reinfection defined as a possible, probable, or confirmed reinfection case according to the following criteria, adapted from WHO case definitions.<br><br>**Possible reinfection case** will be defined as NAAT or AgRDT SARS-CoV-2 positive case with a history of a primary SARS-CoV-2 infection diagnosed by serology, with at least 60 days between the positive serology test and the subsequent positive NAAT or AgRDT.<br><br>**Probable reinfection case** will be defined as NAAT or AgRDT SARS-CoV-2 positive case with a history of a primary SARS-CoV-2 infection diagnosed by NAAT or AgRDT, with at least 90 days between the episodes. Alternatively, genomic evidence for the second episode is available and includes lineage that was not submitted to SARS-Cov-2 genomic databases at the time of first infection.<br><br>**Confirmed reinfection case** will be defined as two PCR positive episodes supported by viral genomic data from both episodes of infection revealing different Pango lineages. If viral genomic data reveal two distinct Pango lineages, this will qualify as adequate evidence to confirm reinfection, regardless of the time elapsed between the two episodes. |

| | |
|---|---|
| | **Hospitalization** was defined as any admission to hospital with a confirmed case of SARS-CoV-2, adapted from WHO case definitions.[3]<br><br>**Severe disease** was defined using a combination of the WHO definitions of severe, critical and fatal COVID-19[4,5]:<br>- Severe COVID-19 disease was a SARS-CoV-2 infected person with "oxygen saturation of <90% on room air, and/or respiratory rate of >30 breaths/minute in adults and children >5 years old (or ≥60 breaths/minute in children <2 months old or ≥50 breaths/minute in children 2-11 months old or ≥40 breaths/minute in children 1–5 years old), and/or signs of severe respiratory distress (accessory muscle use and inability to complete full sentences, and, in children, very severe chest wall indrawing, grunting, central cyanosis, or presence of any other general danger signs)".<br>- Critical COVID-19 disease was a SARS-CoV-2 infected person with "acute respiratory distress syndrome, sepsis, septic shock, or other conditions that would normally require the provision of life sustaining therapies such as mechanical ventilation (invasive or non-invasive) or vasopressor therapy".<br>- COVID-19 death was "a death resulting from a clinically compatible illness, in a probable or confirmed COVID-19 case, unless there is a clear alternative cause of death that cannot be related to COVID-19 disease (e.g. trauma). There should be no period of complete recovery from COVID-19 between illness and death. A death due to COVID-19 may not be attributed to another disease (e.g. cancer) and should be counted independently of preexisting conditions that are suspected of triggering a severe course of COVID-19". |
| **Study design** | Test-negative case-control, case-control, cross-sectional, cohort, non-randomized controlled trials, and randomized controlled trials. |
| **Type of literature** | Published peer-reviewed research articles, preprints, and grey literature in any language. We will prioritize peer-reviewed versions of articles for inclusion and analysis in instances where pre-print versions of peer-reviewed articles are available. |

**Detailed exclusion criteria**

| | |
|---|---|
| **Population** | N/A |
| **Exposure group** | No evidence of prior confirmed case. No information on the timing, brand, or dose number for the vaccination in hybrid immunity studies. |
| **Comparison group** | N/A |
| **Outcome** | Pre-Omicron reinfection. Prior infection studies not reporting the period of time between primary infection and reinfection such that determining reinfection according to the inclusion criteria is not possible. Hybrid immunity studies not reporting the period of time between either the determination of primary infection or vaccination and reinfection. |
| **Study design** | Case reports, case series, incomplete randomized controlled trials, and review papers. |
| **Type of literature** | Media, news stories, and conference abstracts. |

**Requesting data from authors**

Authors were contacted by email to request the data if relevant data were presented in figures but not reported numerically, if data for Omicron was reported in combination with another variant of concern, or if different levels of hybrid immunity were reported in combination (i.e., partial, primary series, first booster, second booster).

**Outcome definition and comparison groups**

Estimates of VE (i.e., vaccine vs. immune naive) against Omicron variant were obtained from a recent systematic and meta-regression to compare to PE and HE estimates generated in our analysis. The VE systematic review used a similar inclusion criteria and analytic methods as our review.[6] The same raw analysis dataset used in the VE paper was obtained by reaching out to the author team, which contained the "primary series vs naive" comparison data and "1st booster dose vs naive" comparison data. We used the same log-odds meta-regression model to project the trends of VE protection waning along with HE and PE. Results are shown in figure 3.

**Analysis**

To model waning protection over time, we used log-odds meta-regression to bound protection between 0% and 100% and to translate non-linear waning on a percentage scale as linear waning on the log-odds scale. In this model, we regressed the log-odds of PE, HE, VE-infected, or PE-vaccinated on the mean time since the last immunological challenge (i.e., last vaccine dose or infection).[7] For studies reporting risk ratios or hazard ratios, we converted them to the odds ratio.[8] Our model also included a random intercept for each study, shared across all estimates of protection used from that study. Hybrid immunity analyses did not consider the order in which the immunity status was conferred (e.g., vaccination then infection or vice versa). We extracted data for each available time point and identified the mean time since the last immunological challenge. We then regressed the log-odds of PE, HE, VE-infected, or PE-vaccinated on months since the last immunological challenge.[7]

When calculating effect measures involving hybrid immunity (i.e., HE, VE-infected, PE-vaccinated) the time points for the exposure group was determined based on the most recently documented immunological challenge (i.e., vaccination or infection) prior to the period assessed for reinfection. Hybrid immunity analyses did not consider the order in which the immunity status was conferred (e.g., vaccination then infection or vice versa).

We did not differentiate between Omicron sub-variants.

**Protocol deviations**

There were two deviations from our protocol which were enacted to expand the scope of eligible data for inclusion:

1) The list of eligible types of vaccination was expanded to include any type of vaccination.
2) We did not restrict inclusion based on the accuracy of rapid diagnostic tests for confirming the index SARS-CoV-2 infection. Instead, the accuracy of the rapid diagnostic tests was considered as part of our risk of bias assessment.

#### S4. Detailed list of comparisons with corresponding effect measures and epidemiological questions

| Epidemiological and policy questions | Effect measures | Definition | Comparisons for analysis[a] |
|---|---|---|---|
| How much protection is conferred by prior infection? | PE: Protective effectiveness of prior infection | The protective effectiveness of prior infection will be 1 - odds ratio for reinfection derived by comparing previously infected (I1) unvaccinated (V0) individuals versus previously uninfected (I0) unvaccinated (V0) individuals. | Prior infection vs Naive: V0/I1 vs. V0/I0 |
| How much protection is conferred by hybrid immunity involving partial primary series, full primary series, or booster vaccination? | HE: Protective effectiveness of hybrid immunity | The protective effectiveness of hybrid immunity will be 1 - odds ratio for reinfection derived by comparing previously infected (I1) individuals who received partial primary series (VP), full primary series (VF), or booster (VB) vaccine versus previously uninfected (I0) unvaccinated (V0) individuals. | Partial primary series hybrid vs Naive: VP/I1 vs. V0/I0<br><br>Primary series hybrid vs Naive: VF/I1 vs. V0/I0<br><br>Booster one hybrid vs Naive: VB/I1 vs. V0/I0 |
| How much protection is gained by providing vaccination to people with a prior infection?<br><br>How many vaccinations does a person with prior infection need? | VE-Infection: Vaccine effectiveness among individuals with prior infection | The vaccine effectiveness among individuals with prior infection will be 1 - odds ratio for reinfection derived by comparing individuals with hybrid immunity from previous infection (I1) and partial primary series (VP), primary series (VF), or booster (VB) vaccination versus previously infected (I1) unvaccinated (V0) individuals, or previously infected (I1) individuals with a lesser number of vaccinations than the exposure group. | Partial primary series hybrid vs Infection: VP/I1 vs. V0/I1<br><br>Full Primary series hybrid vs Infection: VF/I1 vs. V0/I1<br><br>Full Primary series hybrid vs. Partial primary series hybrid: VF/I1 vs. VP/I1<br><br>Booster one hybrid vs Infection: VB/I1 vs. V0/I1<br><br>Booster one hybrid vs. Primary series hybrid: VB/I1 vs. VF/I1 |
| Is an infection equivalent to an additional vaccination amongst people who have already received one or two doses of vaccination?<br><br>Do individuals with hybrid immunity have greater protection compared to individuals with vaccination only? | PE-Vaccination: Protective effectiveness of prior infection among individuals with vaccination | The protective effectiveness of prior infection among individuals with prior vaccination will be 1 - odds ratio for reinfection derived by comparing previously infected (I1) individuals who received partial primary series (VP), full primary series (VF), or booster (VB) vaccine versus previously uninfected (I0) individuals with fewer vaccinations or an equivalent number of vaccinations. When the exposure and comparisons groups have an equivalent number of vaccinations the difference between groups is prior infection. When the comparison group has a greater number of vaccinations than the exposure group this measure helps to determine whether prior infection serves as a substitute for additional vaccination. | Partial primary series hybrid vs. Partial primary series vaccine: VP/I1 vs. VP/I0<br><br>Partial primary series hybrid vs. Full primary series vaccine: VP/I1 vs. VF/I0<br><br>Partial primary series hybrid vs. Booster one vaccine: VP/I1 vs. VB/I0<br><br>Full primary series hybrid vs. Full primary series vaccine: VF/I1 vs. VF/I0<br><br>Full primary series hybrid vs. Booster one vaccine: VF/I1 vs. VB/I0<br><br>Booster one hybrid vs. Booster one vaccine: VB/I1 vs. VB/I0 |

[a]Prior infection abbreviations: previously infected - I1; never infected - I0. Vaccination abbreviations: unvaccinated - V0; partial primary series vaccination - VP; full primary series vaccination - VF; first booster vaccination - VB. The notation when combining immunity status for prior infection and vaccination was a slash (/), which did not imply an order of the events (e.g., full primary series vaccination and prior infection was denoted as VF/I1).

### S5. Flowchart of Study Inclusion

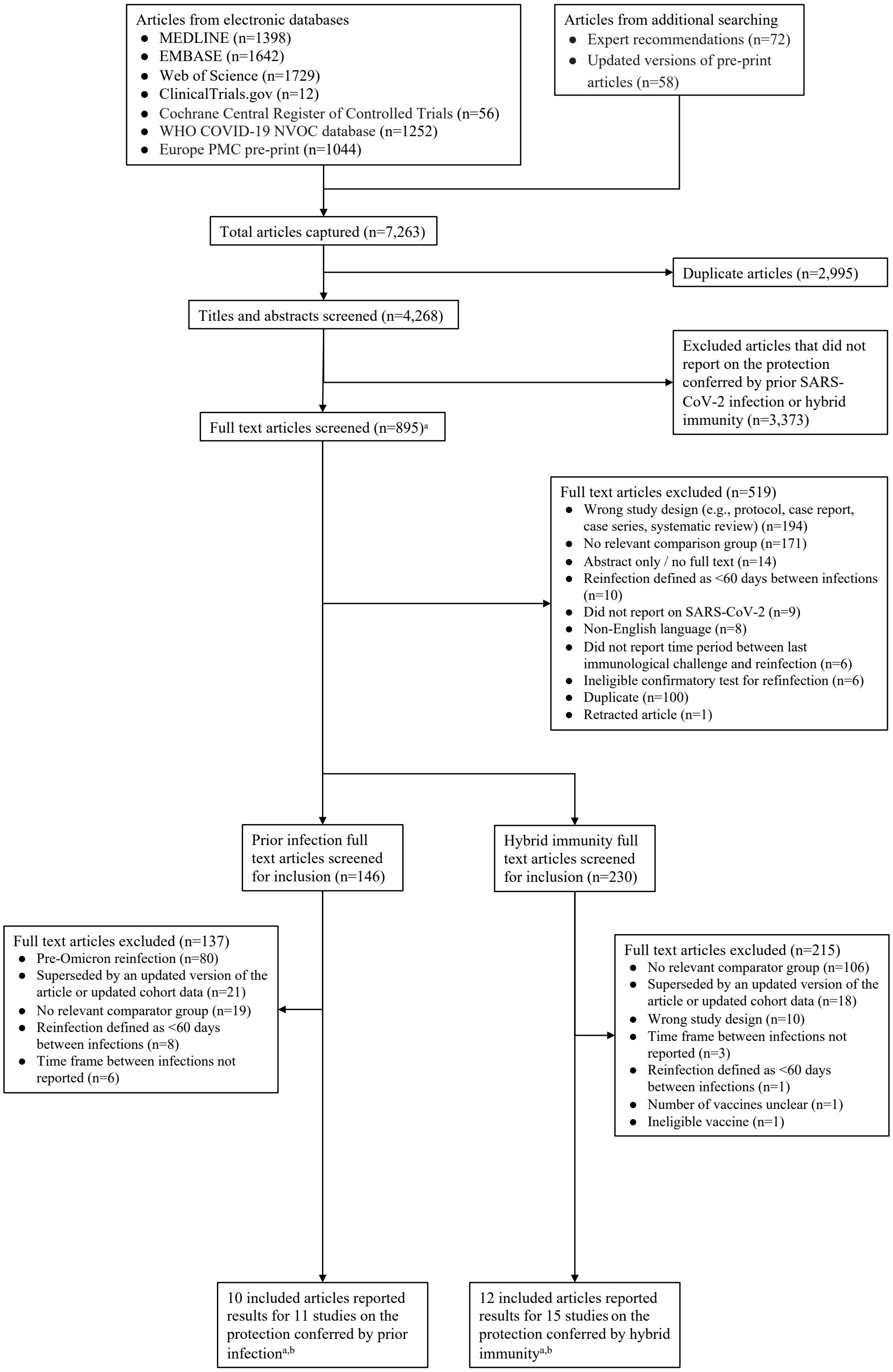

[a]There were two rounds of full text screening. Studies not meeting general inclusion criteria were excluded in the first round. In the second round, articles were screened with criteria specific to the types of exposures and comparators being reported (i.e., prior infection and hybrid immunity). [b]Ten articles reported 1 study, 6 articles reported 2 studies (1 prior infection and 1 hybrid immunity), 1 article reported 3 studies (1 prior infection and 2 hybrid immunity), 1 article reported 4 studies (2 prior infection and 2 hybrid immunity). [c]Seven articles reported data for prior infection and hybrid immunity.

#### S6. Characteristics and results of individual prior infection studies included in meta-analysis

| Author (Country) | Study Design (Variables controlled for in the prior infection protection estimates) | Severity of Infection | Reinfection variant | Prior Infected Variant | Days since Prior Infection/ Vaccination Completion (Median [Range]) | Age Range | Protection[a] [95% CI] | Risk of Bias |
|---|---|---|---|---|---|---|---|---|
| Altarawneh 1 (Qatar)[9] | Test-negative case-control (Matched cohorts. Adjusted for presence of co-morbidities) | | | | | | | |
| | | Any Infection | Omicron | Mixed variant | 180 [90-269] | All ages | 64·0% [54·7-71·4%] | Serious |
| | | Any Infection | Omicron | Mixed variant | 360 [270-449] | All ages | 47·2% [37·5-55·4%] | Serious |
| | | Any Infection | Omicron | Mixed variant | 450 | All ages | 59·6% [50·7-67%] | Serious |
| | | Hospitalization and severe disease | Omicron | Mixed variant | 314 | All ages | 87·8% [47·5-97·1%] | Moderate |
| Altarawneh 2 (Qatar)[10] | Test-negative case-control (Matched cohorts· Adjusted for sex, age group, nationality, and calendar week of PCR test) | Any Infection | Omicron | Mixed variant | 324 | All ages | 44·9% [39·2-50·1%] | Moderate |
| | | Any Infection | Omicron (BA·1) | Mixed variant | 324 | All ages | 50·2% [38·1-59·9%] | Moderate |
| | | Any Infection | Omicron (BA·2) | Mixed variant | 324 | All ages | 46·1% [39·5-51·9%] | Moderate |
| | | Hospitalization and severe disease | Omicron | Mixed variant | 324 | All ages | 85·5% [49·1-95·9%] | Moderate |
| | | Hospitalization and severe disease | Omicron (BA·1) | Mixed variant | 324 | All ages | 92·7% [1·0-99·6%] | Moderate |
| | | Hospitalization and severe disease | Omicron (BA·2) | Mixed variant | 324 | All ages | 73·4% [0·2-92·9%] | Moderate |
| Andeweg (Netherlands)[11] | Test-negative case-control (Adjusted for age, sex, health region, and testing date) | Any Infection | Omicron (BA·1) | Index-Delta | 104 [90-119] | 18-100 | 57% [52-62%] | Serious |
| | | Any Infection | Omicron (BA·1) | Index-Delta | 134 [120-149] | 18-100 | 55% [46-62%] | Serious |

| | | | | | | |
|---|---|---|---|---|---|---|
| Any Infection | Omicron (BA·1) | Index-Delta | 164 [150-179] | 18-100 | 53% [43-60%] | Serious |
| Any Infection | Omicron (BA.1) | Index-Delta | 194 [180-209] | 18-100 | 52% [45-58%] | Serious |
| Any Infection | Omicron (BA.1) | Index-Delta | 210 | 18-100 | 34% [31-38%] | Serious |
| Any Infection | Omicron (BA.1) | Index-Delta | 104 [90-119] | 18-100 | 55% [48-60%] | Serious |
| Any Infection | Omicron (BA.1) | Index-Delta | 134 [120-149] | 18-100 | 51% [41-60%] | Serious |
| Any Infection | Omicron (BA.1) | Index-Delta | 164 [150-179] | 18-100 | 54% [44-62%] | Serious |
| Any Infection | Omicron (BA.1) | Index-Delta | 194 [180-209] | 18-100 | 53% [45-59%] | Serious |
| Any Infection | Omicron (BA.1) | Index-Delta | 210 | 18-100 | 37% [33-40%] | Serious |
| Any Infection | Omicron (BA.2) | Index-Delta | 104 [90-119] | 18-100 | 50% [43-56%] | Serious |
| Any Infection | Omicron (BA.2) | Index-Delta | 134 [120-149] | 18-100 | 57% [49-64%] | Serious |
| Any Infection | Omicron (BA.2) | Index-Delta | 164 [150-179] | 18-100 | 50% [36-61%] | Serious |
| Any Infection | Omicron (BA.2) | Index-Delta | 194 [180-209] | 18-100 | 53% [42-62%] | Serious |
| Any Infection | Omicron (BA.2) | Index-Delta | 210 | 18-100 | 38% [34-43%] | Serious |
| Any Infection | Omicron (BA.2) | Index-Delta | 104 [90-119] | 18-100 | 50% [43-56%] | Serious |
| Any Infection | Omicron (BA.2) | Index-Delta | 134 [120-149] | 18-100 | 58% [50-66%] | Serious |
| Any Infection | Omicron (BA.2) | Index-Delta | 164 [150-179] | 18-100 | 55% [41-65%] | Serious |
| Any Infection | Omicron (BA.2) | Index-Delta | 194 [180-209] | 18-100 | 52% [40-62%] | Serious |

| | | | | | | | | |
|---|---|---|---|---|---|---|---|---|
| | | Any Infection | Omicron (BA.2) | Index-Delta | 210 | 18-100 | 40% [35-44%] | Serious |
| | | Any Infection | Omicron (BA.1) | Index-Delta | 104 [90-119] | 18-100 | 42% [1-68%] | Serious |
| | | Any Infection | Omicron (BA.1) | Index-Delta | 134 [120-149] | 18-100 | 31% [1-54%] | Serious |
| | | Any Infection | Omicron (BA.1) | Index-Delta | 164 [150-179] | 18-100 | 28% [8-43%] | Serious |
| | | Any Infection | Omicron (BA.1) | Index-Delta | 194 [180-209] | 18-100 | 12% [1-43%] | Serious |
| | | Any Infection | Omicron (BA.1) | Index-Delta | 210 | 18-100 | 6% [1-16%] | Serious |
| | | Any Infection | Omicron (BA.1) | Index-Delta | 104 [90-119] | 18-100 | 52% [3-77%] | Serious |
| | | Any Infection | Omicron (BA.1) | Index-Delta | 134 [120-149] | 18-100 | 24% [1-51%] | Serious |
| | | Any Infection | Omicron (BA.1) | Index-Delta | 164 [150-179] | 18-100 | 22% [1-40%] | Serious |
| | | Any Infection | Omicron (BA.1) | Index-Delta | 194 [180-209] | 18-100 | 1% [1-35%] | Serious |
| | | Any Infection | Omicron (BA.1) | Index-Delta | 210 | 18-100 | 5% [1-16%] | Serious |
| Carazo (Canada)[12] | Test-negative case-control (Adjusted for age, sex, testing-indication and epi-week but not comorbidity) | Any Infection | Omicron | Alpha (B.1.1.7),Beta,Gamma,Delta | 134 [90-179] | 12-100 | 67% [57-74%] | Serious |
| | | Any Infection | Omicron | Alpha (B.1.1.7),Beta,Gamma,Delta | 455 [180-730] | 12-100 | 37% [29-43%] | Serious |
| | | Any Infection | Omicron | Alpha (B.1.1.7),Beta,Gamma,Delta | 136 [90-182] | 12-100 | 66% [57-73%] | Serious |
| | | Any Infection | Omicron | Alpha (B.1.1.7),Beta,Gamma,Delta | 228 [183-274] | 12-100 | 49% [32-61%] | Serious |
| | | Any Infection | Omicron | Alpha (B.1.1.7),Beta,Gamma,Delta | 320 [275-364] | 12-100 | 35% [21-47%] | Serious |

| | | | | | | | | |
|---|---|---|---|---|---|---|---|---|
| | | Any Infection | Omicron | Alpha (B.1.1.7),Beta,Gamma,Delta | 456 [365-547] | 12-100 | 29% [17-38%] | Serious |
| | | Any Infection | Omicron | Alpha (B.1.1.7),Beta,Gamma,Delta | 639 [548-730] | 12-100 | 27% [8-42%] | Serious |
| | | Any Infection | Omicron | Alpha (B.1.1.7),Beta,Gamma,Delta | 134 [90-179] | 12-100 | 49% [8-72%] | Serious |
| | | Any Infection | Omicron | Alpha (B.1.1.7),Beta,Gamma,Delta | 455 [180-730] | 12-100 | 1% [1-20%] | Serious |
| | | Hospitalization and severe disease | Omicron | Alpha (B.1.1.7),Beta,Gamma,Delta | 134 [90-179] | 12-100 | 81% [52-92%] | Moderate |
| | | Hospitalization and severe disease | Omicron | Alpha (B.1.1.7),Beta,Gamma,Delta | 455 [180-730] | 12-100 | 62% [36-77%] | Moderate |
| Cerqueira-Silva (Brazil)[13] | Test-negative case-control (Matched cohorts. Adjusted for matched design, comorbidities, pregnancy, race, days elapsed between tests, hospital admission, and age) | Any Infection | Omicron | Alpha (B.1.1.7),Delta (B.1.617.2),Gamma | 134 [90-179] | 18-100 | 52·8% [48·3-56·8%] | Moderate |
| | | Any Infection | Omicron | Alpha (B.1.1.7),Delta (B.1.617.2),Gamma | 272 [180-365] | 18-100 | 32·7% [30·2-35·2%] | Moderate |
| | | Any Infection | Omicron | Alpha (B.1.1.7),Delta (B.1.617.2),Gamma | 365 | 18-100 | 14·7% [10·8-18·5%] | Moderate |
| | | Hospitalization and severe disease | Omicron | Alpha (B.1.1.7),Delta (B.1.617.2),Gamma | 134 [90-179] | 18-100 | 84·5% [73·1-91·1%] | Serious |
| | | Hospitalization and severe disease | Omicron | Alpha (B.1.1.7),Delta (B.1.617.2),Gamma | 272 [180-365] | 18-100 | 89·5% [86-92·2%] | Serious |
| | | Hospitalization and severe disease | Omicron | Alpha (B.1.1.7),Delta (B.1.617.2),Gamma | 365 | 18-100 | 80·3% [74·4-84·8%] | Serious |
| Chin (USA)[14] | Test-negative case-control (Matched cohorts. Adjusted for age group, and gender) | Any Infection | Omicron | Delta (B.1.617.2) | 150 | 18-100 | 54·2% [41·4-66·2%] | Moderate |
| | | Any Infection | Omicron | Delta (B.1.617.2) | 154 | 18-100 | 60·9% [58·5-67·8%] | Moderate |

| | | | | | | | | |
|---|---|---|---|---|---|---|---|---|
| Lind (USA)[15] | Test-negative case-control (Adjusted for date of test, age (continuous), sex, race/ethnicity, comorbidity score, clinical encounters, insurance group, and regional social vulnerability) | Any Infection | Omicron | Mixed variant | 328 | 5-100 | 25·3% [16·1-33·5%] | Moderate |
| Michlmayr (Denmark)[16] | Cohort (Adjusted age, sex, comorbidity, region of affiliation and staying at hospital, vaccination status, and time since vaccination) | Any Infection | Omicron | Mixed variant | 134 [90-179] | 2-100 | 41·7% [40·8-42·6%] | Serious |
| | | Any Infection | Omicron | Mixed variant | 224 [180-269] | 2-100 | 18·8% [17·1-20·3%] | Serious |
| | | Any Infection | Omicron | Mixed variant | 314 [270-359] | 2-100 | 18·5% [16·9-20·1%] | Serious |
| | | Any Infection | Omicron | Mixed variant | 360 | 2-100 | 13·7% [12·3-14·8%] | Serious |
| | | Any Infection | Omicron | Mixed variant | 134 [90-179] | 2-100 | 48·1% [46·3-49·9%] | Serious |
| | | Any Infection | Omicron | Mixed variant | 224 [180-269] | 2-100 | 25·3% [22·5-27·9%] | Serious |
| | | Any Infection | Omicron | Mixed variant | 314 [270-359] | 2-100 | 25·6% [22·9-28·2%] | Serious |
| | | Any Infection | Omicron | Mixed variant | 360 | 2-100 | 20·6% [18·1-22·9%] | Serious |
| | | Hospitalization and severe disease | Omicron | Mixed variant | 90 | 2-100 | 69·8% [51·5-81·2%] | Serious |
| Nyberg (UK)[17] | Retrospective cohort (Adjusted for age, sex, and index of multiple deprivation) | Hospitalization and severe disease | Omicron | Index-Delta | 180 | 18-100 | 45% [37-52%] | Serious |
| | | Hospitalization and severe disease | Omicron | Index-Delta | 180 | 18-100 | 32% [26-38%] | Serious |
| | | Hospitalization and severe disease | Omicron | Index-Delta | 180 | 18-100 | 28% [22-33%] | Serious |

| | | | | | | | | |
|---|---|---|---|---|---|---|---|---|
| | | Hospitalization and severe disease | Omicron | Index-Delta | 180 | 18-100 | 82% [43-94%] | Serious |
| | | Hospitalization and severe disease | Omicron | Index-Delta | 180 | 18-100 | 82% [76-89%] | Serious |
| | | Hospitalization and severe disease | Omicron | Index-Delta | 180 | 18-100 | 93% [88-97%] | Serious |
| | | Hospitalization and severe disease | Omicron | Index-Delta | 180 | 18-100 | 94% [90-99%] | Serious |
| | | Hospitalization and severe disease | Omicron | Index-Delta | 180 | 18-100 | 34% [22-54%] | Serious |
| | | Hospitalization and severe disease | Omicron | Index-Delta | 180 | 18-100 | 69% [55-88%] | Serious |
| | | Hospitalization and severe disease | Omicron | Index-Delta | 180 | 18-100 | 84% [72-99%] | Serious |
| | | Hospitalization and severe disease | Omicron | Index-Delta | 180 | 18-100 | 86% [74-100%] | Serious |
| | | Hospitalization and severe disease | Omicron | Index-Delta | 180 | 18-100 | 48% [21-100%] | Serious |
| | | Hospitalization and severe disease | Omicron | Index-Delta | 180 | 18-100 | 54% [47-62%] | Serious |
| | | Hospitalization and severe disease | Omicron | Index-Delta | 180 | 18-100 | 37% [29-44%] | Serious |
| | | Hospitalization and severe disease | Omicron | Index-Delta | 180 | 18-100 | 32% [25-40%] | Serious |
| | | Hospitalization and severe disease | Omicron | Index-Delta | 180 | 18-100 | 90% [78-100%] | Serious |
| Šmíd (Czechia)[18] | Cross-sectional (Adjusted for age group, sex, and calendar time) | Any Infection | Omicron | Delta (B.1.617.2) | 120 [60-180] | All ages | 66·3% [66·3-67·3%] | Moderate |
| | | Any Infection | Omicron | Delta (B.1.617.2) | 120 [60-180] | All ages | 69% [68-69%] | Moderate |
| | | Any Infection | Omicron | Wild-type,Alpha (B.1.1.7) | 241 [181-301] | All ages | 48% [46-50%] | Moderate |
| | | Any Infection | Omicron | Wild-type,Alpha (B.1.1.7) | 362 [302-422] | All ages | 34% [33-35%] | Moderate |

| | | | | | | |
|---|---|---|---|---|---|---|
| Any Infection | Omicron | Wild-type,Alpha (B.1.1.7) | 423 | All ages | 17% [15-18%] | Moderate |
| Any Infection | Omicron | Wild-type,Alpha (B.1.1.7) | 302 [181-423] | All ages | 12·1% [10·3-13·1%] | Moderate |
| Hospitalization and severe disease | Omicron | Delta (B.1.617.2) | 120 [60-180] | All ages | 73% [55-84%] | Moderate |
| Hospitalization and severe disease | Omicron | Wild-type,Alpha (B.1.1.7) | 302 [181-423] | All ages | 66% [54-75%] | Moderate |
| Hospitalization and severe disease | Omicron | Delta (B.1.617.2) | 120 [60-180] | All ages | 80·8% [40-93·9%] | Moderate |
| Hospitalization and severe disease | Omicron | Wild-type,Alpha (B.1.1.7) | 302 [181-423] | All ages | 87·6% [72·2-94·5%] | Moderate |
| Hospitalization and severe disease | Omicron | Delta (B.1.617.2) | 120 [60-180] | All ages | 82·8% [1-97·6%] | Moderate |
| Hospitalization and severe disease | Omicron | Wild-type,Alpha (B.1.1.7) | 302 [181-423] | All ages | 65·7% [15-86·1%] | Moderate |

n=10 studies.[a] The exposure and comparator groups are "Infection vs naive" for all PE estimates.

#### S7. Characteristics and results of individual hybrid immunity studies included in meta-analysis

| Author (Country) | Study Design (Variables controlled for in the hybrid immunity protection estimates) | Measure | Exposure vs Comparator | Vaccine | Severity of Infection | Reinfection Variant | Prior Infected Variant | Days since Prior Infection/ Vaccination Completion (Median [Range]) | Age Range | Protection (Est. [95% CI]) | Risk of Bias |
|---|---|---|---|---|---|---|---|---|---|---|---|
| Altarawneh 2 (Qatar)[10] | Test-negative case-control (Matched cohorts. Adjusted for sex, age group, nationality, and calendar week of PCR test) | HE | Infection + 1st booster vaccine vs naive | Pfizer/BioNTech-Comirnaty | Any Infection | Omicron (BA.1) | Mixed variant | 42 | 0-100 | 74·4% [63·4-82·2%] | Moderate |
| | | HE | Infection + 1st booster vaccine vs naive | Pfizer/BioNTech-Comirnaty | Any Infection | Omicronn (BA.2) | Mixed variant | 43 | 0-100 | 77·3% [72·4-81·4%] | Moderate |
| | | HE | Infection + 1st booster vaccine vs naive | Pfizer/BioNTech-Comirnaty | Hospitalization and severe disease | Omicronn (BA.2) | Mixed variant | 43 | 0-100 | 97·5% [57·6-99·9%] | Moderate |
| | | HE | Infection + full primary series vaccine vs naive | Pfizer/BioNTech-Comirnaty | Any Infection | Omicronn (BA.1) | Mixed variant | 268 | 0-100 | 51·7% [43·5-58·7%] | Moderate |
| | | HE | Infection + full primary series vaccine vs naive | Pfizer/BioNTech-Comirnaty | Any Infection | Omicronn (BA.2) | Mixed variant | 270 | 0-100 | 55·1% [50·9-58·9%] | Moderate |
| | | HE | Infection + full primary series vaccine vs naive | Pfizer/BioNTech-Comirnaty | Hospitalization and severe disease | Omicronn (BA.2) | Mixed variant | 270 | 0-100 | 97·8% [82·6-99·7%] | Moderate |
| | | HE | Infection + full primary series vaccine vs naive | Pfizer/BioNTech-Comirnaty | Hospitalization and severe disease | Omicronn (BA.1) | Mixed variant | 268 | 0-100 | 96·2% [37·7-99·8%] | Moderate |
| Andeweg (Netherlands)[11] | Test-negative case-control (Adjusted for age, sex, health region, and testing date) | HE | Infection + 1st booster vaccine vs naive | Pfizer/BioNTech-Comirnaty,Moderna-mRNA-1273,AstraZeneca-Vaxzevria,Janssen-Ad26.COV2.S | Any Infection | Omicron (BA.1) | Mixed variant | 104 [90-119] | 18-100 | 66% [46-79%] | Serious |
| | | HE | Infection + 1st booster vaccine vs naive | Pfizer/BioNTech-Comirnaty,Moderna-mRNA-1273,AstraZeneca-Vaxzevria,Janssen-Ad26.COV2.S | Any Infection | Omicron (BA.1) | Mixed variant | 104 [90-119] | 18-100 | 70% [50-82%] | Serious |
| | | HE | Infection + 1st booster vaccine vs naive | Pfizer/BioNTech-Comirnaty,Moderna-mRNA-1273,AstraZeneca-Vaxzevria,Janssen-Ad26.COV2.S | Any Infection | Omicron (BA.2) | Mixed variant | 104 [90-119] | 18-100 | 75% [67-81%] | Serious |
| | | HE | Infection + 1st booster vaccine vs naive | Pfizer/BioNTech-Comirnaty,Moderna-mRNA-1273,AstraZeneca-Vaxzevria,Janssen-Ad26.COV2.S | Any Infection | Omicron (BA.2) | Mixed variant | 104 [90-119] | 18-100 | 74% [66-80%] | Serious |

| | | | | | | | | | |
|---|---|---|---|---|---|---|---|---|---|
| HE | Infection + full primary series vaccine vs naive | Pfizer/BioNTech-Comirnaty,Moderna-mRNA-1273,AstraZeneca-Vaxzevria,Janssen-Ad26.COV2.S | Any Infection | Omicron (BA.1) | Delta (B.1.617.2) | 134 [120-149] | 18-100 | 85% [82-87%] | Serious |
| HE | Infection + full primary series vaccine vs naive | Pfizer/BioNTech-Comirnaty,Moderna-mRNA-1273,AstraZeneca-Vaxzevria,Janssen-Ad26.COV2.S | Any Infection | Omicron (BA.1) | Delta (B.1.617.2) | 164 [150-179] | 18-100 | 81% [75-86%] | Serious |
| HE | Infection + full primary series vaccine vs naive | Pfizer/BioNTech-Comirnaty,Moderna-mRNA-1273,AstraZeneca-Vaxzevria,Janssen-Ad26.COV2.S | Any Infection | Omicron (BA.1) | Delta (B.1.617.2) | 104 [90-119] | 18-100 | 66% [53-75%] | Serious |
| HE | Infection + full primary series vaccine vs naive | Pfizer/BioNTech-Comirnaty,Moderna-mRNA-1273,AstraZeneca-Vaxzevria,Janssen-Ad26.COV2.S | Any Infection | Omicron (BA.1) | Delta (B.1.617.2) | 134 [120-149] | 18-100 | 67% [52-77%] | Serious |
| HE | Infection + full primary series vaccine vs naive | Pfizer/BioNTech-Comirnaty,Moderna-mRNA-1273,AstraZeneca-Vaxzevria,Janssen-Ad26.COV2.S | Any Infection | Omicron (BA.1) | Delta (B.1.617.2) | 164 [150-179] | 18-100 | 68% [50-79%] | Serious |
| HE | Infection + full primary series vaccine vs naive | Pfizer/BioNTech-Comirnaty,Moderna-mRNA-1273,AstraZeneca-Vaxzevria,Janssen-Ad26.COV2.S | Any Infection | Omicron (BA.1) | Delta (B.1.617.2) | 194 [180-209] | 18-100 | 82% [79-85%] | Serious |
| HE | Infection + full primary series vaccine vs naive | Pfizer/BioNTech-Comirnaty,Moderna-mRNA-1273,AstraZeneca-Vaxzevria,Janssen-Ad26.COV2.S | Any Infection | Omicron (BA.1) | Delta (B.1.617.2) | 210 | 18-100 | 82% [75-88%] | Serious |
| HE | Infection + full primary series vaccine vs naive | Pfizer/BioNTech-Comirnaty,Moderna-mRNA-1273,AstraZeneca-Vaxzevria,Janssen-Ad26.COV2.S | Any Infection | Omicron (BA.1) | Delta (B.1.617.2) | 104 [90-119] | 18-100 | 66% [52-76%] | Serious |
| HE | Infection + full primary series vaccine vs naive | Pfizer/BioNTech-Comirnaty,Moderna-mRNA-1273,AstraZeneca-Vaxzevria,Janssen-Ad26.COV2.S | Any Infection | Omicron (BA.1) | Delta (B.1.617.2) | 134 [120-149] | 18-100 | 69% [52-80%] | Serious |
| HE | Infection + full primary series vaccine vs naive | Pfizer/BioNTech-Comirnaty,Moderna-mRNA-1273,AstraZeneca-Vaxzevria,Janssen-Ad26.COV2.S | Any Infection | Omicron (BA.1) | Delta (B.1.617.2) | 164 [150-179] | 18-100 | 72% [54-83%] | Serious |
| HE | Infection + full primary series vaccine vs naive | Pfizer/BioNTech-Comirnaty,Moderna-mRNA-1273,AstraZeneca-Vaxzevria,Janssen-Ad26.COV2.S | Any Infection | Omicron (BA.2) | Delta (B.1.617.2) | 194 [180-209] | 18-100 | 84% [81-86%] | Serious |
| HE | Infection + full primary series vaccine vs naive | Pfizer/BioNTech-Comirnaty,Moderna-mRNA-1273,AstraZeneca-Vaxzevria,Janssen-Ad26.COV2.S | Any Infection | Omicron (BA.2) | Delta (B.1.617.2) | 210 | 18-100 | 85% [80-89%] | Serious |
| HE | Infection + full primary series vaccine vs naive | Pfizer/BioNTech-Comirnaty,Moderna-mRNA-1273,AstraZeneca-Vaxzevria,Janssen-Ad26.COV2.S | Any Infection | Omicron (BA.2) | Delta (B.1.617.2) | 104 [90-119] | 18-100 | 77% [66-85%] | Serious |

| | | | | | | | | | |
|---|---|---|---|---|---|---|---|---|---|
| HE | Infection + full primary series vaccine vs naive | Pfizer/BioNTech-Comirnaty,Moderna-mRNA-1273,AstraZeneca-Vaxzevria,Janssen-Ad26.COV2.S | Any Infection | Omicron (BA.2) | Delta (B.1.617.2) | 194 [180-209] | 18-100 | 64% [43-77%] | Serious |
| HE | Infection + full primary series vaccine vs naive | Pfizer/BioNTech-Comirnaty,Moderna-mRNA-1273,AstraZeneca-Vaxzevria,Janssen-Ad26.COV2.S | Any Infection | Omicron (BA.2) | Delta (B.1.617.2) | 210 | 18-100 | 48% [26-64%] | Serious |
| HE | Infection + full primary series vaccine vs naive | Pfizer/BioNTech-Comirnaty,Moderna-mRNA-1273,AstraZeneca-Vaxzevria,Janssen-Ad26.COV2.S | Any Infection | Omicron (BA.2) | Delta (B.1.617.2) | 194 [180-209] | 18-100 | 84% [81-87%] | Serious |
| HE | Infection + full primary series vaccine vs naive | Pfizer/BioNTech-Comirnaty,Moderna-mRNA-1273,AstraZeneca-Vaxzevria,Janssen-Ad26.COV2.S | Any Infection | Omicron (BA.2) | Delta (B.1.617.2) | 210 | 18-100 | 85% [80-89%] | Serious |
| HE | Infection + full primary series vaccine vs naive | Pfizer/BioNTech-Comirnaty,Moderna-mRNA-1273,AstraZeneca-Vaxzevria,Janssen-Ad26.COV2.S | Any Infection | Omicron (BA.2) | Delta (B.1.617.2) | 104 [90-119] | 18-100 | 76% [63-84%] | Serious |
| HE | Infection + full primary series vaccine vs naive | Pfizer/BioNTech-Comirnaty,Moderna-mRNA-1273,AstraZeneca-Vaxzevria,Janssen-Ad26.COV2.S | Any Infection | Omicron (BA.2) | Delta (B.1.617.2) | 134 [120-149] | 18-100 | 65% [43-78%] | Serious |
| HE | Infection + full primary series vaccine vs naive | Pfizer/BioNTech-Comirnaty,Moderna-mRNA-1273,AstraZeneca-Vaxzevria,Janssen-Ad26.COV2.S | Any Infection | Omicron (BA.2) | Delta (B.1.617.2) | 164 [150-179] | 18-100 | 51% [28-67%] | Serious |
| HE | Infection + full primary series vaccine vs naive | Pfizer/BioNTech-Comirnaty,Moderna-mRNA-1273,AstraZeneca-Vaxzevria,Janssen-Ad26.COV2.S | Any Infection | Omicron (BA.1) | Wild-type,Alpha (B.1.1.7),Delta (B.1.617.2) | 194 [180-209] | 18-100 | 68% [57-75%] | Serious |
| HE | Infection + full primary series vaccine vs naive | Pfizer/BioNTech-Comirnaty,Moderna-mRNA-1273,AstraZeneca-Vaxzevria,Janssen-Ad26.COV2.S | Any Infection | Omicron (BA.1) | Wild-type,Alpha (B.1.1.7),Delta (B.1.617.2) | 210 | 18-100 | 60% [44-71%] | Serious |
| HE | Infection + full primary series vaccine vs naive | Pfizer/BioNTech-Comirnaty,Moderna-mRNA-1273,AstraZeneca-Vaxzevria,Janssen-Ad26.COV2.S | Any Infection | Omicron (BA.1) | Wild-type,Alpha (B.1.1.7),Delta (B.1.617.2) | 104 [90-119] | 18-100 | 61% [47-71%] | Serious |
| HE | Infection + full primary series vaccine vs naive | Pfizer/BioNTech-Comirnaty,Moderna-mRNA-1273,AstraZeneca-Vaxzevria,Janssen-Ad26.COV2.S | Any Infection | Omicron (BA.1) | Wild-type,Alpha (B.1.1.7),Delta (B.1.617.2) | 134 [120-149] | 18-100 | 56% [46-64%] | Serious |
| HE | Infection + full primary series vaccine vs naive | Pfizer/BioNTech-Comirnaty,Moderna-mRNA-1273,AstraZeneca-Vaxzevria,Janssen-Ad26.COV2.S | Any Infection | Omicron (BA.1) | Wild-type,Alpha (B.1.1.7),Delta (B.1.617.2) | 164 [150-179] | 18-100 | 63% [55-70%] | Serious |
| HE | Infection + full primary series vaccine vs naive | Pfizer/BioNTech-Comirnaty,Moderna-mRNA-1273,AstraZeneca-Vaxzevria,Janssen-Ad26.COV2.S | Any Infection | Omicron (BA.1) | Wild-type,Alpha (B.1.1.7),Delta (B.1.617.2) | 194 [180-209] | 18-100 | 68% [56-77%] | Serious |

| | | | | | | | | | |
|---|---|---|---|---|---|---|---|---|---|
| HE | Infection + full primary series vaccine vs naive | Pfizer/BioNTech-Comirnaty,Moderna-mRNA-1273,AstraZeneca-Vaxzevria,Janssen-Ad26.COV2.S | Any Infection | Omicron (BA.1) | Wild-type,Alpha (B.1.1.7),Delta (B.1.617.2) | 134 [120-149] | 18-100 | 53% [32-67%] | Serious |
| HE | Infection + full primary series vaccine vs naive | Pfizer/BioNTech-Comirnaty,Moderna-mRNA-1273,AstraZeneca-Vaxzevria,Janssen-Ad26.COV2.S | Any Infection | Omicron (BA.1) | Wild-type,Alpha (B.1.1.7),Delta (B.1.617.2) | 164 [150-179] | 18-100 | 52% [32-66%] | Serious |
| HE | Infection + full primary series vaccine vs naive | Pfizer/BioNTech-Comirnaty,Moderna-mRNA-1273,AstraZeneca-Vaxzevria,Janssen-Ad26.COV2.S | Any Infection | Omicron (BA.1) | Wild-type,Alpha (B.1.1.7),Delta (B.1.617.2) | 104 [90-119] | 18-100 | 52% [39-62%] | Serious |
| HE | Infection + full primary series vaccine vs naive | Pfizer/BioNTech-Comirnaty,Moderna-mRNA-1273,AstraZeneca-Vaxzevria,Janssen-Ad26.COV2.S | Any Infection | Omicron (BA.1) | Wild-type,Alpha (B.1.1.7),Delta (B.1.617.2) | 134 [120-149] | 18-100 | 63% [54-70%] | Serious |
| HE | Infection + full primary series vaccine vs naive | Pfizer/BioNTech-Comirnaty,Moderna-mRNA-1273,AstraZeneca-Vaxzevria,Janssen-Ad26.COV2.S | Any Infection | Omicron (BA.2) | Wild-type,Alpha (B.1.1.7),Delta (B.1.617.2) | 164 [150-179] | 18-100 | 76% [64-84%] | Serious |
| HE | Infection + full primary series vaccine vs naive | Pfizer/BioNTech-Comirnaty,Moderna-mRNA-1273,AstraZeneca-Vaxzevria,Janssen-Ad26.COV2.S | Any Infection | Omicron (BA.2) | Wild-type,Alpha (B.1.1.7),Delta (B.1.617.2) | 194 [180-209] | 18-100 | 69% [52-80%] | Serious |
| HE | Infection + full primary series vaccine vs naive | Pfizer/BioNTech-Comirnaty,Moderna-mRNA-1273,AstraZeneca-Vaxzevria,Janssen-Ad26.COV2.S | Any Infection | Omicron (BA.2) | Wild-type,Alpha (B.1.1.7),Delta (B.1.617.2) | 210 | 18-100 | 69% [50-81%] | Serious |
| HE | Infection + full primary series vaccine vs naive | Pfizer/BioNTech-Comirnaty,Moderna-mRNA-1273,AstraZeneca-Vaxzevria,Janssen-Ad26.COV2.S | Any Infection | Omicron (BA.2) | Wild-type,Alpha (B.1.1.7),Delta (B.1.617.2) | 104 [90-119] | 18-100 | 62% [46-73%] | Serious |
| HE | Infection + full primary series vaccine vs naive | Pfizer/BioNTech-Comirnaty,Moderna-mRNA-1273,AstraZeneca-Vaxzevria,Janssen-Ad26.COV2.S | Any Infection | Omicron (BA.2) | Wild-type,Alpha (B.1.1.7),Delta (B.1.617.2) | 134 [120-149] | 18-100 | 67% [58-74%] | Serious |
| HE | Infection + full primary series vaccine vs naive | Pfizer/BioNTech-Comirnaty,Moderna-mRNA-1273,AstraZeneca-Vaxzevria,Janssen-Ad26.COV2.S | Any Infection | Omicron (BA.2) | Wild-type,Alpha (B.1.1.7),Delta (B.1.617.2) | 164 [150-179] | 18-100 | 77% [64-85%] | Serious |
| HE | Infection + full primary series vaccine vs naive | Pfizer/BioNTech-Comirnaty,Moderna-mRNA-1273,AstraZeneca-Vaxzevria,Janssen-Ad26.COV2.S | Any Infection | Omicron (BA.2) | Wild-type,Alpha (B.1.1.7),Delta (B.1.617.2) | 194 [180-209] | 18-100 | 67% [47-79%] | Serious |
| HE | Infection + full primary series vaccine vs naive | Pfizer/BioNTech-Comirnaty,Moderna-mRNA-1273,AstraZeneca-Vaxzevria,Janssen-Ad26.COV2.S | Any Infection | Omicron (BA.2) | Wild-type,Alpha (B.1.1.7),Delta (B.1.617.2) | 210 | 18-100 | 65% [40-79%] | Serious |
| HE | Infection + full primary series vaccine vs naive | Pfizer/BioNTech-Comirnaty,Moderna-mRNA-1273,AstraZeneca-Vaxzevria,Janssen-Ad26.COV2.S | Any Infection | Omicron (BA.2) | Wild-type,Alpha (B.1.1.7),Delta (B.1.617.2) | 104 [90-119] | 18-100 | 62% [44-74%] | Serious |

| | | | | | | | | | |
|---|---|---|---|---|---|---|---|---|---|
| HE | Infection + full primary series vaccine vs naive | Pfizer/BioNTech-Comirnaty,Moderna-mRNA-1273,AstraZeneca-Vaxzevria,Janssen-Ad26.COV2.S | Any Infection | Omicron (BA.2) | Wild-type,Alpha (B.1.1.7),Delta (B.1.617.2) | 134 [120-149] | 18-100 | 67% [57-74%] | Serious |
| HE | Infection + full primary series vaccine vs naive | Pfizer/BioNTech-Comirnaty,Moderna-mRNA-1273,AstraZeneca-Vaxzevria,Janssen-Ad26.COV2.S | Any Infection | Omicron (BA.1) | Delta (B.1.617.2) | 164 [150-179] | 18-100 | 42% [23-57%] | Serious |
| HE | Infection + full primary series vaccine vs naive | Pfizer/BioNTech-Comirnaty,Moderna-mRNA-1273,AstraZeneca-Vaxzevria,Janssen-Ad26.COV2.S | Any Infection | Omicron (BA.1) | Delta (B.1.617.2) | 194 [180-209] | 18-100 | 35% [9-53%] | Serious |
| HE | Infection + full primary series vaccine vs naive | Pfizer/BioNTech-Comirnaty,Moderna-mRNA-1273,AstraZeneca-Vaxzevria,Janssen-Ad26.COV2.S | Any Infection | Omicron (BA.1) | Delta (B.1.617.2) | 210 | 18-100 | 39% [20-53%] | Serious |
| HE | Infection + full primary series vaccine vs naive | Pfizer/BioNTech-Comirnaty,Moderna-mRNA-1273,AstraZeneca-Vaxzevria,Janssen-Ad26.COV2.S | Any Infection | Omicron (BA.1) | Delta (B.1.617.2) | 104 [90-119] | 18-100 | 36% [1-63%] | Serious |
| HE | Infection + full primary series vaccine vs naive | Pfizer/BioNTech-Comirnaty,Moderna-mRNA-1273,AstraZeneca-Vaxzevria,Janssen-Ad26.COV2.S | Any Infection | Omicron (BA.1) | Delta (B.1.617.2) | 134 [120-149] | 18-100 | 79% [1-97%] | Serious |
| HE | Infection + full primary series vaccine vs naive | Pfizer/BioNTech-Comirnaty,Moderna-mRNA-1273,AstraZeneca-Vaxzevria,Janssen-Ad26.COV2.S | Any Infection | Omicron (BA.1) | Delta (B.1.617.2) | 164 [150-179] | 18-100 | 41% [14-60%] | Serious |
| HE | Infection + full primary series vaccine vs naive | Pfizer/BioNTech-Comirnaty,Moderna-mRNA-1273,AstraZeneca-Vaxzevria,Janssen-Ad26.COV2.S | Any Infection | Omicron (BA.1) | Delta (B.1.617.2) | 194 [180-209] | 18-100 | 35% [1-59%] | Serious |
| HE | Infection + full primary series vaccine vs naive | Pfizer/BioNTech-Comirnaty,Moderna-mRNA-1273,AstraZeneca-Vaxzevria,Janssen-Ad26.COV2.S | Any Infection | Omicron (BA.1) | Delta (B.1.617.2) | 210 | 18-100 | 28% [1-48%] | Serious |
| HE | Infection + full primary series vaccine vs naive | Pfizer/BioNTech-Comirnaty,Moderna-mRNA-1273,AstraZeneca-Vaxzevria,Janssen-Ad26.COV2.S | Any Infection | Omicron (BA.1) | Delta (B.1.617.2) | 164 [150-179] | 18-100 | 1% [1-44%] | Serious |
| HE | Infection + full primary series vaccine vs naive | Pfizer/BioNTech-Comirnaty,Moderna-mRNA-1273,AstraZeneca-Vaxzevria,Janssen-Ad26.COV2.S | Any Infection | Omicron (BA.1) | Delta (B.1.617.2) | 210 | 18-100 | 51% [1-94%] | Serious |
| HE | Infection + full primary series vaccine vs naive | Pfizer/BioNTech-Comirnaty,Moderna-mRNA-1273,AstraZeneca-Vaxzevria,Janssen-Ad26.COV2.S | Any Infection | Omicron (BA.1) | Wild-type,Alpha (B.1.1.7),Delta (B.1.617.2) | 104 [90-119] | 18-100 | 55% [28-72%] | Serious |
| HE | Infection + full primary series vaccine vs naive | Pfizer/BioNTech-Comirnaty,Moderna-mRNA-1273,AstraZeneca-Vaxzevria,Janssen-Ad26.COV2.S | Any Infection | Omicron (BA.1) | Wild-type,Alpha (B.1.1.7),Delta (B.1.617.2) | 134 [120-149] | 18-100 | 48% [30-62%] | Serious |

| | | | | | | | | | | | |
|---|---|---|---|---|---|---|---|---|---|---|---|
| | | HE | Infection + full primary series vaccine vs naive | Pfizer/BioNTech-Comirnaty,Moderna-mRNA-1273,AstraZeneca-Vaxzevria,Janssen-Ad26.COV2.S | Any Infection | Omicron (BA.1) | Wild-type,Alpha (B.1.1.7),Delta (B.1.617.2) | 134 [120-149] | 18-100 | 40% [25-52%] | Serious |
| | | HE | Infection + full primary series vaccine vs naive | Pfizer/BioNTech-Comirnaty,Moderna-mRNA-1273,AstraZeneca-Vaxzevria,Janssen-Ad26.COV2.S | Any Infection | Omicron (BA.1) | Wild-type,Alpha (B.1.1.7),Delta (B.1.617.2) | 194 [180-209] | 18-100 | 59% [43-70%] | Serious |
| | | HE | Infection + full primary series vaccine vs naive | Pfizer/BioNTech-Comirnaty,Moderna-mRNA-1273,AstraZeneca-Vaxzevria,Janssen-Ad26.COV2.S | Any Infection | Omicron (BA.1) | Wild-type,Alpha (B.1.1.7),Delta (B.1.617.2) | 210 | 18-100 | 50% [26-66%] | Serious |
| | | HE | Infection + full primary series vaccine vs naive | Pfizer/BioNTech-Comirnaty,Moderna-mRNA-1273,AstraZeneca-Vaxzevria,Janssen-Ad26.COV2.S | Any Infection | Omicron (BA.1) | Wild-type,Alpha (B.1.1.7),Delta (B.1.617.2) | 104 [90-119] | 18-100 | 46% [8-69%] | Serious |
| | | HE | Infection + full primary series vaccine vs naive | Pfizer/BioNTech-Comirnaty,Moderna-mRNA-1273,AstraZeneca-Vaxzevria,Janssen-Ad26.COV2.S | Any Infection | Omicron (BA.1) | Wild-type,Alpha (B.1.1.7),Delta (B.1.617.2) | 104 [90-119] | 18-100 | 50% [27-66%] | Serious |
| | | HE | Infection + full primary series vaccine vs naive | Pfizer/BioNTech-Comirnaty,Moderna-mRNA-1273,AstraZeneca-Vaxzevria,Janssen-Ad26.COV2.S | Any Infection | Omicron (BA.1) | Wild-type,Alpha (B.1.1.7),Delta (B.1.617.2) | 164 [150-179] | 18-100 | 44% [26-58%] | Serious |
| | | HE | Infection + full primary series vaccine vs naive | Pfizer/BioNTech-Comirnaty,Moderna-mRNA-1273,AstraZeneca-Vaxzevria,Janssen-Ad26.COV2.S | Any Infection | Omicron (BA.1) | Wild-type,Alpha (B.1.1.7),Delta (B.1.617.2) | 194 [180-209] | 18-100 | 58% [40-71%] | Serious |
| | | HE | Infection + full primary series vaccine vs naive | Pfizer/BioNTech-Comirnaty,Moderna-mRNA-1273,AstraZeneca-Vaxzevria,Janssen-Ad26.COV2.S | Any Infection | Omicron (BA.1) | Wild-type,Alpha (B.1.1.7),Delta (B.1.617.2) | 210 | 18-100 | 47% [18-66%] | Serious |
| Björk (Sweden)[19] | Traditional case-control (Age and sex matched case and control cohorts. Adjusted for for comorbidities) | HE | Infection + full primary series vaccine vs naive | Pfizer/BioNTech-Comirnaty,Moderna-mRNA-1273,AstraZeneca-Vaxzevria | Hospitalization and severe disease | Omicron (BA.1) | Index-Delta | 94 [7-180] | 0-100 | 91% [57-98%] | Serious |
| | | HE | Infection + full primary series vaccine vs naive | Pfizer/BioNTech-Comirnaty,Moderna-mRNA-1273,AstraZeneca-Vaxzevria | Hospitalization and severe disease | Omicron (BA.2) | Index-Delta | 94 [7-180] | 0-100 | 53% [1-82%] | Serious |
| | | HE | Infection + full primary series vaccine vs naive | Pfizer/BioNTech-Comirnaty,Moderna-mRNA-1273,AstraZeneca-Vaxzevria | Hospitalization and severe disease | Omicron | Index-Delta | 94 [7-180] | 0-100 | 92% [59-98%] | Serious |
| Bruel (France)[20] | Cohort (No adjustment) | PEV | Infection + 1st booster vaccine vs 1st booster vaccine | Pfizer/BioNTech-Comirnaty,Moderna-mRNA-1273, | Any Infection | Omicron | Mixed variant | 90 | 72-101 | 89% [29-98%] | Serious |
| Carazo (Canada)[12] | Test-negative case-control (Adjusted for | HE | Infection + 1st booster vaccine vs naive | Pfizer/BioNTech-Comirnaty,Moderna-mRNA-1273 | Any Infection | Omicron | Alpha (B.1.1.7),Beta,Gamma,Delta | 7 | 12-100 | 83% [81-84%] | Serious |

| | | | | | | | | | | |
|---|---|---|---|---|---|---|---|---|---|---|
| age, sex, testing-indication and epi-week but not comorbidity) | HE | Infection + 1st booster vaccine vs naive | Pfizer/BioNTech-Comirnaty,Moderna-mRNA-1273 | Any Infection | Omicron | Alpha (B.1.1.7),Beta,Gamma,Delta | 120 [60-179] | 12-100 | 80% [76-84%] | Serious |
| | HE | Infection + 1st booster vaccine vs naive | Pfizer/BioNTech-Comirnaty,Moderna-mRNA-1273 | Any Infection | Omicron | Alpha (B.1.1.7),Beta,Gamma,Delta | 34 [7-60] | 12-100 | 83% [81-84%] | Serious |
| | HE | Infection + 1st booster vaccine vs naive | Pfizer/BioNTech-Comirnaty,Moderna-mRNA-1273 | Hospitalization and severe disease | Omicron | Alpha (B.1.1.7),Beta,Gamma,Delta | 7 | 12-100 | 99% [84·5-99·9%] | Moderate |
| | HE | Infection + 1st booster vaccine vs naive | Pfizer/BioNTech-Comirnaty,Moderna-mRNA-1273 | Hospitalization and severe disease | Omicron | Alpha (B.1.1.7),Beta,Gamma,Delta | 7 | 12-100 | 99·9% [95·3-98·9%] | Moderate |
| | HE | Infection + 1st booster vaccine vs naive | Pfizer/BioNTech-Comirnaty,Moderna-mRNA-1273 | Hospitalization and severe disease | Omicron | Alpha (B.1.1.7),Beta,Gamma,Delta | 7 | 12-100 | 97% [94-99%] | Moderate |
| | HE | Infection + full primary series vaccine vs naive | Pfizer/BioNTech-Comirnaty,Moderna-mRNA-1273 | Any Infection | Omicron | Alpha (B.1.1.7),Beta,Gamma,Delta | 7 | 12-100 | 68% [67-70%] | Serious |
| | HE | Infection + full primary series vaccine vs naive | Pfizer/BioNTech-Comirnaty,Moderna-mRNA-1273 | Any Infection | Omicron | Alpha (B.1.1.7),Beta,Gamma,Delta | 34 [7-60] | 12-100 | 82% [80-84%] | Serious |
| | HE | Infection + full primary series vaccine vs naive | Pfizer/BioNTech-Comirnaty,Moderna-mRNA-1273 | Any Infection | Omicron | Alpha (B.1.1.7),Beta,Gamma,Delta | 120 [60-179] | 12-100 | 67% [65-68%] | Serious |
| | HE | Infection + full primary series vaccine vs naive | Pfizer/BioNTech-Comirnaty,Moderna-mRNA-1273 | Any Infection | Omicron | Alpha (B.1.1.7),Beta,Gamma,Delta | 224 [180-269] | 12-100 | 63% [60-65%] | Serious |
| | HE | Infection + full primary series vaccine vs naive | Pfizer/BioNTech-Comirnaty,Moderna-mRNA-1273 | Any Infection | Omicron | Alpha (B.1.1.7),Beta,Gamma,Delta | 314 [270-359] | 12-100 | 62% [42-75%] | Serious |
| | HE | Infection + full primary series vaccine vs naive | Pfizer/BioNTech-Comirnaty,Moderna-mRNA-1273 | Hospitalization and severe disease | Omicron | Alpha (B.1.1.7),Beta,Gamma,Delta | 7 | 12-100 | 99·6% [93·3-100%] | Moderate |
| | HE | Infection + full primary series vaccine vs naive | Pfizer/BioNTech-Comirnaty,Moderna-mRNA-1273 | Hospitalization and severe disease | Omicron | Alpha (B.1.1.7),Beta,Gamma,Delta | 97 | 12-100 | 86·3% [1-99·1%] | Moderate |
| | HE | Infection + full primary series vaccine vs naive | Pfizer/BioNTech-Comirnaty,Moderna-mRNA-1273 | Hospitalization and severe disease | Omicron | Alpha (B.1.1.7),Beta,Gamma,Delta | 97 | 12-100 | 99·8% [67·3-99·4%] | Moderate |

| | | | | | | | | | | | |
|---|---|---|---|---|---|---|---|---|---|---|---|
| | | HE | Infection + full primary series vaccine vs naive | Pfizer/BioNTech-Comirnaty,Moderna-mRNA-1273 | Hospitalization and severe disease | Omicron | Alpha (B.1.1.7),Beta,Gamma,Delta | 7 | 12-100 | 94% [91-96%] | Moderate |
| | | HE | Infection + full primary series vaccine vs naive | Pfizer/BioNTech-Comirnaty,Moderna-mRNA-1273 | Hospitalization and severe disease | Omicron | Alpha (B.1.1.7),Beta,Gamma,Delta | 7 | 12-100 | 99·9% [95·1-97·9%] | Moderate |
| | | VEI | Infection + 1st booster vaccine vs infection | Pfizer/BioNTech-Comirnaty,Moderna-mRNA-1273 | Any Infection | Omicron | Alpha (B.1.1.7),Beta,Gamma,Delta | 7 | 12-100 | 69·4% [66·1-72·3%] | Serious |
| | | VEI | Infection + 1st booster vaccine vs infection | Pfizer/BioNTech-Comirnaty,Moderna-mRNA-1273 | Hospitalization and severe disease | Omicron | Alpha (B.1.1.7),Beta,Gamma,Delta | 7 | 12-100 | 82·3% [55·5-92·9%] | Moderate |
| | | VEI | Infection + full primary series vaccine vs infection | Pfizer/BioNTech-Comirnaty,Moderna-mRNA-1273 | Any Infection | Omicron | Alpha (B.1.1.7),Beta,Gamma,Delta | 7 | 12-100 | 29·1% [22·8-34·9%] | Serious |
| | | VEI | Infection + full primary series vaccine vs infection | Pfizer/BioNTech-Comirnaty,Moderna-mRNA-1273 | Hospitalization and severe disease | Omicron | Alpha (B.1.1.7),Beta,Gamma,Delta | 7 | 12-100 | 74·6% [49·8-87·2%] | Moderate |
| | | VEI | Infection + full primary series vaccine vs infection | Pfizer/BioNTech-Comirnaty,Moderna-mRNA-1273 | Hospitalization and severe disease | Omicron | Alpha (B.1.1.7),Beta,Gamma,Delta | 97 | 12-100 | 63·9% [1-95·3%] | Moderate |
| | | VEI[a] | Infection + 1st booster vaccine vs infection + full primary series vaccine | Pfizer/BioNTech-Comirnaty,Moderna-mRNA-1273 | Any Infection | Omicron | Alpha (B.1.1.7),Beta,Gamma,Delta | 7 | 12-100 | 56·8% [53·6-59·7%] | Serious |
| | | VEI[a] | Infection + 1st booster vaccine vs infection + full primary series vaccine | Pfizer/BioNTech-Comirnaty,Moderna-mRNA-1273 | Hospitalization and severe disease | Omicron | Alpha (B.1.1.7),Beta,Gamma,Delta | 7 | 12-100 | 30·1% [1-70%] | Moderate |
| Cerqueira-Silva (Brazil)[13] | Test-negative case-control (Matched cohorts. Adjusted for matched design, comorbidities, pregnancy, race, days elapsed between tests, hospital admission, and age) | HE | Infection + 1st booster vaccine vs naive | AstraZeneca-Vaxzevria | Any Infection | Omicron | Mixed variant | 70 | 18-100 | 50·8% [48·9-52·7%] | Moderate |
| | | HE | Infection + 1st booster vaccine vs naive | AstraZeneca-Vaxzevria | Any Infection | Omicron | Mixed variant | 6 [0-13] | 18-100 | 68·8% [67·5-70%] | Moderate |
| | | HE | Infection + 1st booster vaccine vs naive | AstraZeneca-Vaxzevria | Any Infection | Omicron | Mixed variant | 42 [14-69] | 18-100 | 72·1% [71·4-72·8%] | Moderate |
| | | HE | Infection + 1st booster vaccine vs naive | CoronaVac | Any Infection | Omicron | Mixed variant | 6 [0-13] | 18-100 | 66·9% [64·7-69%] | Moderate |
| | | HE | Infection + 1st booster vaccine vs naive | CoronaVac | Any Infection | Omicron | Mixed variant | 42 [14-69] | 18-100 | 73·4% [72·4-74·3%] | Moderate |

| | | | | | | | | | |
|---|---|---|---|---|---|---|---|---|---|
| HE | Infection + 1st booster vaccine vs naive | CoronaVac | Any Infection | Omicron | Mixed variant | 70 | 18-100 | 54·6% [53·7-55·5%] | Moderate |
| HE | Infection + 1st booster vaccine vs naive | Janssen-Ad26.COV2.S | Any Infection | Omicron | Mixed variant | 6 [0-13] | 18-100 | 52·4% [46·9-57·4%] | Moderate |
| HE | Infection + 1st booster vaccine vs naive | Janssen-Ad26.COV2.S | Any Infection | Omicron | Mixed variant | 42 [14-69] | 18-100 | 44·8% [42·4-47·2%] | Moderate |
| HE | Infection + 1st booster vaccine vs naive | Pfizer/BioNTech-Comirnaty | Any Infection | Omicron | Mixed variant | 6 [0-13] | 18-100 | 68% [65·8-70·2%] | Moderate |
| HE | Infection + 1st booster vaccine vs naive | Pfizer/BioNTech-Comirnaty | Any Infection | Omicron | Mixed variant | 42 [14-69] | 18-100 | 68·2% [66·4-69·9%] | Moderate |
| HE | Infection + 1st booster vaccine vs naive | Pfizer/BioNTech-Comirnaty | Any Infection | Omicron | Mixed variant | 70 | 18-100 | 58·2% [45·4-68·1%] | Moderate |
| HE | Infection + 1st booster vaccine vs naive | AstraZeneca-Vaxzevria | Hospitalization and severe disease | Omicron | Mixed variant | 42 [14-69] | 18-100 | 98·1% [97·7-98·5%] | Moderate |
| HE | Infection + 1st booster vaccine vs naive | AstraZeneca-Vaxzevria | Hospitalization and severe disease | Omicron | Mixed variant | 70 | 18-100 | 97·2% [96·2-98%] | Moderate |
| HE | Infection + 1st booster vaccine vs naive | AstraZeneca-Vaxzevria | Hospitalization and severe disease | Omicron | Mixed variant | 6 [0-13] | 18-100 | 97·6% [96·2-98·5%] | Moderate |
| HE | Infection + 1st booster vaccine vs naive | CoronaVac | Hospitalization and severe disease | Omicron | Mixed variant | 42 [14-69] | 18-100 | 96·9% [96-97·6%] | Moderate |
| HE | Infection + 1st booster vaccine vs naive | CoronaVac | Hospitalization and severe disease | Omicron | Mixed variant | 70 | 18-100 | 96·7% [96·2-97·1%] | Moderate |
| HE | Infection + 1st booster vaccine vs naive | CoronaVac | Hospitalization and severe disease | Omicron | Mixed variant | 6 [0-13] | 18-100 | 95% [89·9-97·6%] | Moderate |
| HE | Infection + 1st booster vaccine vs naive | Janssen-Ad26.COV2.S | Hospitalization and severe disease | Omicron | Mixed variant | 6 [0-13] | 18-100 | 93·3% [72·9-98·3%] | Moderate |
| HE | Infection + 1st booster vaccine vs naive | Janssen-Ad26.COV2.S | Hospitalization and severe disease | Omicron | Mixed variant | 42 [14-69] | 18-100 | 97·8% [94-99·2%] | Moderate |

| | | | | | | | | | |
|---|---|---|---|---|---|---|---|---|---|
| HE | Infection + 1st booster vaccine vs naive | Pfizer/BioNTech-Comirnaty | Hospitalization and severe disease | Omicron | Mixed variant | 6 [0-13] | 18-100 | 99·6% [93·3-100%] | Moderate |
| HE | Infection + 1st booster vaccine vs naive | Pfizer/BioNTech-Comirnaty | Hospitalization and severe disease | Omicron | Mixed variant | 42 [14-69] | 18-100 | 96·8% [94·1-98·2%] | Moderate |
| HE | Infection + full primary series vaccine vs naive | AstraZeneca-Vaxzevria | Any Infection | Omicron | Mixed variant | 6 [0-13] | 18-100 | 59·7% [49·7-67·7%] | Moderate |
| HE | Infection + full primary series vaccine vs naive | AstraZeneca-Vaxzevria | Any Infection | Omicron | Mixed variant | 42 [14-69] | 18-100 | 45·5% [42·6-48·3%] | Moderate |
| HE | Infection + full primary series vaccine vs naive | AstraZeneca-Vaxzevria | Any Infection | Omicron | Mixed variant | 104 [70-139] | 18-100 | 38·8% [37·7-39·8%] | Moderate |
| HE | Infection + full primary series vaccine vs naive | AstraZeneca-Vaxzevria | Any Infection | Omicron | Mixed variant | 140 | 18-100 | 40·7% [39·6-41·7%] | Moderate |
| HE | Infection + full primary series vaccine vs naive | CoronaVac | Any Infection | Omicron | Mixed variant | 6 [0-13] | 18-100 | 53·3% [43·3-61·4%] | Moderate |
| HE | Infection + full primary series vaccine vs naive | CoronaVac | Any Infection | Omicron | Mixed variant | 42 [14-69] | 18-100 | 46% [42·6-49·2%] | Moderate |
| HE | Infection + full primary series vaccine vs naive | CoronaVac | Any Infection | Omicron | Mixed variant | 104 [70-139] | 18-100 | 31% [29·4-32·5%] | Moderate |
| HE | Infection + full primary series vaccine vs naive | CoronaVac | Any Infection | Omicron | Mixed variant | 140 | 18-100 | 36·2% [34·9-37·4%] | Moderate |
| HE | Infection + full primary series vaccine vs naive | Janssen-Ad26.COV2.S | Any Infection | Omicron | Mixed variant | 14 | 18-100 | 39·7% [37·5-41·8%] | Moderate |
| HE | Infection + full primary series vaccine vs naive | Pfizer/BioNTech-Comirnaty | Any Infection | Omicron | Mixed variant | 42 [14-69] | 18-100 | 63·6% [62·5-64·7%] | Moderate |
| HE | Infection + full primary series vaccine vs naive | Pfizer/BioNTech-Comirnaty | Any Infection | Omicron | Mixed variant | 6 [0-13] | 18-100 | 71·1% [66·8-74·8%] | Moderate |
| HE | Infection + full primary series vaccine vs naive | Pfizer/BioNTech-Comirnaty | Any Infection | Omicron | Mixed variant | 104 [70-139] | 18-100 | 50·2% [49·4-50·9%] | Moderate |

| | | | | | | | | | |
|---|---|---|---|---|---|---|---|---|---|
| HE | Infection + full primary series vaccine vs naive | Pfizer/BioNTech-Comirnaty | Any Infection | Omicron | Mixed variant | 140 | 18-100 | 45·7% [43·7-47·7%] | Moderate |
| HE | Infection + full primary series vaccine vs naive | AstraZeneca-Vaxzevria | Hospitalization and severe disease | Omicron | Mixed variant | 42 [14-69] | 18-100 | 89·9% [81·9-94·3%] | Moderate |
| HE | Infection + full primary series vaccine vs naive | AstraZeneca-Vaxzevria | Hospitalization and severe disease | Omicron | Mixed variant | 104 [70-139] | 18-100 | 93·9% [92·8-94·9%] | Moderate |
| HE | Infection + full primary series vaccine vs naive | AstraZeneca-Vaxzevria | Hospitalization and severe disease | Omicron | Mixed variant | 140 | 18-100 | 94·5% [93·8-95·1%] | Moderate |
| HE | Infection + full primary series vaccine vs naive | CoronaVac | Hospitalization and severe disease | Omicron | Mixed variant | 104 [70-139] | 18-100 | 88·7% [85·4-91·3%] | Moderate |
| HE | Infection + full primary series vaccine vs naive | CoronaVac | Hospitalization and severe disease | Omicron | Mixed variant | 42 [14-69] | 18-100 | 88·4% [77·9-93·9%] | Moderate |
| HE | Infection + full primary series vaccine vs naive | CoronaVac | Hospitalization and severe disease | Omicron | Mixed variant | 140 | 18-100 | 90·7% [89·5-91·8%] | Moderate |
| HE | Infection + full primary series vaccine vs naive | Janssen-Ad26.COV2.S | Hospitalization and severe disease | Omicron | Mixed variant | 14 | 18-100 | 91·2% [87·2-93·9%] | Moderate |
| HE | Infection + full primary series vaccine vs naive | Pfizer/BioNTech-Comirnaty | Hospitalization and severe disease | Omicron | Mixed variant | 6 [0-13] | 18-100 | 95·5% [67·6-99·4%] | Moderate |
| HE | Infection + full primary series vaccine vs naive | Pfizer/BioNTech-Comirnaty | Hospitalization and severe disease | Omicron | Mixed variant | 42 [14-69] | 18-100 | 92% [88-94·7%] | Moderate |
| HE | Infection + full primary series vaccine vs naive | Pfizer/BioNTech-Comirnaty | Hospitalization and severe disease | Omicron | Mixed variant | 104 [70-139] | 18-100 | 94·7% [93·4-95·7%] | Moderate |
| HE | Infection + full primary series vaccine vs naive | Pfizer/BioNTech-Comirnaty | Hospitalization and severe disease | Omicron | Mixed variant | 140 | 18-100 | 94% [91·4-95·9%] | Moderate |
| VEI | Infection + 1st booster vaccine vs infection | AstraZeneca-Vaxzevria | Any Infection | Omicron | Mixed variant | 6 [0-13] | 18-100 | 56% [53·8-58%] | Moderate |
| VEI | Infection + 1st booster vaccine vs infection | AstraZeneca-Vaxzevria | Any Infection | Omicron | Mixed variant | 70 | 18-100 | 32·6% [29·4-35·7%] | Moderate |

| | | | | | | | | | |
|---|---|---|---|---|---|---|---|---|---|
| VEI | Infection + 1st booster vaccine vs infection | AstraZeneca-Vaxzevria | Any Infection | Omicron | Mixed variant | 42 [14-69] | 18-100 | 60·5% [59·1-61·9%] | Moderate |
| VEI | Infection + 1st booster vaccine vs infection | CoronaVac | Any Infection | Omicron | Mixed variant | 70 | 18-100 | 37·9% [35·8-40%] | Moderate |
| VEI | Infection + 1st booster vaccine vs infection | CoronaVac | Any Infection | Omicron | Mixed variant | 42 [14-69] | 18-100 | 62·7% [61-64·3%] | Moderate |
| VEI | Infection + 1st booster vaccine vs infection | CoronaVac | Any Infection | Omicron | Mixed variant | 6 [0-13] | 18-100 | 54·7% [51·4-57·8%] | Moderate |
| VEI | Infection + 1st booster vaccine vs infection | Janssen-Ad26.COV2.S | Any Infection | Omicron | Mixed variant | 6 [0-13] | 18-100 | 34·1% [26·2-41·1%] | Moderate |
| VEI | Infection + 1st booster vaccine vs infection | Janssen-Ad26.COV2.S | Any Infection | Omicron | Mixed variant | 42 [14-69] | 18-100 | 22·8% [18·8-26·6%] | Moderate |
| VEI | Infection + 1st booster vaccine vs infection | Pfizer/BioNTech-Comirnaty | Any Infection | Omicron | Mixed variant | 70 | 18-100 | 43·3% [25·8-56·6%] | Moderate |
| VEI | Infection + 1st booster vaccine vs infection | Pfizer/BioNTech-Comirnaty | Any Infection | Omicron | Mixed variant | 6 [0-13] | 18-100 | 55·7% [52·3-58·9%] | Moderate |
| VEI | Infection + 1st booster vaccine vs infection | Pfizer/BioNTech-Comirnaty | Any Infection | Omicron | Mixed variant | 42 [14-69] | 18-100 | 56·4% [53·7-59%] | Moderate |
| VEI | Infection + 1st booster vaccine vs infection | AstraZeneca-Vaxzevria | Hospitalization and severe disease | Omicron | Mixed variant | 42 [14-69] | 18-100 | 84·5% [79·4-88·4%] | Moderate |
| VEI | Infection + 1st booster vaccine vs infection | AstraZeneca-Vaxzevria | Hospitalization and severe disease | Omicron | Mixed variant | 6 [0-13] | 18-100 | 79·4% [66·8-87·3%] | Moderate |
| VEI | Infection + 1st booster vaccine vs infection | AstraZeneca-Vaxzevria | Hospitalization and severe disease | Omicron | Mixed variant | 70 | 18-100 | 81·2% [72·5-87·1%] | Moderate |
| VEI | Infection + 1st booster vaccine vs infection | CoronaVac | Hospitalization and severe disease | Omicron | Mixed variant | 70 | 18-100 | 75·7% [69·6-80·7%] | Moderate |
| VEI | Infection + 1st booster vaccine vs infection | CoronaVac | Hospitalization and severe disease | Omicron | Mixed variant | 42 [14-69] | 18-100 | 76·6% [68·1-82·8%] | Moderate |

| | | | | | | | | | |
|---|---|---|---|---|---|---|---|---|---|
| VEI | Infection + 1st booster vaccine vs infection | CoronaVac | Hospitalization and severe disease | Omicron | Mixed variant | 6 [0-13] | 18-100 | 67·9% [32·6-84·7%] | Moderate |
| VEI | Infection + 1st booster vaccine vs infection | Janssen-Ad26.COV2.S | Hospitalization and severe disease | Omicron | Mixed variant | 42 [14-69] | 18-100 | 84% [56·6-94·1%] | Moderate |
| VEI | Infection + 1st booster vaccine vs infection | Janssen-Ad26.COV2.S | Hospitalization and severe disease | Omicron | Mixed variant | 6 [0-13] | 18-100 | 55·2% [1-89%] | Moderate |
| VEI | Infection + 1st booster vaccine vs infection | Pfizer/BioNTech-Comirnaty | Hospitalization and severe disease | Omicron | Mixed variant | 6 [0-13] | 18-100 | 97·8% [64·9-99·9%] | Moderate |
| VEI | Infection + 1st booster vaccine vs infection | Pfizer/BioNTech-Comirnaty | Hospitalization and severe disease | Omicron | Mixed variant | 42 [14-69] | 18-100 | 75% [53·3-86·7%] | Moderate |
| VEI | Infection + full primary series vaccine vs infection | AstraZeneca-Vaxzevria | Any Infection | Omicron | Mixed variant | 6 [0-13] | 18-100 | 43·6% [29·4-54·9%] | Moderate |
| VEI | Infection + full primary series vaccine vs infection | AstraZeneca-Vaxzevria | Any Infection | Omicron | Mixed variant | 104 [70-139] | 18-100 | 14·5% [11·9-17·1%] | Moderate |
| VEI | Infection + full primary series vaccine vs infection | AstraZeneca-Vaxzevria | Any Infection | Omicron | Mixed variant | 42 [14-69] | 18-100 | 25·5% [21-29·7%] | Moderate |
| VEI | Infection + full primary series vaccine vs infection | AstraZeneca-Vaxzevria | Any Infection | Omicron | Mixed variant | 140 | 18-100 | 17% [14·4-19·6%] | Moderate |
| VEI | Infection + full primary series vaccine vs infection | CoronaVac | Any Infection | Omicron | Mixed variant | 140 | 18-100 | 12·3% [9·4-15·1%] | Moderate |
| VEI | Infection + full primary series vaccine vs infection | CoronaVac | Any Infection | Omicron | Mixed variant | 42 [14-69] | 18-100 | 23·4% [18·2-28·3%] | Moderate |
| VEI | Infection + full primary series vaccine vs infection | CoronaVac | Any Infection | Omicron | Mixed variant | 6 [0-13] | 18-100 | 34·5% [20·4-46·1%] | Moderate |
| VEI | Infection + full primary series vaccine vs infection | CoronaVac | Any Infection | Omicron | Mixed variant | 104 [70-139] | 18-100 | 7·3% [4-10·4%] | Moderate |
| VEI | Infection + full primary series vaccine vs infection | Janssen-Ad26.COV2·S | Any Infection | Omicron | Mixed variant | 14 | 18-100 | 16·2% [12·4-19·8%] | Moderate |

| | | | | | | | | | |
|---|---|---|---|---|---|---|---|---|---|
| VEI | Infection + full primary series vaccine vs infection | Pfizer/BioNTech-Comirnaty | Any Infection | Omicron | Mixed variant | 42 [14-69] | 18-100 | 51·9% [50-53·8%] | Moderate |
| VEI | Infection + full primary series vaccine vs infection | Pfizer/BioNTech-Comirnaty | Any Infection | Omicron | Mixed variant | 6 [0-13] | 18-100 | 60·3% [54·3-65·5%] | Moderate |
| VEI | Infection + full primary series vaccine vs infection | Pfizer/BioNTech-Comirnaty | Any Infection | Omicron | Mixed variant | 140 | 18-100 | 26·2% [22·8-29·4%] | Moderate |
| VEI | Infection + full primary series vaccine vs infection | Pfizer/BioNTech-Comirnaty | Any Infection | Omicron | Mixed variant | 104 [70-139] | 18-100 | 32·8% [30·7-34·7%] | Moderate |
| VEI | Infection + full primary series vaccine vs infection | AstraZeneca-Vaxzevria | Hospitalization and severe disease | Omicron | Mixed variant | 42 [14-69] | 18-100 | 41% [1-67·8%] | Moderate |
| VEI | Infection + full primary series vaccine vs infection | AstraZeneca-Vaxzevria | Hospitalization and severe disease | Omicron | Mixed variant | 140 | 18-100 | 55·4% [44·6-64·1%] | Moderate |
| VEI | Infection + full primary series vaccine vs infection | AstraZeneca-Vaxzevria | Hospitalization and severe disease | Omicron | Mixed variant | 104 [70-139] | 18-100 | 57·1% [44·8-66·7%] | Moderate |
| VEI | Infection + full primary series vaccine vs infection | CoronaVac | Hospitalization and severe disease | Omicron | Mixed variant | 42 [14-69] | 18-100 | 34·1% [1-66·3%] | Moderate |
| VEI | Infection + full primary series vaccine vs infection | CoronaVac | Hospitalization and severe disease | Omicron | Mixed variant | 140 | 18-100 | 34·4% [18·3-47·3%] | Moderate |
| VEI | Infection + full primary series vaccine vs infection | CoronaVac | Hospitalization and severe disease | Omicron | Mixed variant | 104 [70-139] | 18-100 | 39·8% [16·9-56·4%] | Moderate |
| VEI | Infection + full primary series vaccine vs infection | Janssen-Ad26.COV2.S | Hospitalization and severe disease | Omicron | Mixed variant | 14 | 18-100 | 39·5% [8·3-60%] | Moderate |
| VEI | Infection + full primary series vaccine vs infection | Pfizer/BioNTech-Comirnaty | Hospitalization and severe disease | Omicron | Mixed variant | 140 | 18-100 | 53·6% [30·2-69·1%] | Moderate |
| VEI | Infection + full primary series vaccine vs infection | Pfizer/BioNTech-Comirnaty | Hospitalization and severe disease | Omicron | Mixed variant | 104 [70-139] | 18-100 | 67·8% [57·4-75·6%] | Moderate |
| VEI | Infection + full primary series vaccine vs infection | Pfizer/BioNTech-Comirnaty | Hospitalization and severe disease | Omicron | Mixed variant | 42 [14-69] | 18-100 | 59·6% [36·6-74·2%] | Moderate |

| | | | | | | | | | | | |
|---|---|---|---|---|---|---|---|---|---|---|---|
| | | VEI | Infection + full primary series vaccine vs infection | Pfizer/BioNTech-Comirnaty | Hospitalization and severe disease | Omicron | Mixed variant | 6 [0-13] | 18-100 | 72·6% [1-96·2%] | Moderate |
| Chin (USA)[14] | Test-negative case-control (Matched based on test week, prison, position, Covid-19 risk score, and room type· Adjusted for age group, and gender) | HE | Infection + 1st booster vaccine vs naive | Pfizer/BioNTech-Comirnaty,Moderna-mRNA-1273 | Any Infection | Omicron | Delta (B.1.617.2) | 68 | 18-100 | 87·4% [81·2-94%] | Moderate |
| | | HE | Infection + 1st booster vaccine vs naive | Pfizer/BioNTech-Comirnaty,Moderna-mRNA-1273 | Any Infection | Omicron | Delta (B.1.617.2) | 59 | 18-100 | 86% [77·2-95·9%] | Moderate |
| | | HE | Infection + full primary series vaccine vs naive | Pfizer/BioNTech-Comirnaty,Moderna-mRNA-1273 | Any Infection | Omicron | Delta (B.1.617.2) | 201 | 18-100 | 72·2% [68-80%] | Moderate |
| | | HE | Infection + full primary series vaccine vs naive | Pfizer/BioNTech-Comirnaty,Moderna-mRNA-1273 | Any Infection | Omicron | Delta (B.1.617.2) | 230 | 18-100 | 74·9% [66·1-86·7%] | Moderate |
| Lind (USA)[15] | Test-negative case-control (Adjusted for date of test, age (continuous), sex, race/ethnicity, comorbidity score, clinical encounters, insurance group, and regional social vulnerability) | VEI | Infection + 1st booster vaccine vs infection | Pfizer/BioNTech-Comirnaty,Moderna-mRNA-1273 | Any Infection | Omicron | Mixed variant | 14 | 5-100 | 45·1% [19-62·8%] | Serious |
| | | VEI | Infection + 1st booster vaccine vs infection | Pfizer/BioNTech-Comirnaty,Moderna-mRNA-1273 | Any Infection | Omicron | Mixed variant | 82 [14-149] | 5-100 | 45·8% [20-63·2%] | Serious |
| | | VEI | Infection + 1st booster vaccine vs infection | Pfizer/BioNTech-Comirnaty,Moderna-mRNA-1273 | Any Infection | Omicron | Mixed variant | 14 | 5-100 | 36% [1-76·2%] | Serious |
| | | VEI | Infection + 1st booster vaccine vs infection | Pfizer/BioNTech-Comirnaty,Moderna-mRNA-1273 | Any Infection | Omicron | Mixed variant | 14 | 5-100 | 38·5% [7·2-59·3%] | Serious |
| | | VEI | Infection + 1st booster vaccine vs infection | Pfizer/BioNTech-Comirnaty,Moderna-mRNA-1273 | Any Infection | Omicron | Mixed variant | 14 | 5-100 | 36·3% [1-76·4%] | Serious |
| | | VEI | Infection + 1st booster vaccine vs infection | Pfizer/BioNTech-Comirnaty,Moderna-mRNA-1273 | Any Infection | Omicron | Mixed variant | 14 | 5-100 | 48·5% [22·2-65·9%] | Serious |
| | | VEI | Infection + 1st booster vaccine vs infection | Pfizer/BioNTech-Comirnaty,Moderna-mRNA-1273 | Any Infection | Omicron | Mixed variant | 14 | 5-100 | 34·3% [1-75·7%] | Serious |
| | | VEI | Infection + 1st booster vaccine vs infection | Pfizer/BioNTech-Comirnaty,Moderna-mRNA-1273 | Any Infection | Omicron | Mixed variant | 14 | 5-100 | 21·3% [1-71%] | Serious |
| | | VEI | Infection + full primary series vaccine vs infection | Pfizer/BioNTech-Comirnaty,Moderna-mRNA-1273 | Any Infection | Omicron | Mixed variant | 82 [14-149] | 5-100 | 37·3% [8·4-57·1%] | Serious |
| | | VEI | Infection + full primary series vaccine vs infection | Pfizer/BioNTech-Comirnaty,Moderna-mRNA-1273 | Any Infection | Omicron | Mixed variant | 150 | 5-100 | 34·2% [18·7-46·8%] | Serious |

| | | | | | | | | | | | |
|---|---|---|---|---|---|---|---|---|---|---|---|
| | | VEI | Infection + full primary series vaccine vs infection | Pfizer/BioNTech-Comirnaty,Moderna-mRNA-1273 | Any Infection | Omicron | Mixed variant | 82 [14-149] | 5-100 | 38·4% [10·5-57·6%] | Serious |
| | | VEI | Infection + full primary series vaccine vs infection | Pfizer/BioNTech-Comirnaty,Moderna-mRNA-1273 | Any Infection | Omicron | Mixed variant | 150 | 5-100 | 33·3% [17·6-45·9%] | Serious |
| | | VEI | Infection + full primary series vaccine vs infection | Pfizer/BioNTech-Comirnaty,Moderna-mRNA-1273 | Any Infection | Omicron | Mixed variant | 82 [14-149] | 5-100 | 30·8% [1-52·4%] | Serious |
| | | VEI | Infection + full primary series vaccine vs infection | Pfizer/BioNTech-Comirnaty,Moderna-mRNA-1273 | Any Infection | Omicron | Mixed variant | 150 | 5-100 | 23·8% [6-38·2%] | Serious |
| | | VEI | Infection + full primary series vaccine vs infection | Pfizer/BioNTech-Comirnaty,Moderna-mRNA-1273 | Any Infection | Omicron | Mixed variant | 14 | 5-100 | 33·2% [3·7-53·6%] | Serious |
| | | VEI | Infection + full primary series vaccine vs infection | Pfizer/BioNTech-Comirnaty,Moderna-mRNA-1273 | Any Infection | Omicron | Mixed variant | 14 | 5-100 | 33·1% [3·6-53·6%] | Serious |
| | | VEI | Infection + full primary series vaccine vs infection | Pfizer/BioNTech-Comirnaty,Moderna-mRNA-1273 | Any Infection | Omicron | Mixed variant | 14 | 5-100 | 32·4% [2·6-53%] | Serious |
| | | VEI | Infection + full primary series vaccine vs infection | Pfizer/BioNTech-Comirnaty,Moderna-mRNA-1273 | Any Infection | Omicron | Mixed variant | 82 [14-149] | 5-100 | 36·1% [7·1-56·1%] | Serious |
| | | VEI | Infection + full primary series vaccine vs infection | Pfizer/BioNTech-Comirnaty,Moderna-mRNA-1273 | Any Infection | Omicron | Mixed variant | 150 | 5-100 | 34% [18·5-46·5%] | Serious |
| | | VEI | Infection + full primary series vaccine vs infection | Pfizer/BioNTech-Comirnaty,Moderna-mRNA-1273 | Any Infection | Omicron | Mixed variant | 14 | 5-100 | 26·2% [1-48·6%] | Serious |
| Medic (Serbia)[21] | Traditional case-control (Matched chorts· Adjusted for age and sex·) | VEI | Infection + 1st booster vaccine vs infection | Pfizer/BioNTech-Comirnaty,Moderna-mRNA-1273,AstraZeneca-Vaxzevria, Gamaleja-Sputnik-V, Gam-COVID-Vac,BBIBP-CorV | Any Infection | Omicron | Mixed variant | 7 | 18-100 | 18·7% [12·3-24·8%] | Serious |
| | | VEI[a] | Infection + 1st booster vaccine vs infection + partial vaccine | Pfizer/BioNTech-Comirnaty,Moderna-mRNA-1273,AstraZeneca-Vaxzevria,Gamaleja-Sputnik-V,BBIBP-CorV | Any Infection | Omicron | Mixed variant | 14 | 18-100 | 24·8% [7·4-39%] | Serious |
| | | VEI[a] | Infection + 1st booster vaccine vs infection + full primary series vaccine | Pfizer/BioNTech-Comirnaty,Moderna-mRNA-1273,AstraZeneca-Vaxzevria,Gamaleja-Sputnik-V,BBIBP-CorV | Any Infection | Omicron | Mixed variant | 7 | 18-100 | 33·3% [27-38·7%] | Serious |
| Nielsen (Denmark)[22] | Retrospective cohort (Adjusted for | VEI | Infection + full primary series vaccine vs infection | Pfizer/BioNTech-Comirnaty,Moderna-mRNA-1273,AstraZeneca-Vaxzevria,Janssen-Ad26.COV2·S | Any Infection | Omicron | Index-Delta | 298 [284-313] | 0-100 | 25·8% [1-46%] | Serious |

| | | | | | | | | | | | |
|---|---|---|---|---|---|---|---|---|---|---|---|
| | age, sex, comorbidity, region of affiliation, staying at hospital, vaccination status, and time since vaccination) | VEI | Infection + full primary series vaccine vs infection | Pfizer/BioNTech-Comirnaty,Moderna-mRNA-1273,AstraZeneca-Vaxzevria,Janssen-Ad26.COV2.S | Any Infection | Omicron | Index-Delta | 328 [314-343] | 0-100 | 24·8% [1-48%] | Serious |
| | | VEI | Infection + full primary series vaccine vs infection | Pfizer/BioNTech-Comirnaty,Moderna-mRNA-1273,AstraZeneca-Vaxzevria,Janssen-Ad26.COV2.S | Any Infection | Omicron | Index-Delta | 344 | 0-100 | 28·6% [1-52%] | Serious |
| | | VEI | Infection + full primary series vaccine vs infection | Pfizer/BioNTech-Comirnaty,Moderna-mRNA-1273,AstraZeneca-Vaxzevria,Janssen-Ad26.COV2.S | Any Infection | Omicron | Index-Delta | 208 [194-223] | 0-100 | 18·4% [11·2-24·8%] | Serious |
| | | VEI | Infection + full primary series vaccine vs infection | Pfizer/BioNTech-Comirnaty,Moderna-mRNA-1273,AstraZeneca-Vaxzevria,Janssen-Ad26.COV2.S | Any Infection | Omicron | Index-Delta | 238 [224-253] | 0-100 | 19·3% [6·9-31·4%] | Serious |
| | | VEI | Infection + full primary series vaccine vs infection | Pfizer/BioNTech-Comirnaty,Moderna-mRNA-1273,AstraZeneca-Vaxzevria,Janssen-Ad26.COV2.S | Any Infection | Omicron | Index-Delta | 268 [254-283] | 0-100 | 37·1% [19·3-51%] | Serious |
| | | VEI | Infection + full primary series vaccine vs infection | Pfizer/BioNTech-Comirnaty,Moderna-mRNA-1273,AstraZeneca-Vaxzevria,Janssen-Ad26.COV2.S | Any Infection | Omicron | Index-Delta | 28 [14-43] | 0-100 | 56·1% [54-58·2%] | Serious |
| | | VEI | Infection + full primary series vaccine vs infection | Pfizer/BioNTech-Comirnaty,Moderna-mRNA-1273,AstraZeneca-Vaxzevria,Janssen-Ad26.COV2.S | Any Infection | Omicron | Index-Delta | 88 [74-103] | 0-100 | 39·1% [35·2-42%] | Serious |
| | | VEI | Infection + full primary series vaccine vs infection | Pfizer/BioNTech-Comirnaty,Moderna-mRNA-1273,AstraZeneca-Vaxzevria,Janssen-Ad26.COV2.S | Any Infection | Omicron | Index-Delta | 118 [104-133] | 0-100 | 30·4% [28·6-33·3%] | Serious |
| | | VEI | Infection + full primary series vaccine vs infection | Pfizer/BioNTech-Comirnaty,Moderna-mRNA-1273,AstraZeneca-Vaxzevria,Janssen-Ad26.COV2.S | Any Infection | Omicron | Index-Delta | 148 [134-163] | 0-100 | 17·5% [14·8-19·3%] | Serious |
| | | VEI | Infection + full primary series vaccine vs infection | Pfizer/BioNTech-Comirnaty,Moderna-mRNA-1273,AstraZeneca-Vaxzevria,Janssen-Ad26.COV2.S | Any Infection | Omicron | Index-Delta | 178 [164-193] | 0-100 | 12·1% [8·6-14·8%] | Serious |
| | | VEI | Infection + full primary series vaccine vs infection | Pfizer/BioNTech-Comirnaty,Moderna-mRNA-1273,AstraZeneca-Vaxzevria,Janssen-Ad26.COV2.S | Any Infection | Omicron | Index-Delta | 58 [44-73] | 0-100 | 46% [43-49%] | Serious |
| Plumb (USA)[23] | Test-negative case-control (Matched cohorts. Adjusted for sex, race/ethnicity, clinical encounters, underlying | VEI | Infection + 1st booster vaccine vs infection | Moderna-mRNA-1273,Pfizer/BioNTech-Comirnaty | Hospitalization and severe disease | Omicron | Mixed variant | 14 | 18-100 | 61·6% [51·4-69·7%] | Moderate |
| | | VEI | Infection + full primary series vaccine vs infection | Pfizer/BioNTech-Comirnaty,Moderna-mRNA-1273 | Hospitalization and severe disease | Omicron | Mixed variant | 14 | 18-100 | 40·3% [30·6-48·6%] | Moderate |

| | | | | | | | | | | | |
|---|---|---|---|---|---|---|---|---|---|---|---|
| | health conditions, and days since the previous infection) | | | | | | | | | | |
| Shrestha (USA)[24] | Retrospective cohort (Adjusted for boosting dose, time since SARS-CoV-2 exposure, time since prior infection, and vaccine doses) | VEI[a] | Infection + 1st booster vaccine vs infection + partial primary series vaccine | Pfizer/BioNTech-Comirnaty,Moderna-mRNA-1273 | Any Infection | Omicron | Index-Delta | 7 | 0-100 | 1% [1-23%] | Serious |
| Šmíd (Czechia)[18] | Traditional case-control (Adjusted for age group, sex, and calendar time) | HE | Infection + 1st booster vaccine vs naive | Pfizer/BioNTech-Comirnaty,AstraZeneca-Vaxzevria,Moderna-mRNA-1273,Janssen-Ad26.COV2.S | Any Infection | Omicron | Delta (B.1.617.2) | 30 [0-60] | 0-100 | 91·4% [88·2-93·6%] | Moderate |
| | | HE | Infection + 1st booster vaccine vs naive | Pfizer/BioNTech-Comirnaty,AstraZeneca-Vaxzevria,Moderna-mRNA-1273,Janssen-Ad26.COV2.S | Any Infection | Omicron | Delta (B.1.617.2) | 61 | 0-100 | 80·8% [70·4-88·2%] | Moderate |
| | | HE | Infection + 1st booster vaccine vs naive | Pfizer/BioNTech-Comirnaty,AstraZeneca-Vaxzevria,Moderna-mRNA-1273,Janssen-Ad26.COV2.S | Any Infection | Omicron | Wild-type,Alpha (B.1.1.7) | 30 [0-60] | 0-100 | 72·5% [71·5-73·5%] | Moderate |
| | | HE | Infection + 1st booster vaccine vs naive | Pfizer/BioNTech-Comirnaty,AstraZeneca-Vaxzevria,Moderna-mRNA-1273,Janssen-Ad26.COV2.S | Any Infection | Omicron | Wild-type,Alpha (B.1.1.7) | 61 | 0-100 | 46·1% [43·1-50·1%] | Moderate |
| | | HE | Infection + 1st booster vaccine vs naive | Pfizer/BioNTech-Comirnaty,AstraZeneca-Vaxzevria,Moderna-mRNA-1273,Janssen-Ad26.COV2.S | Hospitalization and severe disease | Omicron | Wild-type,Alpha (B.1.1.7) | 30 [0-60] | 0-100 | 95% [78-99%] | Moderate |
| | | HE | Infection + 1st booster vaccine vs naive | Pfizer/BioNTech-Comirnaty,AstraZeneca-Vaxzevria,Moderna-mRNA-1273,Janssen-Ad26.COV2.S | Hospitalization and severe disease | Omicron | Wild-type,Alpha (B.1.1.7) | 61 | 0-100 | 90% [64-98%] | Moderate |
| | | HE | Infection + 1st booster vaccine vs naive | Pfizer/BioNTech-Comirnaty,AstraZeneca-Vaxzevria,Moderna-mRNA-1273,Janssen-Ad26.COV2.S | Hospitalization and severe disease | Omicron | Delta (B.1.617.2) | 61 | 0-100 | 71% [1-96%] | Moderate |
| | | HE | Infection + full primary series vaccine vs naive | Pfizer/BioNTech-Comirnaty,AstraZeneca-Vaxzevria,Moderna-mRNA-1273,Janssen-Ad26.COV2.S | Any Infection | Omicron | Delta (B.1.617.2) | 30 [0-60] | 0-100 | 80·8% [73·5-86·1%] | Moderate |
| | | HE | Infection + full primary series vaccine vs naive | Pfizer/BioNTech-Comirnaty,AstraZeneca-Vaxzevria,Moderna-mRNA-1273,Janssen-Ad26.COV2.S | Any Infection | Omicron | Delta (B.1.617.2) | 61 | 0-100 | 85·1% [84-87·2%] | Moderate |

| | | | | | | | | | |
|---|---|---|---|---|---|---|---|---|---|
| HE | Infection + full primary series vaccine vs naive | Pfizer/BioNTech-Comirnaty,AstraZeneca-Vaxzevria,Moderna-mRNA-1273,Janssen-Ad26.COV2.S | Any Infection | Omicron | Wild-type,Alpha (B.1.1.7) | 30 [0-60] | 0-100 | 75·6% [74·6-76·7%] | Moderate |
| HE | Infection + full primary series vaccine vs naive | Pfizer/BioNTech-Comirnaty,AstraZeneca-Vaxzevria,Moderna-mRNA-1273,Janssen-Ad26.COV2.S | Any Infection | Omicron | Wild-type,Alpha (B.1.1.7) | 61 | 0-100 | 43·1% [42·1-44·1%] | Moderate |
| HE | Infection + full primary series vaccine vs naive | Pfizer/BioNTech-Comirnaty,AstraZeneca-Vaxzevria,Moderna-mRNA-1273,Janssen-Ad26.COV2.S | Hospitalization and severe disease | Omicron | Delta (B.1.617.2) | 61 | 0-100 | 93% [49-99%] | Moderate |
| HE | Infection + full primary series vaccine vs naive | Pfizer/BioNTech-Comirnaty,AstraZeneca-Vaxzevria,Moderna-mRNA-1273,Janssen-Ad26.COV2.S | Hospitalization and severe disease | Omicron | Wild-type,Alpha (B.1.1.7) | 30 [0-60] | 0-100 | 94% [77-95%] | Moderate |
| HE | Infection + full primary series vaccine vs naive | Pfizer/BioNTech-Comirnaty,AstraZeneca-Vaxzevria,Moderna-mRNA-1273,Janssen-Ad26.COV2.S | Hospitalization and severe disease | Omicron | Wild-type,Alpha (B.1.1.7) | 61 | 0-100 | 73% [78-99%] | Moderate |

n=13 studies. VEI=Ve-infected, PEV=PE-Vaccinated. [a]VE-Infected indicates the estimate used comparison of hybrid immunity to hybrid immunity.

#### S8. Risk of bias Assessment using the ROBINS-I tool for observational studies

##### a) Prior infection studies

| First author (Country) | Estimates in analysis (n) | Variant | Outcome | Bias dut to confounding | Bias due to participant selection | Bias due to intervention classification | Bias due to deviations from intended interventions | Bias due to missing data | Bias in measurement of outcomes | Bias due to selection of reported result | Overall Risk of bias |
|---|---|---|---|---|---|---|---|---|---|---|---|
| Altarawneh 1 (Qatar)[9] | 3 | Omicron | All infections | Moderate | Low | Low | Low | Serious | Low | Moderate | Serious |
| | 1 | Omicron | Hospitalization and severe disease | Moderate | Low | Low | Low | Moderate | Moderate | Moderate | Moderate |
| Altarawneh 2 (Qatar)[10] | 1 | Omicron | All infections (Symptomatic) | Moderate | Low | Low | Low | Moderate | Moderate | Moderate | Moderate |
| | 1 | Omicron | Hospitalization and severe disease | Moderate | Low | Low | Low | Moderate | Low | Moderate | Moderate |
| | 1 | Omicron (BA.1) | All infections (Symptomatic) | Moderate | Low | Low | Low | Moderate | Moderate | Moderate | Moderate |
| | 1 | Omicron (BA.1) | Hospitalization and severe disease | Moderate | Low | Low | Low | Moderate | Low | Moderate | Moderate |
| | 1 | Omicron (BA.2) | All infections (Symptomatic) | Moderate | Low | Low | Low | Moderate | Moderate | Moderate | Moderate |
| | 1 | Omicron (BA.2) | Hospitalization and severe disease | Moderate | Low | Low | Low | Moderate | Low | Moderate | Moderate |
| Andeweg (Netherland)[11] | 10 | Omicron (BA.1) | All infections | Moderate | Serious | Moderate | Low | Low | Low | Moderate | Serious |
| | 10 | Omicron (BA.1) | All infections (Symptomatic) | Moderate | Serious | Moderate | Low | Low | Low | Moderate | Serious |
| | 5 | Omicron (BA.2) | All infections | Moderate | Serious | Moderate | Low | Low | Low | Moderate | Serious |
| | 5 | Omicron (BA.2) | All infections (Symptomatic) | Moderate | Serious | Moderate | Low | Low | Low | Moderate | Serious |
| Carazo (Canada)[12] | 7 | Omicron | All infections | Moderate | Serious | Low | Low | Low | Low | Moderate | Serious |

| | | | | | | | | | | | |
|---|---|---|---|---|---|---|---|---|---|---|---|
| | 2 | Omicron | All infections (Symptomatic) | Moderate | Low | Low | Low | Serious | Low | Moderate | Serious |
| | 2 | Omicron | Hospitalization and severe disease | Moderate | Low | Low | Low | Low | Low | Moderate | Moderate |
| Cerqueira-Silva (Brazil)[13] | 3 | Omicron | All infections (Symptomatic) | Low | Low | Low | Low | Low | Low | Moderate | Moderate |
| | 3 | Omicron | Hospitalization | Moderate | Serious | Low | Low | Moderate | Moderate | Moderate | Serious |
| Chin (USA)[14] | 2 | Omicron | All infections | Low | Low | Low | Low | Low | Low | Moderate | Moderate |
| Lind (USA)[15] | 1 | Omicron | All infections | Moderate | Low | Low | Low | Moderate | Moderate | Moderate | Moderate |
| Michlmayr (Denmark)[16] | 4 | Omicron | All infections | Moderate | Moderate | Low | Low | Low | Low | Serious | Serious |
| | 4 | Omicron | All infections (Symptomatic) | Moderate | Moderate | Low | Low | Low | Serious | Serious | Serious |
| | 1 | Omicron | Hospitalization and severe disease | Moderate | Moderate | Low | Low | Low | Low | Serious | Serious |
| Nyberg (UK)[17] | 8 | Omicron | Hospitalization and severe disease | Serious | Moderate | Serious | Low | Serious | Low | Moderate | Serious |
| | 8 | Omicron | Severe disease | Serious | Moderate | Serious | Low | Serious | Low | Moderate | Serious |
| Šmíd (Czech)[18] | 6 | Omicron | All infections | Moderate | Low | Low | Low | Moderate | Low | Moderate | Moderate |
| | 6 | Omicron | Hospitalization and severe disease | Moderate | Low | Low | Low | Moderate | Low | Moderate | Moderate |

#### b) Hybrid immunity study

| First author (country) | Measure | Estimates in analysis (n) | Outcome | Variant | Bias dut to confounding | Bias due to participant selection | Bias due to intervention classification | Bias due to deviations from intended interventions | Bias due to missing data | Bias in measurement of outcomes | Bias due to selection of reported result | Overall risk of bias |
|---|---|---|---|---|---|---|---|---|---|---|---|---|
| Altarawneh 2 (Qatar)[10] | HE | 7 | All infection (symptomatic) | Omicron (BA.1, BA.2) | Moderate | Low | Low | Low | Low | Low | Moderate | Moderate |
| Andeweg (Netherland)[11] | HE | 21 | All infection | Omicron (BA.1) | Moderate | Serious | Moderate | Low | Low | Low | Moderate | Serious |
| | HE | 21 | All infection (symptomatic) | Omicron (BA.1) | Moderate | Serious | Moderate | Low | Low | Low | Moderate | Serious |
| | HE | 11 | All infection | Omicron (BA.2) | Moderate | Serious | Moderate | Low | Low | Low | Moderate | Serious |
| | HE | 11 | All infection (symptomatic) | Omicron (BA.2) | Moderate | Serious | Moderate | Low | Low | Low | Moderate | Serious |
| Björk (Sweden)[19] | HE | 3 | Hospitalization and severe disease | Omicron | Moderate | Low | Low | Low | Low | Low | Serious | Serious |
| Bruel (France)[20] | PEV | 1 | All infection | Omicron | Serious | Low | Low | Low | Moderate | Low | Moderate | Serious |
| Carazo (Canada)[12] | HE,VEI[a] | 11 | All infection | Omicron | Moderate | Serious | Low | Low | Serious | Low | Moderate | Serious |
| | HE,VEI | 12 | Hospitalization and severe disease | Omicron | Moderate | Low | Low | Low | Low | Low | Moderate | Moderate |
| Cerqueira-Silva (Brazil)[13] | HE,VEI | 48 | All infection (symptomatic) | Omicron | Low | Moderate | Low | Low | Low | Low | Moderate | Moderate |
| | HE,VEI | 42 | Hospitalization and severe disease | Omicron | Low | Low | Low | Low | Low | Moderate | Moderate | Moderate |

| | | | | | | | | | | | | |
|---|---|---|---|---|---|---|---|---|---|---|---|---|
| Chin (USA)[14] | HE | 4 | All infection | Omicron | Low | Low | Low | Low | Low | Low | Moderate | Moderate |
| Lind (USA)[15] | VEI | 20 | All infection | Omicron | Moderate | Low | Low | Low | Moderate | Serious | Moderate | Serious |
| Medić (Serbia)[21] | VEI[a] | 3 | All infection | Omicron | Serious | Moderate | Low | Low | Low | Moderate | Moderate | Serious |
| Nielsen (Denmark)[22] | VEI | 12 | All infection | Omicron | Moderate | Moderate | Low | Low | Low | Serious | Moderate | Serious |
| Plumb (USA)[23] | VEI | 2 | Hospitalization | Omicron | Moderate | Low | Moderate | Low | Low | Low | Moderate | Moderate |
| Shrestha (USA)[24] | VEI[a] | 1 | All infection | Omicron | Serious | Moderate | Serious | Low | Low | Serious | Moderate | Serious |
| Šmíd (Czech)[18] | HE | 8 | All infection | Omicron | Moderate | Low | Low | Low | Moderate | Low | Moderate | Moderate |
| | HE | 6 | Hospitalization | Omicron | Moderate | Low | Low | Low | Moderate | Low | Moderate | Moderate |

VEI=Ve-infected, PEV=PE-Vaccinated. [a]VE-Infected includes results from comparisons of hybrid immunity to prior infection or hybrid immunity.

**S9. Sensitivity analysis of protection conferred by prior infection or hybrid immunity over time using the WHO definition of severe disease.**

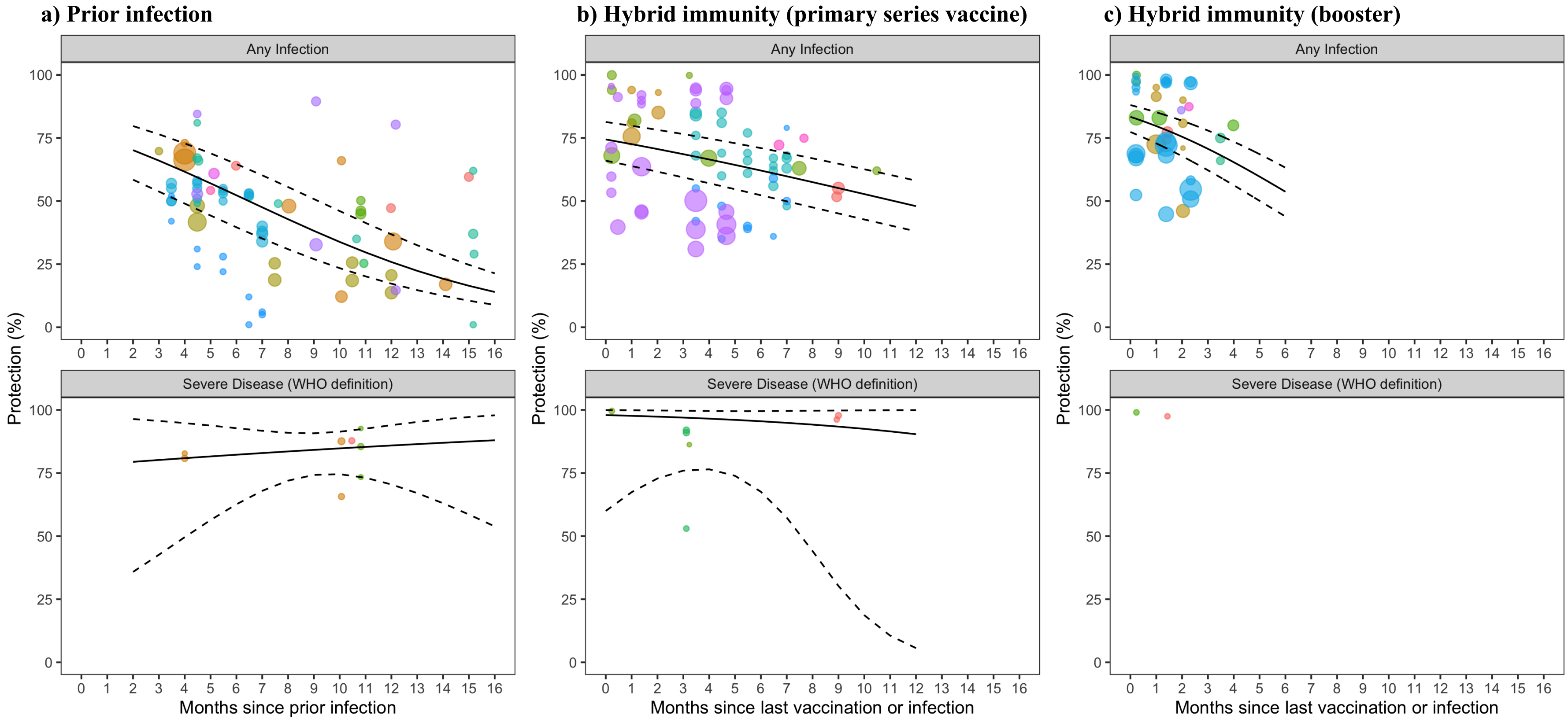

Points of the same color represent estimates from the same study. The diameter of points varies with the sample size of the study.

#### S10. Sensitivity analysis of the protection against reinfection and severe disease conferred by the primary-series vaccine, first booster vaccine, prior infection, and hybrid immunity compared to immune naive

| Severity of Infection | No. studies | Month 1[a] | Month 2[b] | Month 3 | Month 4 | Month 6 | Month 9 | Month 12 | Month 15 | Percentage point change in protection from 3 to 6 months [95% CI][c] | Percentage point change in protection from 3 to 12 months [95% CI][c] |
|---|---|---|---|---|---|---|---|---|---|---|---|
| **Prior Infection** | | | | | | | | | | | |
| Any Infection[d] | 10 | NA | 70·2% [58·5-79·7%] | 66% [53·8-76·5%] | 61·6% [49-72·8%] | 52·3% [39·6-64·7%] | 38·2% [27-50·8%] | 25·8% [17·3-36·8%] | 16·4% [10·5-24·7%] | -14·0 [-12·0 to -18·2] | -40·5 [-33·9 to -51·9] |
| Severe Disease | 3 | NA | 79·4% [35·8-96·4%] | 80·2% [42·6-95·7%] | 80·9% [49·6-94·8%] | 82·3% [62·6-92·8%] | 84·2% [74·3-90·7%] | 85·9% [70·5-94%] | 87·5% [58·6-97·2%] | -2·4 [-4·7 to +5·1] | -7·8 [-12·1 to +20·9] |
| **Hybrid Immunity (primary series)** | | | | | | | | | | | |
| Any Infection | 7 | 72·6% [63·9-79·8%] | 70·6% [61·6-78·2%] | 68·6% [59·4-76·5%] | 66·5% [57-74·8%] | 62·1% [52·3-71%] | 55·1% [45·1-64·8%] | 48·0% [38·0-58·1%] | NA | -6·5 [-15·1 to +4·9] | -20·6 [-48·8 to +11·4] |
| Severe Disease | 3 | 97·7% [67·4-99·9%] | 97·4% [72·8-99·8%] | 97% [76-99·7%] | 96·6% [76·5-99·6%] | 95·5% [67·6-99·5%] | 93·4% [30·3-99·8%] | 90·4% [5·6-99·9%] | NA | -1·5 [-3·4 to +11·5] | -6·6 [-20·9 to +17·8] |
| **Hybrid Immunity (booster)** | | | | | | | | | NA | | |
| Any Infection | 6 | 79·7% [72·9-85·2%] | 75·5% [67·8-81·8%] | 70·7% [62·2-77·9%] | 65·4% [56·3-73·5%] | 53·7% [43·9-63·2%] | NA | NA | NA | NA | NA |
| Severe Disease | 2 | 98·2% [85·5-99·8%] | 96·1% [17·4-100%] | NA | NA | NA | NA | NA | NA | NA | NA |

This table displays the data shown in Figure S9. This analysis uses the same log-odds meta-regression model as Figure 2. [a]Month 1 data are for persons with hybrid immunity whose last immunological challenge was vaccination and thus were eligible for reinfection within a shorter time frame than people who most recently had prior infection (2 month minimum for probable reinfection). [b]Month 2 data represent the minimum time period for an infection among persons with prior infection [c]Confidence intervals calculated using the bootstrap method. [d]Any infections contains mild infections, symptomatic infections and asymptomatic infections. NA: insufficient data for model extrapolation. Prior infection data is available for 2-16 month predictions; hybrid immunity data was available for 1-11 month predictions. Data were extrapolated to a maximum of 3 months beyond the final follow-up date.

#### S11. Severe disease definitions from included articles

| First Author (Country) | Measure | Sensitivity defined as severe, critical, or fatal COVID-19 | Severe disease definition |
|---|---|---|---|
| Altarawneh 1 (Qatar)[9] | Prior infection | Yes | Severe, critical, or fatal COVID-19 as defined per the WHO classification. |
| Altarawneh 2 (Qatar)[10] | Prior infection and Hybrid immunity | Yes | Severe, critical, or fatal COVID-19 as defined per the WHO classification. |
| Andeweg (Netherland)[11] | Prior infection and Hybrid immunity | No | N/A |
| Björk (Sweden)[19] | Hybrid immunity | Yes | A case who was hospitalised for at least 24 h from 5 days before until 14 days after a positive SARS-CoV-2 test and required oxygen supply (≥ 5 L/min) or admittance to an ICU. |
| Bruel (France)[20] | Hybrid immunity | No | N/A |
| Carazo (Canada)[12] | Prior infection and Hybrid immunity | No | COVID-19 hospitalization, defined by admission, ≥24-hours and within 14 days of a SARS-CoV-2 positive specimen |
| | | Yes | COVID-19 death |
| Cerqueira-Silva (Brazil)[13] | Prior infection and Hybrid immunity | No | N/A |
| Chin (USA)[14] | Prior infection and Hybrid immunity | No | N/A |
| Lind (USA)[15] | Prior infection and Hybrid immunity | No | N/A |
| Medić (Serbia)[21] | Prior infection and Hybrid immunity | Yes | COVID-19 pneumonia confirmed by chest imaging |
| | | Yes | COVID-19 pneumonia required mechanical ventilation and/or admission to the ICU |
| Michlmayr (Denmark)[16] | Prior infection | No | Hospital admission associated with ICD-10 primary diagnosis codes occurring no earlier than two days before, and no later than 14 days after a positive RT-PCR test. |
| Nielsen (Denmark)[22] | Hybrid immunity | No | N/A |
| Nyberg (UK)[17] | Prior infection | No | Any hospital attendances, including admissions and attendances at accident and emergency departments, 0–14 days after the first specimen date of the most recent infection episode. |
| | Prior infection | No | Hospital attendances, admissions and diagnoses during hospital stay |
| | | Yes | Death occurring 0–28 days after the first positive specimen date of the most recent infection episode, again matching the definition used in routine UK government reporting. |
| Plumb (USA)[23] | Hybrid immunity | No | At least one hospital admission for a COVID-19-like illness, with a hospitalization-associated NAAT performed from 10 days before through 3 days after admission. |
| | | No | COVID-19-like illness: acute respiratory illness or related signs or symptomsusing diagnosis codes from the ICD-10 |

| | | | |
|---|---|---|---|
| Shrestha (USA)[24] | Hybrid immunity | No | N/A |
| Šmíd (Czech)[18] | Prior infection and Hybrid Immunity | No | Hospital admission of a person, who tested positive on a PCR test, within two weeks after the confirmed infection or earlier |
| | | Yes | Admission to ICU during the hospitalization. |
| | | Yes | Use of any type of oxygen therapy |

#### S12. Summary of results for studies reporting sub-group data by age

| Study | Age group | Exposure and comparator | Time since last immunological hit | Vaccine | Protection against any infection [95% CI] |
|---|---|---|---|---|---|
| Carazo (Canada)[12] | 12-17 | Prior infection vs naive | 90-730 days | N/A | 57% [36-71%] |
| Carazo (Canada) | 18-49 | Prior infection vs naïve | 90-730 days | N/A | 44% [29-43%] |
| Carazo (Canada) | 50-69 | Prior infection vs naïve | 90-730 days | N/A | 51% [38-60%] |
| Carazo (Canada) | 70+ | Prior infection vs naïve | 90-730 days | N/A | 46% [16-65%] |
| Carazo (Canada) | 12-17 | Infection + partial primary series vaccine vs naïve | 21 days | Mixed | 78% [70-83%] |
| Carazo (Canada) | 18-49 | Infection + partial primary series vaccine vs naïve | 21 days | Mixed | 62% [60-65%] |
| Carazo (Canada) | 50-69 | Infection + partial primary series vaccine vs naïve | 21 days | Mixed | 71% [66-75%] |
| Carazo (Canada) | 70+ | Infection + partial primary series vaccine vs naïve | 21 days | Mixed | 79% [65-87%] |
| Carazo (Canada) | 12-17 | Infection + primary series vs naïve | 7 days | Mixed | 79% [74-93%] |
| Carazo (Canada) | 18-49 | Infection + primary series vs naïve | 7 days | Mixed | 67% [65-68%] |
| Carazo (Canada) | 50-69 | Infection + primary series vs naïve | 7 days | Mixed | 72% [69-74%] |
| Carazo (Canada) | 70+ | Infection + primary series vs naïve | 7 days | Mixed | 67% [60-73%] |
| Carazo (Canada) | 12-17 | Infection + 1$^{st}$ booster vaccine vs naïve | 7 days | Mixed | 96% [65-99%] |
| Carazo (Canada) | 18-49 | Infection + 1$^{st}$ booster vaccine vs naïve | 7 days | Mixed | 79% [77-81%] |
| Carazo (Canada) | 50-69 | Infection + 1$^{st}$ booster vaccine vs naïve | 7 days | Mixed | 86% [83-88%] |
| Carazo (Canada) | 70+ | Infection + 1$^{st}$ booster vaccine vs naïve | 7 days | Mixed | 81% [75-86%] |
| Andeweg (Netherlands) (Jan to Mar 2022 Cohort, BA.1 infections)[11] | 0-11 | Prior infection vs naïve | 180 days | N/A | 41% [34-48%] |
| Andeweg (Netherlands) (Jan to Mar 2022 Cohort, BA.1 infections) | 12-17 | Prior infection vs naïve | 180 days | N/A | 39% [30-46%] |
| Andeweg (Netherlands) (Jan to Mar 2022 Cohort, BA.1 infections) | 18-29 | Prior infection vs naive | 180 days | N/A | 37% [32-42%] |
| Andeweg (Netherlands) (Jan to Mar 2022 Cohort, BA.1 infections) | 30-59 | Prior infection vs naïve | 180 days | N/A | 35% [31-40%] |
| Andeweg (Netherlands) (Jan to Mar 2022 Cohort, BA.1 infections) | 60+ | Prior infection vs naïve | 180 days | N/A | 45% [30-57%] |
| Andeweg (Netherlands) (Jan to Mar 2022 Cohort, BA.1 infections) | 12-17 | Infection + primary series vs naïve | 180 days | Mixed | 69% [44-83%] |
| Andeweg (Netherlands) (Jan to Mar 2022 Cohort, BA.1 infections) | 18-29 | Infection + primary series vs naïve | 180 days | Mixed | 65% [57-72%] |

| | | | | | |
|---|---|---|---|---|---|
| Andeweg (Netherlands) (Jan to Mar 2022 Cohort, BA.1 infections) | 30-59 | Infection + primary series vs naïve | 180 days | Mixed | 58% [51-65%] |
| Andeweg (Netherlands) (Jan to Mar 2022 Cohort, BA.1 infections) | 60+ | Infection + primary series vs naive | 180 days | Mixed | 71% [48-83%] |
| Andeweg (Netherlands) (Jan to Mar 2022 Cohort, BA.2 infections) | 0-11 | Prior infection vs naïve | 180 days | N/A | 35% [23-46%] |
| Andeweg (Netherlands) (Jan to Mar 2022 Cohort, BA.2 infections) | 12-17 | Prior infection vs naïve | 180 days | N/A | 54% [43-63%] |
| Andeweg (Netherlands) (Jan to Mar 2022 Cohort, BA.2 infections) | 18-29 | Prior infection vs naïve | 180 days | N/A | 43% [37-48%] |
| Andeweg (Netherlands) (Jan to Mar 2022 Cohort, BA.2 infections) | 30-59 | Prior infection vs naïve | 180 days | N/A | 37% [30-42%] |
| Andeweg (Netherlands) (Jan to Mar 2022 Cohort, BA.2 infections) | 60+ | Prior infection vs naive | 180 days | N/A | 45% [24-61%] |
| Andeweg (Netherlands) (Jan to Mar 2022 Cohort, BA.2 infections) | 12-17 | Infection + primary series vs naïve | 180 days | Mixed | 81% [56-92%] |
| Andeweg (Netherlands) (Jan to Mar 2022 Cohort, BA.2 infections) | 18-29 | Infection + primary series vs naïve | 180 days | Mixed | 67% [57-75%] |
| Andeweg (Netherlands) (Jan to Mar 2022 Cohort, BA.2 infections) | 30-59 | Infection + primary series vs naïve | 180 days | Mixed | 62% [52-69%] |
| Andeweg (Netherlands) (Jan to Mar 2022 Cohort, BA.2 infections) | 60+ | Infection + primary series vs naïve | 180 days | Mixed | 36% [1-64%] |
| Andeweg (Netherlands) (Nov 2021 to Jan 2022 Cohort)[11] | 0-11 | Prior infection vs naïve | 180 days | N/A | 42% [16-60%] |
| Andeweg (Netherlands) (Nov 2021 to Jan 2022 Cohort) | 12-17 | Prior infection vs naive | 180 days | N/A | 37% [6-58%] |
| Andeweg (Netherlands) (Nov 2021 to Jan 2022 Cohort) | 18-29 | Prior infection vs naïve | 180 days | N/A | 7% [-8-20%] |
| Andeweg (Netherlands) (Nov 2021 to Jan 2022 Cohort) | 30-59 | Prior infection vs naïve | 180 days | N/A | 5% [-13-20%] |
| Andeweg (Netherlands) (Nov 2021 to Jan 2022 Cohort) | 60+ | Prior infection vs naive | 180 days | N/A | 20% [-49-57%] |
| Andeweg (Netherlands) (Nov 2021 to Jan 2022 Cohort) | 12-17 | Infection + primary series vs naïve | 180 days | Mixed | 50% [1-94%] |
| Andeweg (Netherlands) (Nov 2021 to Jan 2022 Cohort) | 18-29 | Infection + primary series vs naïve | 180 days | Mixed | 54% [35-67%] |
| Andeweg (Netherlands) (Nov 2021 to Jan 2022 Cohort) | 30-59 | Infection + primary series vs naïve | 180 days | Mixed | 50% [31-64%] |
| Andeweg (Netherlands) (Nov 2021 to Jan 2022 Cohort) | 60+ | Infection + primary series vs naive | 180 days | Mixed | 69% [27-87%] |

#### S13. Protective effectiveness of hybrid immunity by prior infection variant

| Prior infection variant | Exposure | Comparator | Severity | Number of studies | Month 3 | Month 6 |
|---|---|---|---|---|---|---|
| **Prior infection** | | | | | | |
| Alpha | Infection | Naïve | Any Infection | 1 | 64·2% [60·3-67·9%] | 52·8% [50·1-55·5%] |
| Delta | Infection | Naïve | Any Infection | 3 | 65·5% [29·5-89·6%] | 55·3% [22·0-84·4%] |
| Mixed variant | Infection | Naïve | Any Infection | 8 | 60·8% [44·6-75·0%] | 47·4% [31·9-63·5%] |
| Alpha | Infection | Naïve | Hospitalization or severe disease | 1 | - | 66·0% [54·0-75·0%][c] |
| Delta | Infection | Naïve | Hospitalization or severe disease | 2 | 74·7% [60·6-85·1%] | - |
| Mixed variant | Infection | Naïve | Hospitalization or severe disease | 5 | 77·7% [62·5-87·9%] | 77·4% [66·6-85·5%] |
| **Hybrid immunity** | | | | | | |
| Alpha | Infection + primary series | Naïve | Any Infection | 1 | 75·6% [74·6-76·7%][a] | - |
| Delta | Infection + primary series | Naïve | Any Infection | 5 | 79·6% [61·8-90·4%] | 62·4% [40·6-80·1%] |
| Mixed variant | Infection + primary series | Naïve | Any Infection | 5 | 63·7% [53·9-72·5%] | 54·8% [44·6-64·5%] |
| Alpha | Infection + primary series | Naïve | Hospitalization or severe disease | 1 | 94·0% [77·0-95·0%][a] | - |
| Delta | Infection + primary series | Naïve | Hospitalization or severe disease | 1 | 93·0% [49·0-99·0%][b] | - |
| Mixed variant | Infection + primary series | Naïve | Hospitalization or severe disease | 4 | 96·4% [86·9-99·1%] | 96·8% [88·3-99·2%] |

[a]Single estimate at month 1 from Šmíd 2 (Czechia). [b]Single estimate at month 2 from Šmíd 2 (Czechia).[c]Single estimate at month 10 from Šmíd 1 (Czechia)

#### S14. Protective effectiveness of hybrid immunity by vaccine type

| Vaccine type | Exposure | Comparator | Severity | No. studies | Month 3 | Month 6 | Percentage point change in protection from 3 to 6 months [95% CI] |
|---|---|---|---|---|---|---|---|
| mRNA[a] | Prior infection + primary series | Naïve | Any Infection | 5 | 68·8% [56·9-78·6%] | 60·9% [48·3-72·2%] | -7·9 [-2·0 to -15·7] |
| NRVV[b] | Prior infection + primary series | Naïve | Any Infection | 1 | 38·8% [37·7-39·8%][e] | 40·7% [39·6-41·7%][f] | - |
| Inactivated[c] | Prior infection + primary series | Naïve | Any Infection | 1 | 31·0% [29·4-32·5%][e] | 36·2% [34·9-37·4%][f] | - |
| Mixed (NRVV + mRNA)[d] | Prior infection + primary series | Naïve | Any Infection | 3 | 70·8% [55·1-82·8%] | 60·1% [43·0-75·1%] | -10·7 [-4·5 to +24·8] |
| mRNA[a] | Prior infection + primary series | Naïve | Hospitalization or severe disease | 3 | 97·7% [89·5-99·5%] | 98·1% [91·2-99·6%] | -0·47 [-1·4 to +2·0] |
| NRVV[b] | Prior infection + primary series | Naïve | Hospitalization or severe disease | 1 | 93·9% [92·8-94·9%][e] | 94·5% [93·8-95·1%][f] | - |
| Inactivated[c] | Prior infection + primary series | Naïve | Hospitalization or severe disease | 1 | 88·4% [77·9-93·9%][e] | 90·7% [89·5-91·8%][f] | - |
| Mixed (NRVV + mRNA)[d] | Prior infection + primary series | Naïve | Hospitalization or severe disease | 2 | 89·2% [73·9-96·0%] | 75·5% [13·7-98·4%] | -13·6 [-67·0 to -3·8] |

[a]mRNA vaccine type contain Pfizer and Moderna; [b]NRVV (Non-replicating viral vectors) vaccine type contain AstraZeneca; [c]Inactivated type of vaccine refers to CoronaVac; [d]Mixed vaccine type refers to Pfizer, Moderna, AstraZeneca, and Johnson & Johnson (i.e., mRNA and NRVV). [e]Single estimate at week 10-19 from Cerqueira-Silva (Brazil). [f]Single estimate at over 20 weeks from Cerqueira-Silva (Brazil).

**S15. Six-month protection against reinfection and severe disease conferred by the primary-series vaccine, first booster vaccine, prior infection, and hybrid immunity compared to immune naïve individuals**

| Group | Number of studies | Six-month protection | p-value |
|---|---|---|---|
| **Any infection** | | | |
| Primary series vaccine | 15 | 15·1% [11·3-19·8%] | <0·0001 |
| First booster vaccine | 9 | 24·8% [18·5-32·5%] | <0·0001 |
| Prior infection | 10 | 51·2% [38·6-63·7%] | 0·28 |
| Hybrid immunity (primary series vaccine) | 7 | 60·4% [49·6-70·3%] | *ref* |
| Hybrid immunity (first booster) | 6 | 46·5% [36·0-57·3%] | 0·08 |
| **Hospitalization or severe disease** | | | |
| Primary series vaccine | 12 | 64·6% [54·5-73·6%] | <0·0001 |
| First booster vaccine | 10 | 76·7% [72·5-80·4%] | <0·0001 |
| Prior infection | 6 | 80·1% [70·3-87·2%] | 0·01 |
| Hybrid immunity (primary series vaccine) | 5 | 96·5% [90·2-98·8%] | *ref.* |
| Hybrid immunity (first booster) | 4 | 95·3% [81·9-98·9%] | 0·75 |

To obtain six-month protection, we ran a log-odds meta-regression model on all data allowing for different slopes and intercepts for each group and a random intercept for each study. We centered the month variable by subtracting six from the number of months such that the intercept for each group represents the protection at six months. See also Figure 3. Vaccine effectiveness data (primary series vaccine, first booster vaccine) are from a previously published systematic review.[6]

3. World Health Organization. *WHO COVID-19: Case Definitions*.; 2020. file:///Users/niklasbobrovitz/Downloads/WHO-2019-nCoV-Surveillance_Case_Definition-2020.2-eng.pdf

4. World Health Organization. *Living Guidance for Clinical Management of COVID-19*.; 2021. https://apps.who.int/iris/bitstream/handle/10665/349321/WHO-2019-nCoV-clinical-2021.2-eng.pdf

5. World Health Organization. *International Guidelines for Certification and Classification (Coding) of COVID-19 as Cause of Death.* https://www.who.int/publications/m/item/international-guidelines-for-certification-and-classification-(coding)-of-covid-19-as-cause-of-death
